# Concerted changes in the pediatric single-cell intestinal ecosystem before and after anti-TNF blockade

**DOI:** 10.1101/2021.09.17.21263540

**Authors:** Hengqi Betty Zheng, Benjamin A. Doran, Kyle Kimler, Alison Yu, Victor Tkachev, Veronika Niederlova, Kayla Cribbin, Ryan Fleming, Brandi Bratrude, Kayla Betz, Lorenzo Cagnin, Connor McGuckin, Paula Keskula, Alexandre Albanese, Maria Sacta, Joshua de Sousa Casal, Ruben van Esch, Andrew C. Kwong, Conner Kummerlowe, Faith Taliaferro, Nathalie Fiaschi, Baijun Kou, Sandra Coetzee, Sumreen Jalal, Yoko Yabe, Michael Dobosz, Matthew F. Wipperman, Sara Hamon, George D. Kalliolias, Andrea Hooper, Wei Keat Lim, Sokol Haxhinasto, Yi Wei, Madeline Ford, Lusine Ambartsumyan, David L. Suskind, Dale Lee, Gail Deutsch, Xuemei Deng, Lauren V. Collen, Vanessa Mitsialis, Scott B. Snapper, Ghassan Wahbeh, Alex K. Shalek, Jose Ordovas-Montanes, Leslie S. Kean

## Abstract

Crohn’s disease is an inflammatory bowel disease (IBD) commonly treated through anti-TNF blockade. However, most patients still relapse and inevitably progress. Comprehensive single-cell RNA-sequencing (scRNA-seq) atlases have largely sampled patients with established treatment-refractory IBD, limiting our understanding of which cell types, subsets, and states at diagnosis anticipate disease severity and response to treatment. Here, through combining clinical, flow cytometry, histology, and scRNA-seq methods, we profile diagnostic human biopsies from the terminal ileum of treatment-naïve pediatric patients with Crohn’s disease (pediCD; n=14), matched repeat biopsies (pediCD-treated; n=8) and from non-inflamed pediatric controls with functional gastrointestinal disorders (FGID; n=13). To resolve and annotate epithelial, stromal, and immune cell states among the 201,883 baseline single-cell transcriptomes, we develop a principled and unbiased tiered clustering approach, ARBOL. Through flow cytometry and scRNA-seq, we observe that treatment-naïve pediCD and FGID have similar broad cell type composition. However, through high-resolution scRNA-seq analysis and microscopy, we identify significant differences in cell subsets and states that arise during pediCD relative to FGID. By closely linking our scRNA-seq analysis with clinical meta-data, we resolve a vector of T cell, innate lymphocyte, myeloid, and epithelial cell states in treatment-naïve pediCD (pediCD-TIME) samples which can distinguish patients along the trajectory of disease severity and anti-TNF response. By using ARBOL with integration, we position repeat on-treatment biopsies from our patients between treatment-naïve pediCD and on-treatment adult CD. We identify that anti-TNF treatment pushes the pediatric cellular ecosystem towards an adult, more treatment-refractory state. Our study jointly leverages a treatment-naïve cohort, high-resolution principled scRNA-seq data analysis, and clinical outcomes to understand which baseline cell states may predict Crohn’s disease trajectory.

## Main

Inflammatory bowel diseases (IBDs) arise when homeostatic mechanisms regulating gastrointestinal (GI) tract tissue integrity, nutrient absorption, and protective immunity are replaced by pathogenic inflammation (Baumgart and Sandborn, 2012; Chang, 2020; Corridoni et al., 2020b; Friedrich et al., 2019; Graham and Xavier, 2020; Selin et al., 2021). The initiating triggers are not fully known, but host genetics and the microbiome are being increasingly appreciated to play important, and in some cases causal roles in the IBDs (Chang, 2020; Cohen et al., 2019; Franzosa et al., 2019; Jain et al., 2021; Limon et al., 2019). Crohn’s disease (CD) presents predominantly in the terminal ileum and the proximal colon, though lesions may develop anywhere along the gastrointestinal tract (Baumgart and Sandborn, 2012; Chang, 2020; Kobayashi et al., 2020; Roda et al., 2020). Among the IBDs, pediatric-onset Crohn’s disease (pediCD) is particularly common (25% of all IBD cases, 60-70% of pediatric IBD) and is a debilitating form due to its early presentation, impact on the terminal ileum and proximal colon, and the lack of disease-specific therapies developed for children (Hyams et al., 1991; Ruemmele et al., 2014; Sýkora et al., 2018; Turner et al., 2012; Ye et al., 2020). In contrast to pediCD, there exists a group of pediatric disorders termed functional gastrointestinal disorders (FGIDs), which include GI symptoms that necessitate endoscopy, but lack laboratory markers, endoscopic findings, and histologic evidence associated with inflammation (Black et al., 2020; Hyams et al., 2016; McOmber and Shulman, 2008; Santucci et al., 2020). Thus, while FGID is a distinct disease entity that does not represent completely healthy tissue, it is a critical non-inflamed age-matched control cohort with which to contextualize the inflammation observed in pediCD.

The current standard of care for pediCD (as with adult CD) is tailored to the patient’s disease location, clinical behavior and severity, though use of prednisone and other immunomodulators, as well as monoclonal antibodies including anti-TNFα, are common (Hyams et al., 1991; Ruemmele et al., 2014; Turner et al., 2012). While targeting TNF is shared across many autoimmune and inflammatory conditions, it is not successful in all patients, and many go on to develop anti-TNF-refractory disease. It is of tremendous importance for the field to precisely understand and characterize for which patients anti-TNF therapy is not necessary, in which it may succeed in controlling disease, and which will likely be refractory to treatment. Several ideas based on individual immunogenicity and pharmacokinetics have been proposed to explain TNF-refractory disease, including gender (M>F), low albumin levels, high BMI, and high baseline C-Reactive Protein (CRP) (Atreya et al., 2020; Digby-Bell et al., 2020). However, no single identifiable clinical or biochemical biomarker reliably predicts disease response versus resistance to anti-TNF treatment (Stevens et al., 2018).

The primary cellular lineages sampled from intestinal biopsies of CD patients represent both the epithelium and lamina propria, and include epithelial cells, stromal cells, hematopoietic cells and neuronal processes whose cell bodies are present outside of these regions (Buisine et al., 2001; Leeb et al., 2003; Leonard et al., 1995; Lilja et al., 2000; Müller et al., 1998; Souza et al., 1999; Stappenbeck and McGovern, 2017; Takayama et al., 2010). Alterations to all cellular lineages have been implicated in CD (Furey et al., 2019; Martin et al., 2019). Histologically, CD is characterized by a granulomatous inflammation, and by alterations in almost every leukocyte cell type studied, including an increase of cytotoxic lymphocytes, alterations in γδ T cells, increases in mast cells and their production of TNF, activation and shifts in antibody isotypes towards IgM and IgG from B cells and plasma cells, and cytokine production by macrophages (Catalan-Serra et al., 2017; Lilja et al., 2000; Meijer et al., 1979; Mitsialis et al., 2020; Müller et al., 1998; Sieber et al., 1984; Takayama et al., 2010). In the stromal compartment, there is evidence for enhanced vascularization and increased expression of ICAM-1 and MAdCAM-1 by vascular beds, substantial remodeling of collecting lymphatics, and altered migratory potential of fibroblasts (Leeb et al., 2003; Souza et al., 1999). Epithelial barrier dysfunction has also been noted, including alterations to mucus production, microvilli, and Paneth cell dysfunction (Buisine et al., 2001; Stappenbeck and McGovern, 2017). Previous studies have therefore identified that essentially all cell types may be meaningfully altered during CD, highlighting the important need to comprehensively chart the concerted cellular changes that define CD, its severity, and its expected treatment course. Importantly, which changes are predictive of more severe disease, response to treatment, or treatment resistance in pediCD remain largely unknown.

Single-cell RNA-sequencing (scRNA-seq) is enhancing our ability to comprehensively map and resolve the cell types, subsets, and states present during health and disease. Recent work has generated cellular atlases of adult CD and UC from patients who were already on treatment and without linking to clinical follow-up studies (Corridoni et al., 2020b, 2020a; Drokhlyansky et al., 2019; Elmentaite et al., 2020; Huang et al., 2019; Kinchen et al., 2018; Martin et al., 2019; Parikh et al., 2019; Smillie et al., 2019). The potential impact of scRNA-seq on our understanding of IBD is evidenced by studies of adult UC, which have identified potential functional roles for poorly understood colonic cell subsets, such as the BEST4+ enterocyte, and identified pathological alterations in UC biopsies compared to healthy controls, including an expansion of microfold-like cells, *IL13RA2+IL11+* inflammatory fibroblasts, *CD4+CD8+IL17A+* T cells and *CD8+GZMK+* T cells (Smillie et al., 2019). A single-cell study of on-treatment adult CD patients comparing non-inflamed and inflamed tissue from surgically-resected bowel identified IgG+ plasma cells, inflammatory mononuclear phagocytes, activated T cells and stromal cells, comprising the “GIMATS” pathogenic cellular module (Martin et al., 2019). This module was used to derive a gene signature associated with resistance to anti-TNF therapy in a distinct cohort profiled by bulk RNA-seq. Very recent work has profiled how fetal transcription factors are reactivated in Crohn’s disease epithelium, leading to significant decreases in mature absorptive epithelial cells and increases in secretory/goblet cells (Elmentaite et al., 2020). Together, these and other studies have demonstrated the power of scRNA-seq to nominate individual and collective cell states that are associated with disease, and underscored the unmet need to apply these techniques to untreated disease and associate them with disease severity in order to more specifically identify pathognomonic and prognostic cell states.

Most comprehensive scRNA-seq atlases of inflammatory disease conditions include patients being treated with a variety of agents, and for which the biopsies often reflect a partial treatment-refractory state to combinations of antibiotics, corticosteroids, immunomodulators, and biologics including anti-TNF monoclonal antibodies. A treatment-naïve single-cell atlas in an inflammatory disease condition linking observed baseline cell clusters with disease trajectory and treatment outcomes has yet to be reported. In order to address this unmet need in pediCD, we created the prospective PREDICT study (Clinicaltrials.gov #NCT03369353) to identify, profile, and understand pediatric IBD, as well as uninflamed FGID controls. Here, we present detailed diagnostic data from the first cohort of 27 patients enrolled on PREDICT, including 14 pediCD and 13 FGID patients, together with flow cytometric and scRNA-seq studies of the cellular composition of the terminal ileum (**Figure 1**). Furthermore, through prospective annotation of clinical metadata and detailed longitudinal follow-up, we stratify our pediCD cohort by clinically-guided therapeutic decisions separating patients treated with anti-TNF mAbs versus those with biopsy-proven pediCD, but for whom clinical symptoms were sufficiently mild that the treating physician did not prescribe anti-TNF agents (this cohort is termed “Not On Anti-TNF” or “NOA”). Importantly, we were also able to separate the cohort of patients treated with anti-TNF agents into sub-cohorts of those who achieved a full response (FR) to this therapy versus those who achieved only a partial response (PR). Because PREDICT enrolled patients prior to their diagnostic endoscopy, we were able to relate these clinical outcomes to the patients’ cell states at diagnosis, and were further able to integrate matched on-treatment biopsies from these same pediCD patients. We contextualize our findings in pediCD relative to a cohort of FGID patients, which provides an age-matched comparator cohort with clinical GI symptoms, but non-inflammatory disease proven by endoscopy and histologic examination.

**Figure 1.**
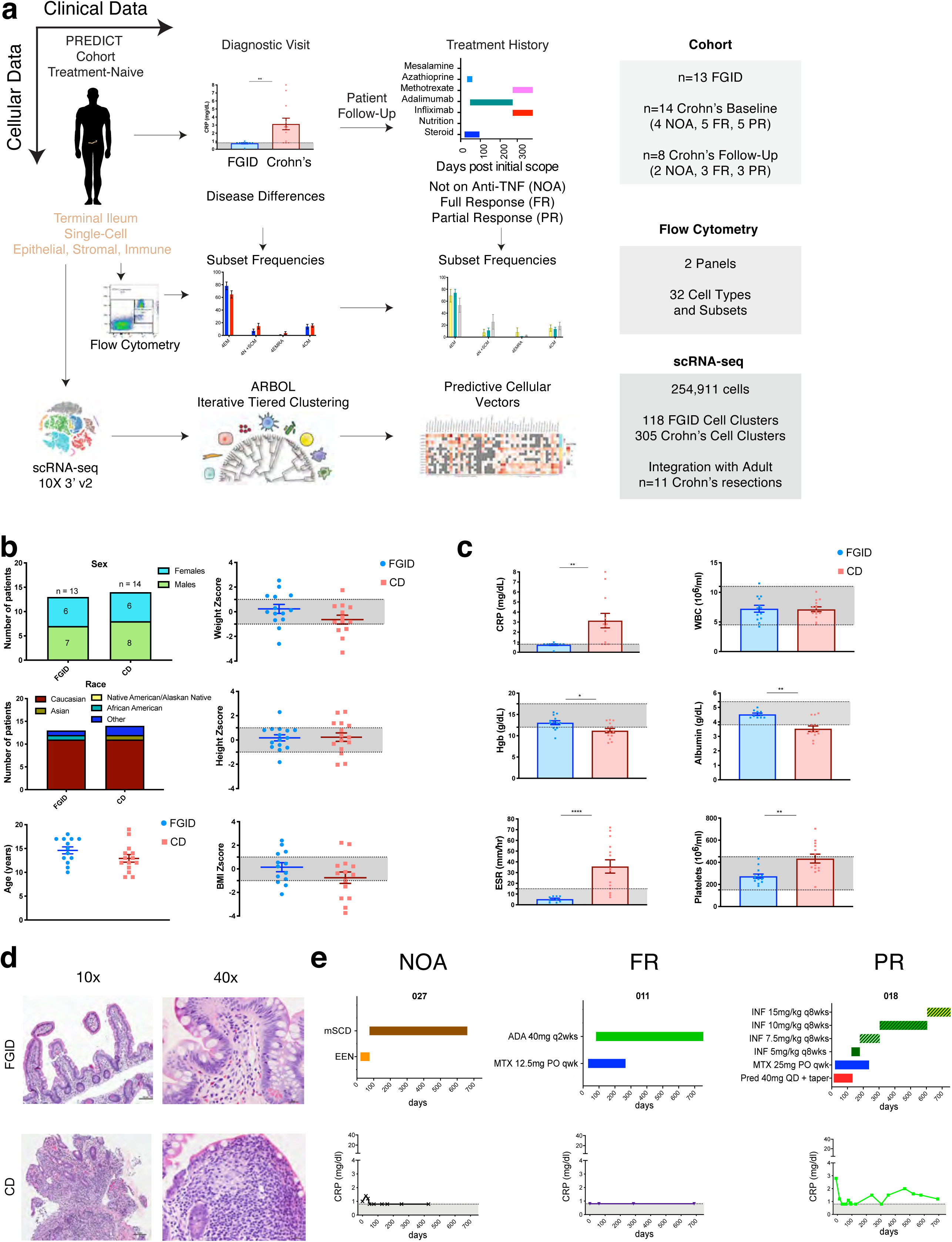
PREDICT Study Design with Patient Diagnostic Criteria and Histopathology. a. Study overview depicting clinical and cellular measurements from 13 functional gastrointestinal disorder (FGID) patients and 14 pediatric Crohn’s disease (pediCD) patients. Terminal ileum biopsies were isolated at a treatment-naïve diagnostic visit, and pediCD patients were followed up to determine their anti-TNF response and categorized as not on anti-TNF (NOA), Full Response (FR), or Partial Response (PR) (see **Methods**). Two panels of flow cytometry allowed for relative frequency quantification of 32 cell types and subsets, and 10X 3’ v2 single-cell RNA-sequencing (scRNA-seq) captured 254,911 total cells including 118 FGID and 305 pediCD end clusters through ARBOL (see **Methods**). b. Demographic data, weight, height, and BMI for cohort (see **Table 1** and **Supplemental Figure 1**); Significance testing by Mann-Whitney U-test. No significance noted in age, weight, height, or BMI z-score for the two cohorts. c. Clinical inflammatory laboratory values for cohort (see **Table 1** and **Supplemental Figure 1**); Significance testing by Mann-Whitney U-test. * indicates p<0.05. ** indicates p <0.005. Significance noted for C-reactive protein (CRP), hemoglobin (Hgb), end sedimentation rate (ESR), albumin, and platelets. No significance noted for WBC. d. Representative histopathology of FGID (top) and pediCD (bottom) at 10x (scale bar = 100um) and 40x (scale bar = 20um) magnification. e. Representative treatment history and clinical inflammatory parameters used for determination of NOA (p027), FR (p011) and PR (p018) status (see **Methods**, **Table 1**, and **Supplemental Figure 1;** ADA: adalimumab, INF: infliximab; MTX: methotrexate; Pred: prednisone; mSCD: modified specific carbohydrate diet; EEN: exclusive enteral nutrition).

**Table 1.** Demographics and clinical characteristics of patients analyzed on PREDICT. *indicates significance detected between Crohn’s Disease (CD) and Functional Gastrointestinal Disorder (FGID).

|  |  | **Crohn's Disease** | **FGID** | **P-values** |
| --- | --- | --- | --- | --- |
|  |  | n = 14 | n = 13 |  |
| **Demographics** |  |  |  |  |
|  | Age at diagnosis (year) | 12.5 (9-19) | 16 (10-18) | 0.096 |
|  | Female sex | 64% (9 patients) | 46% (6 patients) | >0.99 |
|  | Caucasian | 79% (11 patients) | 84% (11 patients) |  |
|  | African American | 0% (0 patients) | 8% (1 patient) |  |
|  | Native American or Alaskan Native | 0% (0 patients) | 0% (0 patients) |  |
|  | Asian | 7% (1 patient) | 0% (0 patients) |  |
|  | Mixed race | 14% (2 patients) | 0% (0 patients) |  |
|  | Other | 0% (0 patients) | 8% (1 patient) |  |
| **Growth** |  |  |  |  |
|  | Weight Z score | -0.61<br>(-3.31 to 2.31) | 0.12<br>(-1.34 to 2.54) | 0.12 |
|  | Height Z score | 0.09<br>(-2.04 to 2.32) | 0.01<br>(-1.82 to 2.3) | 0.86 |
|  | BMI Z score | -0.44<br>( -3.74 to 2.1) | 0.2<br>( -2.15 to 2.4) | 0.23 |
| **Disease location/activity** |  |  |  |  |
|  | Location |  |  |  |
|  | L1 | 0% (0 patients) |  |  |
|  | L2 | 0% (0 patients) |  |  |
|  | L3 | 93% (13 patient) |  |  |
|  | P modifier | 14% (2 patients) |  |  |
|  | Behavior |  |  |  |
|  | B1 | 93% (13 patient) |  |  |
|  | B2 | 0% (0 patients) |  |  |
|  | B3 | 0% (0 patients) |  |  |
|  | Severity in wPCDAI |  |  |  |
| **Laboratory evaluation** |  |  |  |  |
|  | CRP (mg/dl) | 3.9 ± 0.8<br>(0.8 to 8) | 0.7 ± 0.1<br>(0.1 to 1) | 0.0019* |
|  | ESR (mm/hr) | 35.8 ± 6.2<br>(7 to 69) | 5.2 ± 0.9<br>(2 to 8) | <0.0001* |
|  | Hgb (g/dl) | 11.2 ± 0.5 | 13.1 ± 0.5 | 0.0018* |
|  |  | (8.3 to 13.7) | (9.4 to 15.5) |  |
|  | PLT (k/mm<sup>3</sup>) | 424.4 ± 38.8<br>(176 to 704) | 274.2 ± 19.3<br>(186 to 433) | 0.0213* |
|  | WBC (k/mm<sup>3</sup>) | 7.1 ± 0.4<br>(4.6 to 10.1) | 7.1 ± 0.6<br>(4.2 to 11.5) | 0.914 |
|  | Albumin (g/dl) | 3.5 ± 0.2<br>(2.5 to 4.6) | 4.5 ± 0.1<br>(4.3 to 5) | 0.0023* |

Several analytical approaches have been developed to enable the generation and interrogation of clusters during the curation of single-cell transcriptomic atlases (Hie et al., 2020). One such method, sub-clustering of broad clusters, has proven to be a powerful tool for isolating highly specific axes of variation that are obscured by analyses whose principal axes of variation are broad cell types (Bakken et al., 2021; La Manno et al., 2021; Sikkema et al., 2022; Tasic et al., 2018; Zeisel et al., 2018). However, while sub-clustering analysis is a powerful tool allowing access to the hierarchy of cell states, this method is manually intensive and there is little consensus or standardization in clustering parameters and annotation methods. To address this issue, we designed a principled, modular, automated, iterative sub-clustering routine made possible by application of parameter scanning methods (Rousseeuw, 1987; Shekhar et al., 2016). We developed this tool, ‘ARBOL’ (named after tree in Spanish), of which iterative tiered clustering (ITC) is a key component, in R and python, integrating with Seurat and Scanpy functions, to make it accessible and easily incorporated into common workflows, and have curated a GitHub repository with illustrative vignettes. Here, we use ARBOL to standardize fine-grained cell state discovery by the creation and cultivation of a tree of cell states, followed by the automated generation of cell names to aid in the annotation of end clusters by unique and descriptive genes, inclusive of a method for identifying clusters containing high or low patient diversity, and for pruning of the resultant tree.

In order to respect the distinct clinical entities of FGID and pediCD, and the co-variation structure that can define rare but important cell subsets and states, we generate two fully-annotated cellular atlases for these pediatric GI diseases. These atlases consist of 94,451 cells for FGID and 107,432 for pediCD. We provide key gene-list resources for further studies, and nominate a vector of lymphoid, myeloid and epithelial cell states that anticipate disease severity and treatment outcomes. We term this vector ‘pediCD T cell, innate lymphocyte, myeloid and epithelial’ (pediCD-TIME). This cellular vector correlates strongly with both the clinical presentation of pediCD severity, and with the distinction between anti-TNF full-or partial-response. The significant changes in cell composition we have discovered to be associated with disease severity are increases of proliferating T cells, cytotoxic NK cells, subsets of monocytes/macrophages, and plasmacytoid dendritic cells (pDCs), accompanied by decreases of metabolically-specialized epithelial cell subsets. We successfully validate this vector in two bulk RNA-seq treatment-naïve IBD cohorts. We contextualize our treatment-naïve pediCD atlas by joining it with the FGID atlas through label-retaining (manual clustering; Random Forest; ARBOL) and label-free (Hellinger distance on UMAP) methods. Finally, we integrate our pediCD atlas with follow-up post-treatment biopsies from our cohort, and with the adult on-treatment CD patients of the “GIMATS” scRNA-seq atlas. Taken together, we identify that anti-TNF treatment over time pushes the overall pediCD cellular ecosystem towards the more treatment-refractory disease state found in adult CD.

## Results

### Study cohort clinical outcomes

The PREDICT study prospectively enrolled treatment-naïve, previously undiagnosed pediatric patients with GI symptoms necessitating diagnostic endoscopy. The current analysis focuses on patients enrolled in the first year of the study, during which time 14 patients with pediCD and 13 patients with FGID were enrolled and had adequate ileal samples for single-cell analysis (**Figure 1; Supplemental Figure 1**). After their initial diagnosis, patients with pediCD were followed clinically for up to 3 years (with the determination of anti-TNF response made at 2 years). Patients with FGID were followed clinically only as needed. The median time from diagnosis for the pediCD and FGID cohorts to the time of database lock was 32.5 and 31 months, respectively. Of the pediCD and FGID patients analyzed, the median age at diagnosis was 12.5 years and 16 years respectively (p = 0.095; Mann-Whitney U-test), with no significant differences in gender (**Figure 1b**; **Table 1**). Patient weight, height, and BMI z-scores were not significantly different between pediCD and FGID (**Figure 1b**; **Table 1**); however, in addition to the diagnostic differences on histologic analysis, several key clinical laboratory values, including C-reactive protein (CRP), Erythrocyte Sedimentation Rate (ESR), hemoglobin concentration, albumin concentration, and platelet count were significantly different between pediCD and FGID (**Figure 1c,d**; **Table 1**).

### Treatment with anti-TNF agents and response to therapy

Patients with pediCD were initially divided into two cohorts. Those with milder disease characteristics (n = 4) as determined by their treating physician, were not treated with anti-TNF therapies, and are noted as ‘NOA’. For patients with more severe disease (n = 10), anti-TNF therapy (with either infliximab or adalimumab, **Table 2**) was initiated within 90 days of diagnostic endoscopy. All pediCD patients were followed prospectively and categorized as FR (n = 5) or PR (n = 5) to anti-TNF therapy based on the following criteria: FR was defined as clinical symptom control and biochemical response (measuring CRP, ESR, albumin, and complete blood counts (CBC)), and with a weighted Pediatric Crohn’s Disease Activity Index (PCDAI) score of <12.5 on maintenance anti-TNF therapy with no dose adjustments required (Cappello and Morreale, 2016; Hyams et al., 1991; Sandborn, 2014; Turner et al., 2017, 2012). PR to anti-TNF therapy was defined as a lack of full clinical symptom control as determined by the treating physician or lack of full biochemical response, with documented escalation of anti-TNF therapy or addition of other agents (**Figure 1e**; NB: patients in our cohort were dose-escalated because of clinical symptoms). Medication timelines and clinical laboratory data through 2 years post-diagnosis for all pediCD patients is shown in **Supplemental Figure 1.** The designation of FR or PR was made at 2 years of follow-up for all pediCD patients. During our study window, 8 pediCD (2 NOA, 3 FR, 3 PR) received repeat biopsies which we also analyzed through scRNA-seq (see below).

**Table 2.** Demographics and clinical characteristics of Crohn’s Disease cohorts. *indicates significance detected between.

|  |  | Partial Responder | Full Responder | Not on anti-TNF |
| --- | --- | --- | --- | --- |
|  |  | n = 5 | n = 5 | n = 4 |
| **Demographics and follow-up** |  |  |  |  |
|  | Age at diagnosis (year) | 12 (10 to 14) | 12 (9 to 14) | 15.5 (9-19) |
|  | Female sex | 40% (2 patients) | 40% (2 patients) | 50% (2 patients) |
|  | Caucasian | 80% (4 patients) | 80% (4 patients) | 50% (2 patients) |
|  | African American | 0% (0 patient) | 0% (0 patient) | 0% (0 patient) |
|  | Native American or Alaskan Native | 0% (0 patient) | 0% (0 patient) | 0% (0 patient) |
|  | Asian | 0% (0 patient) | 0% (0 patient) | 0% (0 patient) |
|  | Mixed race | 20% (1 patient) | 0% (0 patient) | 25% (1 patient) |
|  | Other | 0% (0 patient) | 20% (1 patient) | 0% (0 patient) |
| **Disease location/activity** |  |  |  |  |
|  | Location |  |  |  |
|  | L1 | 0% (0 patient) | 0% (0 patient) | 0% (0 patient) |
|  | L2 | 0% (0 patient) | 0% (0 patient) | 0% (0 patient) |
|  | L3 | 100% (5 patients) | 100% (5 patients) | 75% (3 patients) |
|  | P modifier | 20% (1 patient) | 0% (0 patients) | 25% (1 patient) |
|  | Behavior |  |  |  |
|  | B1 | 100% (5 patients) | 100% (5 patients) | 75% (3 patients) |
|  | B2 | 0% (0 patients) | 0% (0 patients) | 0% (0 patients) |
|  | B3 | 0% (0 patients) | 0% (0 patients) | 0% (0 patients) |
|  | Severity in wPCDAI | 65<br>(35 to 77.5) | 65<br>(22.5 to 87.5) | 26.3<br>(15 to 57.5) |
| **Medications** |  |  |  |  |
|  | Mesalamine | 0% (0 patient) | 20% (1 patients) | 50% (2 patients) |
|  | Azathioprine | 20% (1 patient) | 20% (1 patient) | 25% (1 patient) |
|  | Methotrexate | 80% (4 patient) | 60% (3 patient) | 0% (0 patient) |
|  | Anti - TNF | 100% (5 patients) | 100% (5 patients) | 0% (0 patient) |
|  | Nutritional therapy | 20% (1 patient) | 20% (1 patient) | 25% (1 patient) |
|  | Steroids | 60% (3 patients) | 60% (3 patients) | 50% (2 patients) |
| **Median time since diagnosis** |  |  |  |  |
|  | Median | 31.7<br>(29.6 to 37.2) | 36.3<br>(29.6 to 37.2) | 32<br>(24.2 to 33.7) |

### Flow cytometry of the terminal ileum reveals minimal changes in leukocyte subsets in FGID vs. pediCD, and no significant differences across the pediCD spectrum

We collected terminal ileum biopsies from 14 pediCD patients and 13 uninflamed FGID patients, and prepared single-cell suspensions for flow cytometry and scRNA-seq. Biopsies from pediCD were from actively-inflamed areas adjacent to ulcerations. Biopsies from FGID were from non-inflamed terminal ileum. The epithelium was first separated from the lamina propria before enzymatic dissociation, and flow cytometric analysis was performed on the viable single-cell fraction, which recovered predominantly hematopoietic cells with some remnant epithelial cells (<20% of all cells), likely representing those in deeper crypt regions (**Figure 2; Supplemental Figure 2**). We utilized two flow cytometry panels, allowing us to resolve the principal lymphoid (CD4 or CD8 T cells, NK cells, B cells, innate lymphoid cells, γδ T cells, CD8αα+ IELs, pDCs) and myeloid (monocytes, granulocytes, HLA-DR+ mononuclear phagocyte) cell subsets (**Supplemental Figure 2, Supplemental Table 1**). From these panels, which generated 32 gates identifying cell lineages, types and subsets, only HLA-DR+ macrophages/DCs and pDCs were significantly increased in pediCD relative to FGID (**Figure 2d**). We also analyzed within pediCD, comparing the baseline samples of 4 NOA, 5 FR and 5 PR patients, and noted no significant differences between NOA and patients on anti-TNF, or between FRs and PRs to anti-TNF (**Figure 2e**). Together, this suggests that despite the substantial endoscopic, histologic, and clinical parameters that distinguish FGID and pediCD (**Figure 1**), the broad single-cell type composition of the terminal ileum appears minimally altered in pediCD, save for an increased frequency of pDCs and HLA-DR+ mononuclear phagocytes.

**Figure 2.**
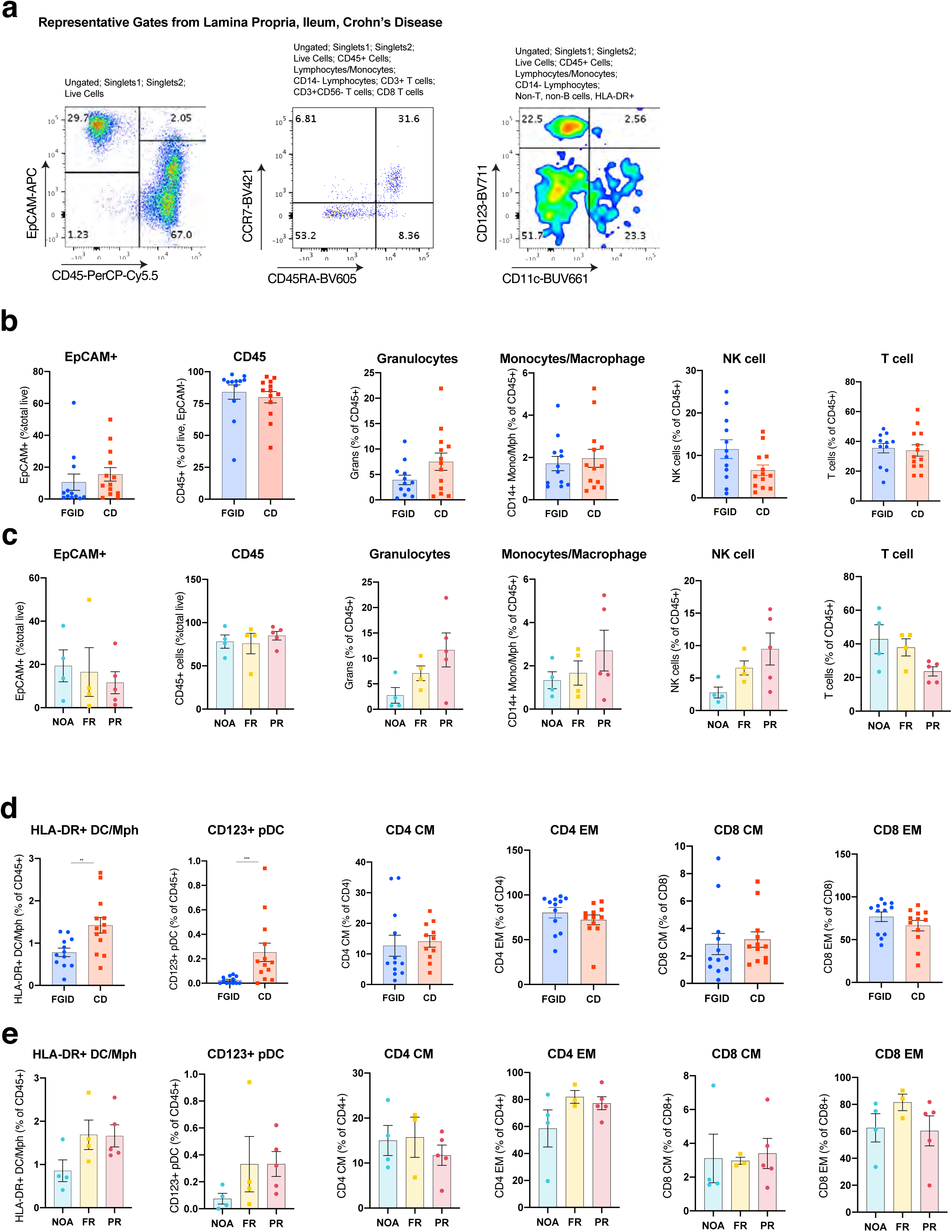
Flow Cytometry of Ileal Biopsies Does Not Reveal Significant Changes in Cell Composition in FGID vs. pediCD or across the pediCD Treatment Response Spectrum. a. Representative flow cytometry end gates for selected cell subsets (left: epithelial and hematopoietic; middle: naïve and effector T cells; right: pDCs and antigen-presenting cells) from single-cell dissociated samples from one terminal ileum biopsy for pediCD patients (see **Supplemental Figure 2** for full gating strategy; numbers represent percentage of cells in each gate). b. Fractional composition of selected cell subsets of CD45+ cells from 13 FGID and 14 pediCD patients (error bars are s.e.m). c. Fractional composition of selected cell subsets of CD45+ cells from 4 NOA, 5 FR and 5 PR patients. d. Fractional composition of dendritic, pDC, central memory (CM) and effector memory (EM) CD4+ and CD8+ cells from 13 FGID vs 14 pediCD patients. Dendritic cells and pDC plotted as percentage of CD45+ cells. CM/EM CD4+ and CD8+ cells plotted as percentage of total CD4+ and CD8+ cells, respectively. p < 0.05 by Mann-Whitney for pediCD versus FGID and 1-way ANOVA for pediCD cohorts). e. Fractional composition of dendritic cells, pDCs, central memory (CM) and effector memory (EM) CD4+ and CD8+ cells from 4 NOA, 5FR, and 5 PR patients. Graphs plotted as in **d**.

### Traditional clustering of scRNA-seq data from FGID and pediCD patients

In addition to flow cytometry, we performed droplet-based scRNA-seq on cell suspensions from the 14 pediCD and 13 FGID patients using the 10X Genomics 3’ V2 platform (**Figure 1**). The analyzed cell suspensions were derived from lamina propria preparations, which our flow cytometry data suggested would be composed primarily of CD45+ leukocytes, alongside a fraction of epithelial cells and stromal/vascular cells. Deconstructing these tissues into their component cells provided us with the ability to identify some of the corresponding cell types (e.g. T or B cell) and subsets (CD8αα+ IEL or CD4+ T cell) to those we identified by flow cytometry. Importantly, it also enabled us to: 1. characterize these major cell types and subsets without needing to pre-select markers, and 2. gain substantially enhanced resolution into the cell states (i.e. gene expression programs determined by co-expression of genes) within these cell types and subsets.

Following library preparation and sequencing, we derived a unified cells-by-genes expression matrix from the 27 samples, containing digital gene expression values for all cells passing quality thresholds (n=254,911 cells; **Supplemental Figure 3; Supplemental Tables 2 and 3; Methods**). We then performed dimensionality reduction and graph-based clustering, noting that despite no computational integration methods being used, FGID and pediCD were highly similar to each other when clustered together and visualized on a uniform manifold approximation and projection (UMAP) plot (**Supplemental Figure 4a-c**). We recovered the following cell types from both patient groups: epithelial cells, T cells, B cells, plasma cells, glial cells, endothelial cells, myeloid cells, mast cells, fibroblasts, and a proliferating cell cluster. We noted that the fractional composition amongst all cells of T cells, B cells, and myeloid cells was not significantly different between FGID and pediCD, similar to the flow cytometric data, and this was also the case for endothelial, fibroblasts, glial, mast and plasma cells, which were not measured through flow cytometry (**Supplemental Figure 4d**). This provided validation and extension of our flow cytometry data documenting that the broad cell type composition of FGID and pediCD is not significantly altered, despite highly-distinct endoscopic and histologic diseases. Based on this joint clustering and annotation of top-level cell types, we then performed differential expression testing identifying significant up- and down-regulated genes across cell types (**Supplemental Figure 4e; Supplemental Table 4**). Within myeloid cells we identified some of the most significantly upregulated genes in pediCD versus FGID to be *CXCL9* and *CXCL10*, canonical IFNγ-stimulated genes, and *S100A8* and *S100A9* which form the biomarker fecal calprotectin (**Supplemental Figure 4f**)(Leach et al., 2007; Ziegler et al., 2021). Within epithelial cells we identified that *APOA1* and *APOA4* were amongst the most significantly downregulated genes, and correspondingly *REG1B*, *SPINK4* and *REG4A* were amongst the most significantly upregulated indicating tradeoffs between lipid metabolism and host defense in pediCD versus FGID (**Supplemental Figure 4f**) (Haberman et al., 2014). In T cells, we noted that the cytotoxic genes *GNLY* and *GZMA* were amongst the most significantly upregulated in pediCD, with almost no genes downregulated (**Supplemental Figure 4f**).

We then systematically re-clustered each broad cell type, identifying increasing cellular heterogeneity. Given that we detected changes in the frequency of HLA-DR+ macrophages/dendritic cells and pDCs between pediCD and FGID by flow cytometry, we initially focused on the myeloid cell type sub-clustering, containing dendritic cells, macrophages, monocytes, and pDCs (**Supplemental Figure 4g)**. Working within this analysis paradigm revealed that a traditional clustering approach had difficulty identifying the boundaries of clusters, and whether a cluster composed primarily of pediCD rather than FGID cells represented a unique cell subset, or a cell state overlaid onto a core cell subset gene expression program (**Supplemental Figure 4g, Methods**). These initial findings also raised the possibility that subjective analytical decisions in this manual joint clustering approach might obscure some of the unique biology of FGID and pediCD. For instance, this analytical approach could lead to hybrid clusters, informed by cells from both FGID and pediCD, that according to recent Human Cell Atlas-scale efforts in the lung, may limit discovery of rare but critical disease-specific cell types, subsets, or states (Sikkema et al., 2022).

### ARBOL automated iterative tiered clustering

In order to approach the analytic challenge from a more principled direction, we made four key changes to our workflow. First, as FGID represents a non-inflamed condition and pediCD and inflamed state, we proceeded to analyze FGID and pediCD samples separately to define corresponding cell type, subset, and state clusters and markers in order to respect the underlying disease biology and maximize the potential for high-quality annotation of cell subsets. Second, to minimize analyst-to-analyst variability in the choice of clustering parameters, which can significantly under- or over-cluster data, we developed ARBOL (https://github.com/jo-m-lab/ARBOL). ARBOL is an automated clustering approach—built with conventional Seurat functions—to maximize the silhouette score at each tier of iterative sub-clustering and stop sub-clustering when a specific granularity is reached (**Supplemental Figure 5a,b; Methods**), Third, we systematically generated descriptive names for cell types and subsets together with differentiating marker genes. Fourth, we accounted for the number and diversity of patients which compose each cluster using Simpson’s Index of Diversity. This enabled us to focus our study on clusters that, despite being rare, were reproducibly found in the majority of patients (Simpson, 1949). Using ARBOL, each tier of analysis is typically under-clustered relative to traditional empirical analyses, but the automation proceeds through several more tiers (typically 4 to 5) until stop conditions (e.g. cell numbers and differentially expressed genes; **Methods**) are met. Intriguingly, this number of tiers was also manually selected in the HCA expert-annotated lung cell atlas, where reliable and robust cell clusters were found in the 0.05% cell frequency range (Sikkema et al., 2022).

We then inspected all outputs (182 FGID and 425 pediCD clusters) and provided descriptive cell cluster names independently for FGID and pediCD (**Supplemental Figures 5b and 6**). We also focused at this stage on flagging putative doublet clusters or clusters where the majority of differentially expressed genes which triggered further clustering consisted of known technical confounders in scRNA-seq data (e.g. mitochondrial, ribosomal, and spillover genes from cells with high secretory capacity) yielding a final number of 118 FGID and 305 pediCD clusters (**Supplemental Figure 5b**) (Smillie et al., 2019). Importantly, this quality-control step could often be missed by traditional scRNA-seq analysis, since these analysis paradigms retain such cells within higher-tier clusters, yet their inclusion of these cells would confound both differential composition and expression analyses if not identified. We note this clustering method represents a data-driven approach, though it may not always reflect a cellular program or transcriptional module of known biological significance or stability. Nevertheless, we highlight that many of these small clusters (∼100 cells; 0.1% of total cells) which may represent transient cell states, are found across all FGID or pediCD patients surveyed, thus suggesting a level of consistency across diseases, and across genetically and environmentally unrelated individuals.

### Two comprehensive atlases of FGID and pediCD

To generate the final dendrograms for the FGID and pediCD cell atlases, we hierarchically clustered all end cell states, and performed 1 vs. rest within-Tier 1 clusters (i.e. broad cell types) differential expression to provide systematic names for cells. The naming heuristic was based on the cell type classification, cell subset classification with literature-based markers, and two genes using a rank-score function (**Figures 3 and 4; Supplemental Figure 6; Methods**). We provide complete gene lists at three levels of comparison: cell types (1 vs. rest across all cells), subsets (1 v. rest across all cell types), and states (1 vs. rest within-Tier 1 cell-type) in **Supplemental Tables 5-10**. More specifically, to select marker genes for naming in a data driven manner, we used 1 vs. rest within-cell-type differential expression (**Supplemental Tables 7 and 10**; Wilcoxon, Bonferroni adjusted p<0.05), and a rank-score function (-log(sig+1) * avg_logFC * (pct.in / pct.out)) to prioritize the top 2 marker genes. For example, using this ranking system, we identified *CCL3* and *CD160* as two genes significantly enriched in one NK cluster (adj. p-value = 0, expression within-cluster >40% cells positive and in other Tier 1 T cells <6%). This resulted in a final name for this cluster of CD.NK.CCL3.CD160. We repeated this process for all FGID and pediCD cells within Tier 1 B cell, endothelial, epithelial, fibroblast, plasma cell, myeloid cell, mast cell, and T cell identified clusters, and provide systematically generated names for all (**Supplemental Figure 5**; **Supplemental Tables 11 and 12**).

**Figure 3.**
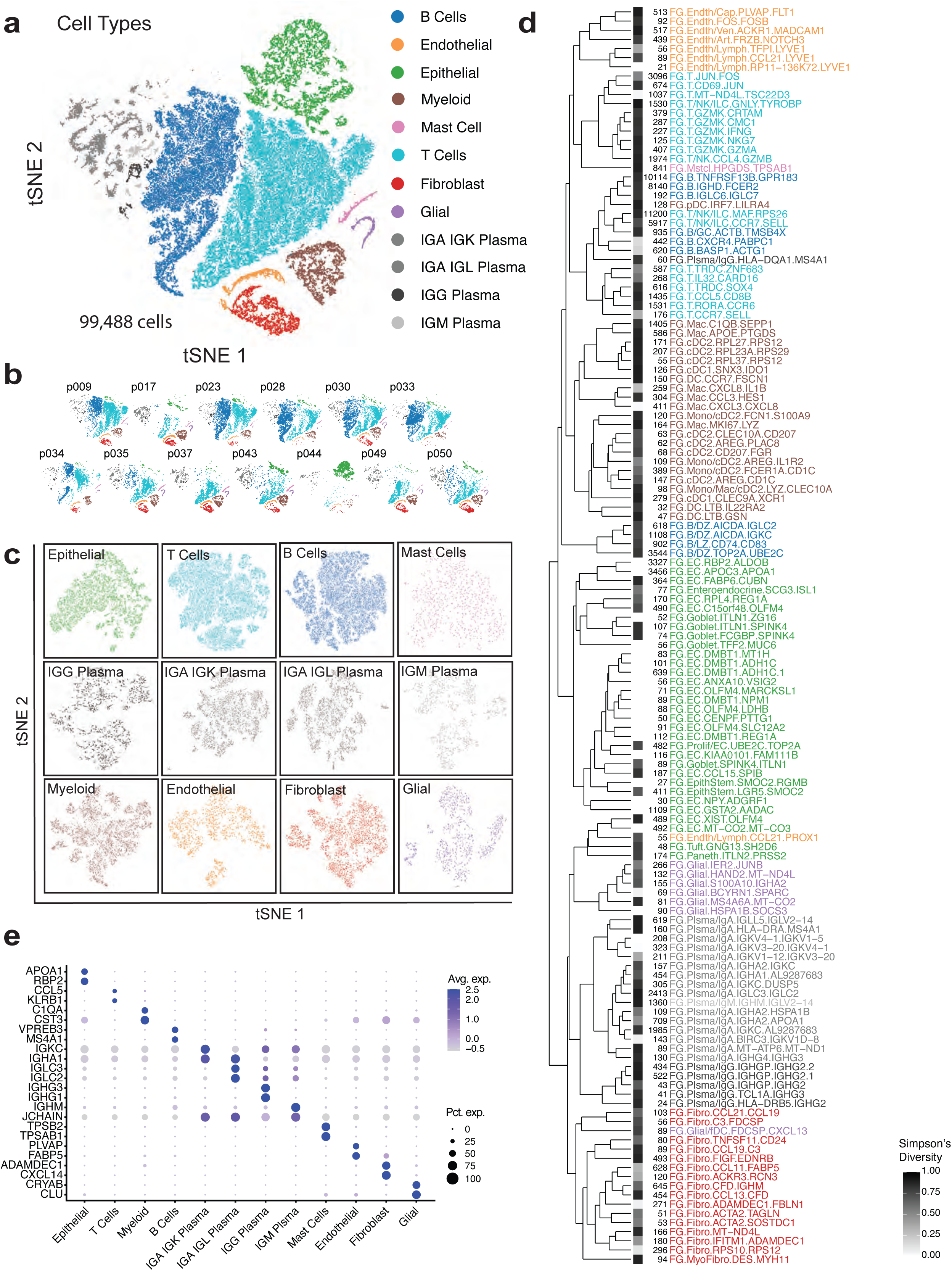
A Comprehensive Cell Atlas of Terminal Ileum in Non-Inflammatory FGID. a. tSNE of 99,488 single-cells isolated from terminal ileal biopsies of 13 FGID patients. Colors represent major cell type groups determined via Louvain clustering with resolution set by optimized silhouette score. b. tSNE as in a with individual patients plotted. For specific proportions please see Supplemental Figure 4. c. tSNE of each major cell type which was used as input into iterative tiered clustering (ITC). d. End clusters determined by ARBOL of complete FGID data set were hierarchically clustered on the median expression of 4,445 pairwise differentially expressed genes, using complete linkage and distance calculated with Pearson correlation, between each end cell cluster. Simpson’s Index of Diversity represented as 1-Simpson’s where 1 (black) indicates equivalent richness of all patients in that cluster, and 0 (white) indicates a completely patient-specific subset. Numbers represent the number of cells in that cluster. Names of subsets are determined by Disease.CellType.GeneA.GeneB as in **Methods**. e. Dot plot of 2 defining genes for each cell type. Dot size represents fraction of cells expressing the gene, and color intensity represents binned count-based expression level (log(scaled UMI+1)) amongst expressing cells. All cluster defining genes are provided in **Supplemental Tables 5 to 7**.

**Figure 4.**
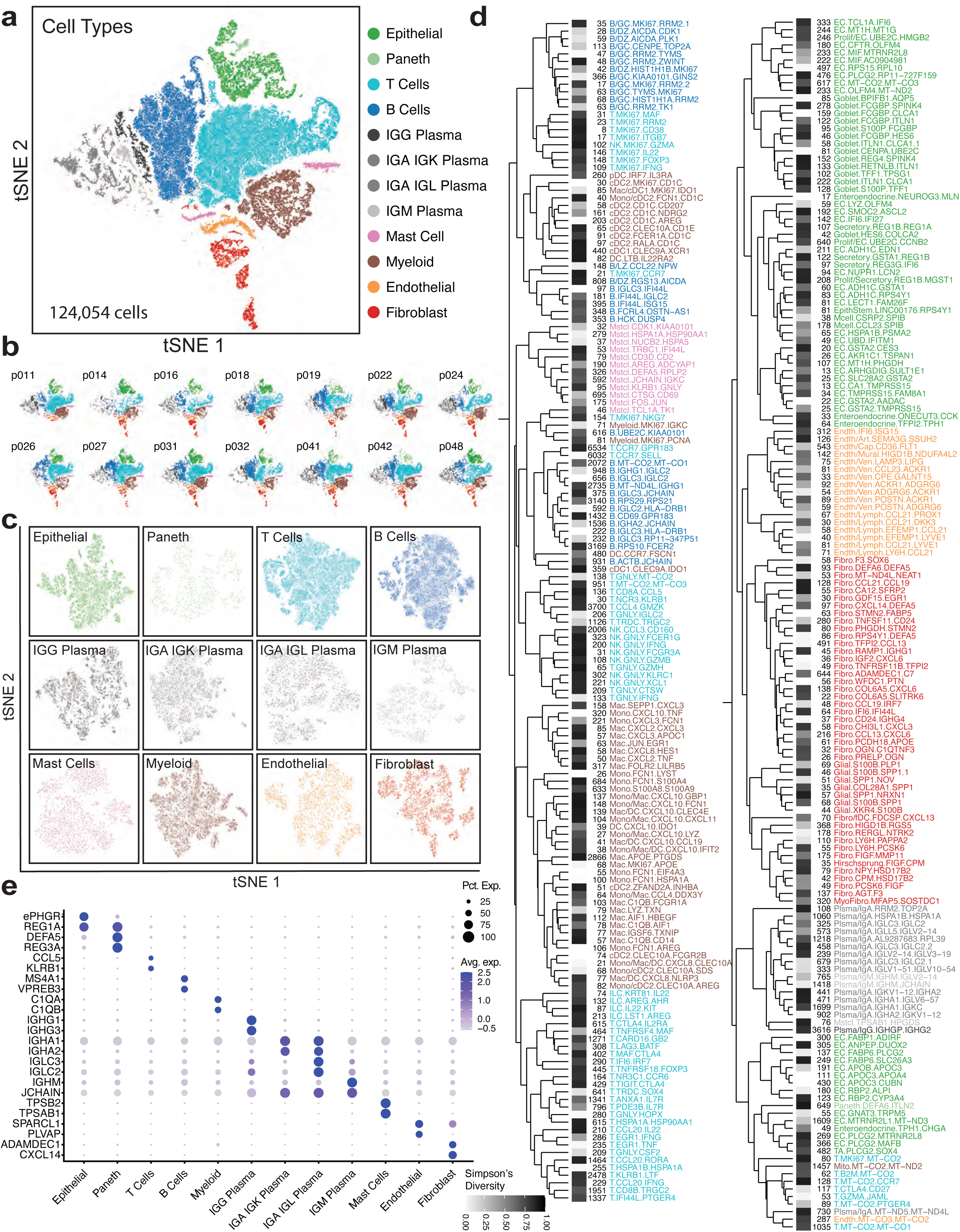
A Comprehensive Cell Atlas of Terminal Ileum in pediCD. a. tSNE of 124,054 single-cells isolated from terminal ileal biopsies of 14 pediCD patients. Colors represent major cell type groups determined via Louvain clustering with resolution set by optimized silhouette score. b. tSNE as in a with individual patients plotted. For specific proportions please see **Supplemental Figure 4**. c. tSNE of each major cell type which was used as input into iterative tiered clustering (ITC). d. End clusters determined by ARBOL of complete pediCD data set were hierarchically clustered on the median expression of 1,844 pairwise differentially expressed genes, using complete linkage and distance calculated with Pearson correlation, between each end cell cluster. Simpson’s Index of Diversity represented as 1-Simpson’s where 1 (black) indicates equivalent richness of all patients in that cluster, and 0 (white) indicates a completely patient-specific subset. Numbers represent the number of cells in that cluster. Names of subsets are determined by Disease.CellType.GeneA.GeneB as in **Methods**. e. Dot plot of 2 defining genes for each cell type. Dot size represents fraction of cells expressing the gene, and color intensity represents binned count-based expression level (log(scaled UMI+1)) amongst expressing cells. All cluster defining genes are provided in **Supplemental Tables 8 to 10**.

From the 99,488 cells profiled from 13 FGID patients, we recovered 12 Tier 1 clusters, representing the main cell types found in the lamina propria and remnant epithelium contained in an ileal biopsy, which we display on a t-stochastic neighbor embedding (t-SNE) plot colored by cluster identity (**Figure 3a; Supplemental Figure 5b**). From the 124,054 cells profiled from 14 pediCD patients, we recovered 12 Tier 1 clusters which we also display on a t-SNE plot colored by cluster identity (**Figure 4a; Supplemental Figure 5b**). Distinct from FGID, Paneth cells clustered separately at Tier 1 in pediCD, while glial cells were now found within the fibroblast Tier 1 cluster. Inspecting each individual patient’s contribution to each broad cell type, we noted that all patients contributed to all Tier 1 clusters in both FGID and pediCD (FGID: **Figure 3b**; pediCD: **Figure 4b; Supplemental Figure 4c,d; Methods Supplemental Table 13**). As patient identity did not factor into ARBOL stop conditions, we then calculated Simpson’s Index of Diversity for each of the end clusters (**Figure 3d; Supplemental Figure 5**). Although low diversity clusters may still reflect important biology for individual patients, we comment more extensively on clusters with high patient diversity. In pediCD, most end clusters are conserved across multiple patients, while only 16/305 clusters were single-patient clusters (**Figure 4d; Supplemental Figure 5**). We note that in pediCD, relative to FGID, a higher fraction of cell clusters exhibited lower patient diversity. In the **Methods** section, we provide full explanations for how names for subsets and clusters were defined for B cells, myeloid cells, T/NK/ILC’s, epithelial cells, endothelial cells, fibroblasts, mast cells and plasma cells, and include links to corresponding **Supplemental Tables** containing full supporting gene lists and data visualization. We share portal links in **Data Availability**.

Using this analytical workflow, we present two comprehensive cellular atlases, separately mapping FGID (**Figure 3**) and pediCD (**Figure 4**). The pediCD atlas enabled us to nominate cell states associated with disease severity and treatment outcomes in this disease (**Figures 5 and 6**), identify histological relationships between epithelial/myeloid/lymphoid cells (**Figure 7**), and understand how anti-TNF pushes the pediatric treatment-naïve ecosystem towards an adult tissue state (**Figure 8**).

**Figure 5.**
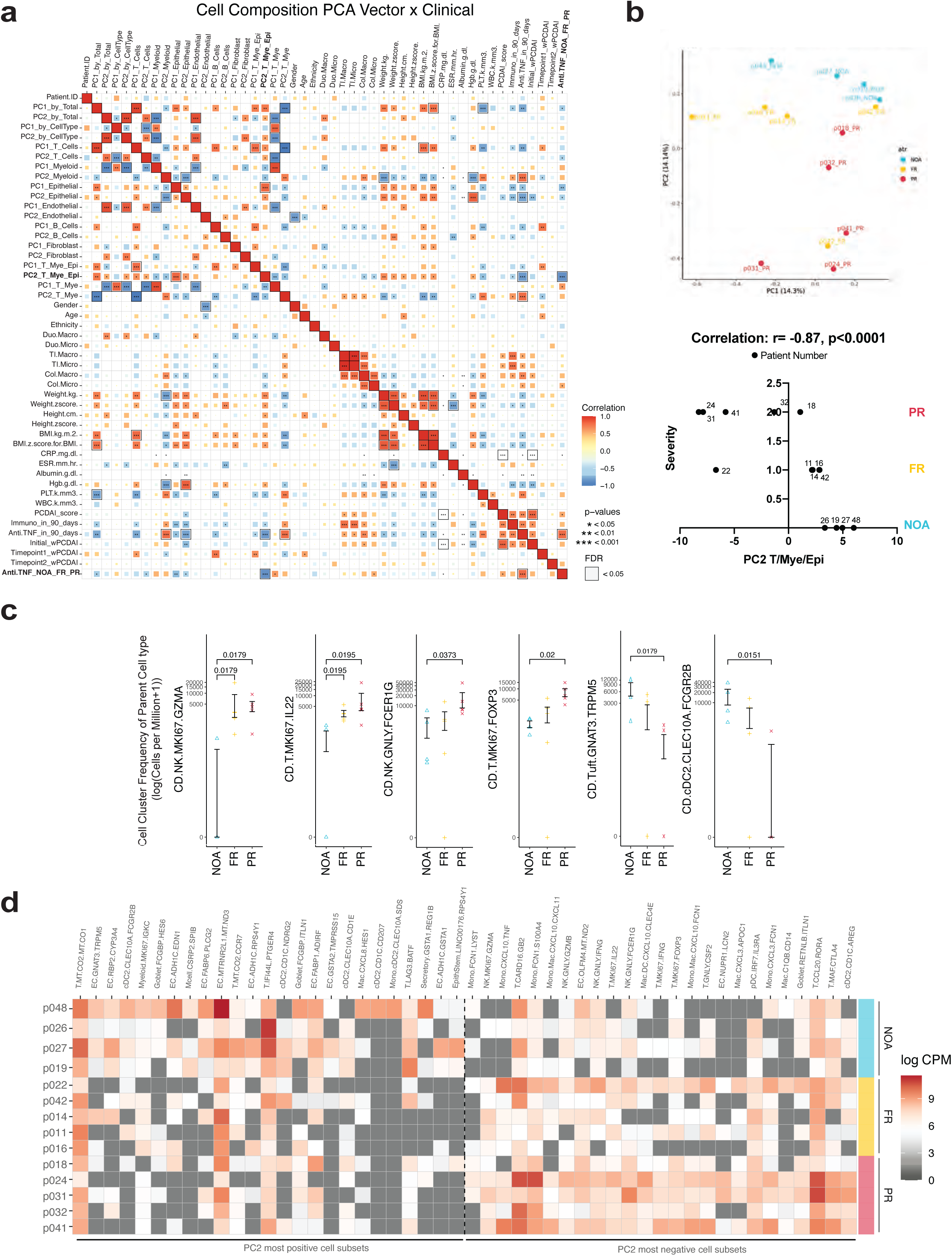
A Collective Cell Vector in pediCD Reveals Predictive Axes of Disease Trajectory and Treatment Response. a. Spearman rank correlation heatmap of principal components calculated from the frequencies of each end cluster per main cell type together with clinical metadata. Correlation is represented by both the intensity and size of the box and those which are FDR < 0.05 have a bounding box. b. A PCA plot of each patient’s end cell cluster composition as determined by ARBOL for T/NK/ILC, from the pediCD dataset where the end cell clusters frequency is calculated amongst all cell types and colors represent anti-TNF response. (inset highlights the specific correlation between PC2 of the T, Myeloid, Epithelial cell frequency analysis with anti-TNF response). We term this principal component “pediCD-TIME”. c. Cell cluster frequencies of the parent cell type found to be significant by Mann-Whitney U test between selected clusters (see **Supplemental** Figure 7 for all graphs; **Supplemental Table 15**). d. Heatmap showing cell frequencies per patient of most positive and negative cell subsets of PC2 from PCA performed on T/NK/ILC, myeloid and epithelial cell subsets (**Supplemental Table 16**). Cell subsets are sorted by PC2 score, and patients were sorted by anti-TNF response. Heatmap is not normalized and displaying the log counts-per million of each cell subset normalized per cell type. *Patient p022’s response category changed from FR to PR after database lock in December of 2020. No other patient’s categorization has changed.

**Figure 6.**
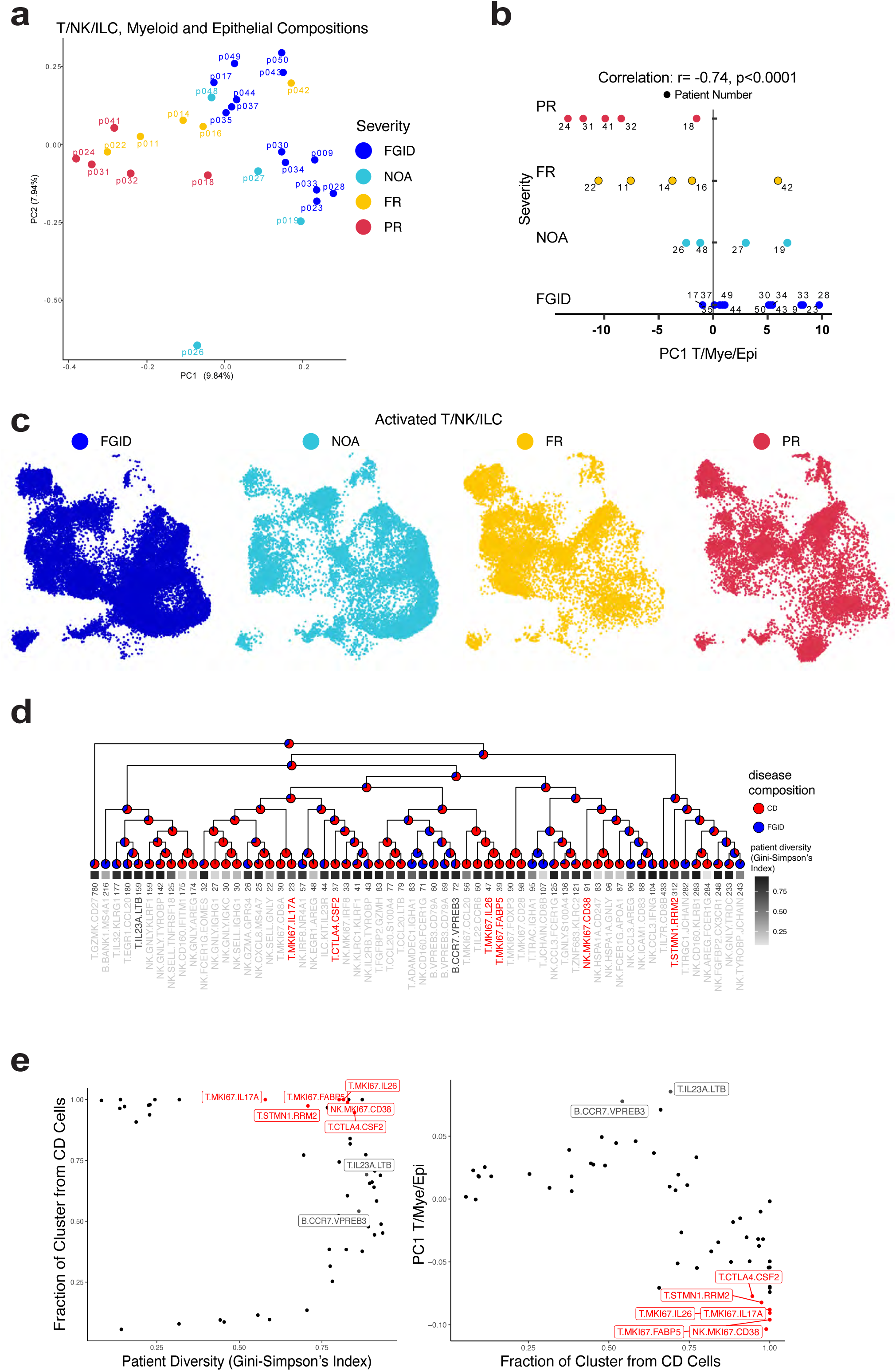
ARBOL analysis of joint FGID and pediCD T/NK/ILCs, Epithelial and Myeloid cells identifies high-diversity clusters that drive disease severity signature. a. A principal component analysis (PCA) plot of each patient’s end cell cluster composition as determined by ARBOL for T/NK/ILC, Myeloid, and Epithelial cells from FGID and pediCD patients where the end cell clusters frequency is calculated amongst total cells and colors represent disease state and treatment response. b. The Spearman rank correlation between PC1 of the joint T/NK/ILC, Myeloid, and Epithelial cell frequency analysis with anti-TNF response. c. A UMAP plot of 7,366 activated T/NK/ILC cells from FGID (colored by disease) and pediCD (colored by treatment response) data sets. d. Hierarchical clustering of activated T/NK/ILC cells from FGID and pediCD data set with input clusters determined based on results of ARBOL, and performed on the median expression of all genes, using complete linkage and distance calculated with Pearson correlation, between each end cell cluster. Simpson’s Index of Diversity represented as 1-Simpson’s where 1 (black) indicates equivalent richness of all patients in that cluster, and 0 (white) indicates a completely patient-specific subset. Numbers represent the number of cells in that cluster. Names of subsets are determined by Disease.CellType.GeneA.GeneB as in **Methods,** and names in red indicate strongest PR-PC1-negative loadings. Pie charts represent the fractional composition of FGID or pediCD cells (note the initial proportions for reference). e. Top JOINT-PC1-negative clusters are both (left) high diversity and (right) almost exclusively found in pediCD patients.

**Figure 7.**
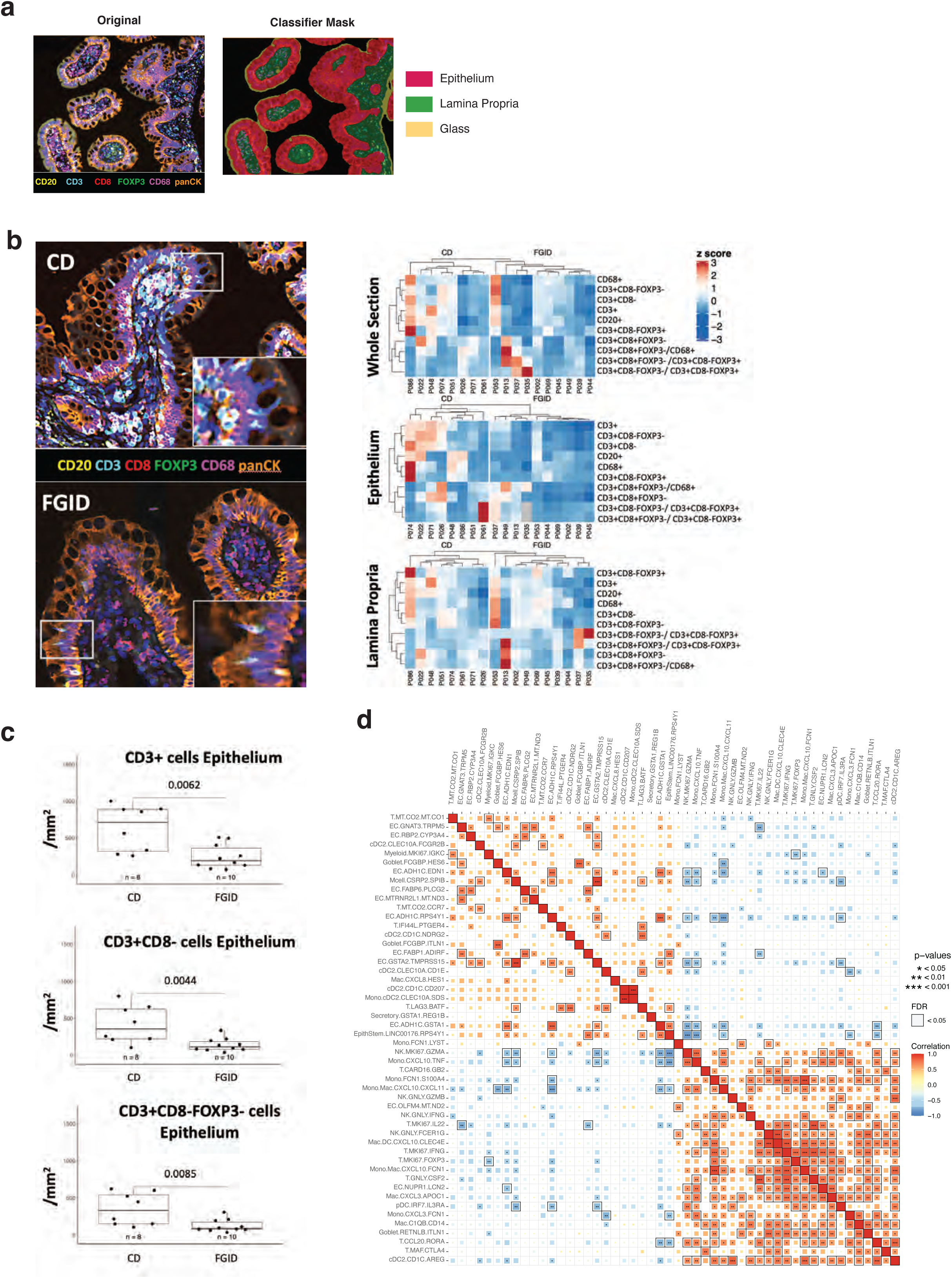
Histological and scRNA-seq analyses indicate CD4+ T cell infiltration into the epithelium of pediCD samples. a. Representative (left) multiplex immunofluorescence of B cells (CD20), T cells (CD3), cytotoxic T cells (CD8), regulatory T cells (FOXP3), myeloid cells (CD68) and epithelium (panCK) which was then (right) analyzed using an automated classifier mask to divide sections into epithelium, lamina propria and glass areas for cellular quantification. b. Representative CD and FGID sections stained with indicated markers, inset focuses on T cells within epithelium. Quantification of 8 pediCD and 10 FGID participants is represented as a row z-scored hierarchically-clustered heatmap for total number of indicated cells per mm^2^ of whole section, epithelium, or lamina propria. c. Cells per mm^2^ of epithelium for CD3+ cells, CD3+CD8-cells (CD4 T cells), and CD3+CD8-FOXP3-(effector CD4 T cells) for pediCD and FGID participants. d. Spearman rank correlation heatmap of the counts-per-million for each of the top 25 clusters defining pediCD-TIME positive (NOA-associated) and pediCD-TIME negative (PR-associated) vectors. Correlation is represented by both the intensity and size of the box and those which are FDR < 0.05 have a bounding box.

**Figure 8.**
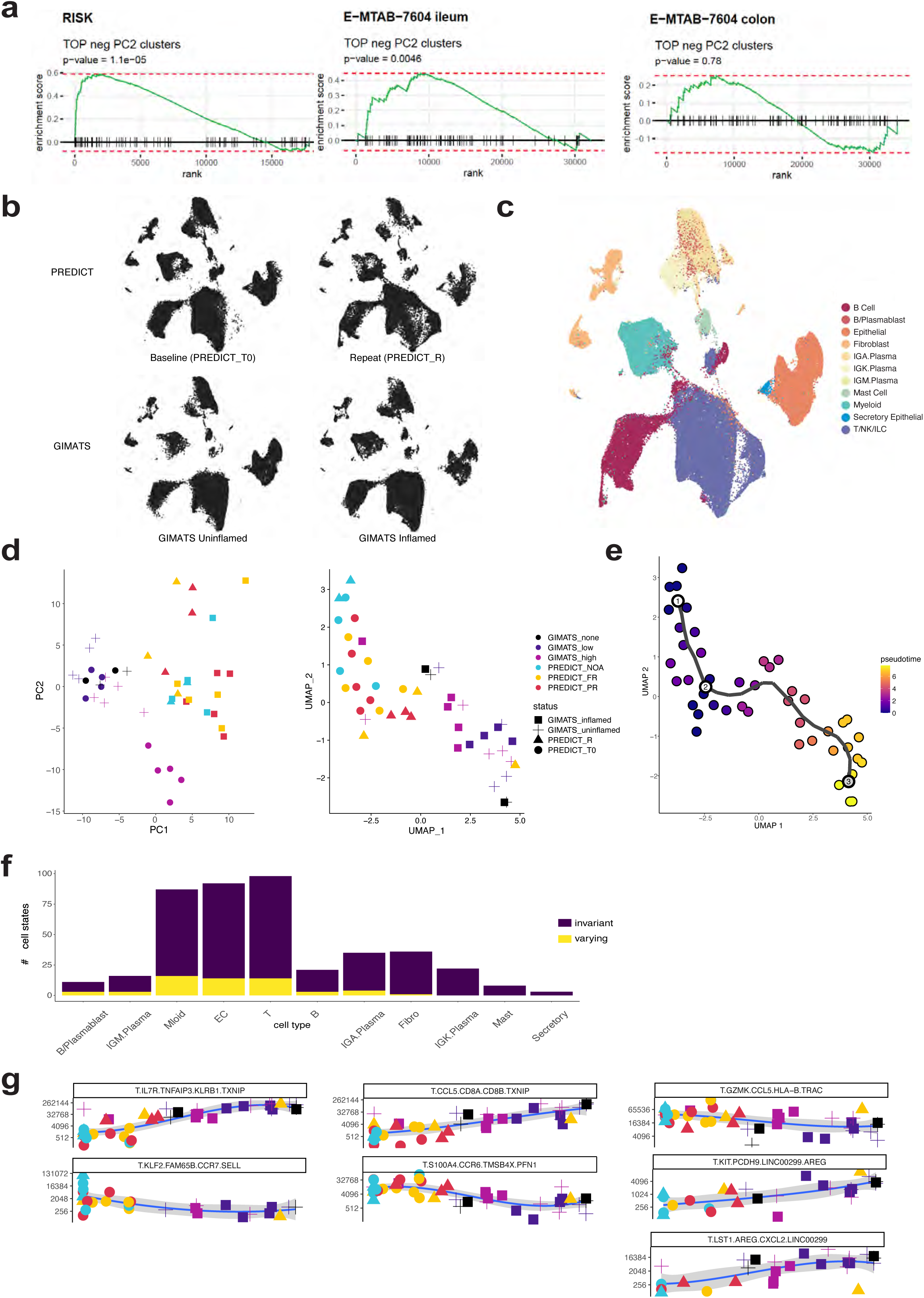
Anti-TNF treatment shifts the pediCD cellular ecosystem towards adult treatment-refractory disease. a. GSEA analysis showing the ranks of 92 PREDICT markers (markers of top 25 cell states associated with disease severity and treatment outcomes) in bulk RNA sequencing of illeal or colonic mucosa of two other treatment-naïve cohorts (pediatric RISK cohort, n = 69, adult E−MTAB−7604 cohort, n = 43) comparing pediCD patients who did or did not respond to anti-TNF therapy. P-value is estimated based on an adaptive multi-level split Monte-Carlo scheme. b. UMAP plots of 275,544 cells from PREDICT ileal biopsies (Baseline: 107,432 cells; Repeat: 47,796 cells) or GIMATS anti-TNF treated ileal resections (GIMATS Uninflamed: 61,965; GIMATS Inflamed 58,351) integrated using Harmony (batch = 10x version). c. A UMAP plot from cells in (**b**) colored by major cell type. d. A PCA plot (left) or UMAP plot (right) of each patient’s end cell cluster composition as determined by ARBOL with Harmony integration (batch = 10x version) for T, B, Myeloid, Fibroblast, Plasma from the treatment-naïve PREDICT pediCD samples (n=13), repeat PREDICT biopsies (n=2 NOA, 6 on-anti-TNF), and adult on-treatment GIMATS biopsies (n=22) where the end cell clusters frequency is calculated amongst total cells and colors represent severity and cohorts are shapes. e. A UMAP trajectory of the CPM table underlying (**d**). f. The top 50 varying (yellow) cell clusters along the pseudotime trajectory as determined by Moran’s I test (q-value < 0.000029). g. Cells per million of significantly varying T/NK/ILC clusters where individual points represent patient biopsies colored by severity and cohorts as shapes (as in **d**).

### Clinical variables and cellular variance that associates with pediCD severity

As the pediCD atlas was curated from treatment-naïve diagnostic samples, we were able to interrogate the data to test if overall shifts in cellular composition, specific cell states, and/or gene expression signatures underlie clinically-appreciated disease severity and treatment decisions (NOA vs. FR/PR), and those that are further associated with response to anti-TNF therapies (FR vs. PR). Here, we leveraged the detailed clinical trajectories collected from all patients as the ultimate functional test: resolving how cellular composition and cell states anticipate disease and treatment outcomes.

In order to capture the overall principal axes of variation explaining changes in cellular composition, we calculated the fractional composition of all 305 end cell clusters in pediCD within its parent cell type (“per cell type”), or within all cells (“per total cells”), and performed a principal component analysis (PCA) over both of these sample x cell cluster frequency tables (**Supplemental Table 14**) (Mathew et al., 2020). We then used the PC1 (13.4% variation “per cell type” and 13.5% variation “per total cells”) and PC2 (12.7% variation “per cell type” and 11.8% variation “per total cells”) values for each sample as numerical variables which we correlated with clinical metadata (**Figure 5a**, r by Spearman-rank). Amongst the clinical variables, we noted strong correlation between Initial wPCDAI and CRP (r=0.83, FDR<0.05), and moderate correlation between Initial wPCDAI and either anti-TNF within 90 days (r=0.65) or anti-TNF response coded as NOA (0) FR (1) and PR (2) (r=0.49). The pediCD atlas also enabled us to correlate the transcriptome with initial disease severity and treatment response: Thus, for PC1-“per total cells”, we identified strong correlations with anti-TNF treatment within 90 days (r=-0.76), and moderate correlation anti-TNF_NOA_FR_PR (r=-0.58; **Methods**).

### Discovery of pediCD-TIME: a collective cell vector that anticipates CD clinical severity and response spectrum

In order to understand if multiple cell types, acting in a concerted manner, were predominantly driving the association with clinical disease severity at initial presentation of pediCD, we then further deconvoluted the overall PCA on 305 end clusters and performed PCA over each cell type’s fractional composition of end clusters individually (B cells: 33 clusters, Endothelial: 18 clusters, Epithelial: 68 clusters, Fibroblast 45 clusters, Myeloid: 54 clusters, T/NK/ILCs: 57 clusters), and correlated the first two PCs (all PC1s and PC2s each accounted for >13% variance) with all of the clinical variables (**Supplemental Table 14**). The PCs derived from T/NK/ILC cells, myeloid cells, and epithelial cells were all moderately correlated with anti-TNF_NOA_FR_PR status (r>0.49) individually, and had higher values than the other cell types; therefore, we asked if a PCA-based metric considering all three cell types would synergistically capture both disease severity and treatment response. When we calculated the PCA accounting for frequencies of each cell subset of T/NK/ILC cells, myeloid cells, and epithelial cells amongst all cell types together, we found strong correlation for PC2 with both anti-TNF within 90 days (r=-0.83) and anti-TNF-NOA_FR_PR status (r=-0.87, FDR<0.05) (**Figure 5a,b**). This represented the two strongest correlations of any variable we tested with anti-TNF treatment and response status. This “PC2-T/NK/ILC/Myeloid/Epithelial” is further referred to as ‘pediCD T cell, innate lymphocyte, myeloid and epithelial’ (pediCD-TIME).

To further deconstruct pediCD-TIME, we used frequency-based statistics and analysis of PC loadings. We first identified which cell clusters accounted for the most significant changes in relative frequency based on the relative frequency of an end cell cluster within its parent cell type, noting limitations related to dissociation-induced biases (**Discussion**) (Gomariz et al., 2018). We performed a Fisher’s exact test between NOA vs. FR; NOA vs. PR; or FR vs. PR, and then performed a Mann-Whitney U test to highlight specific clusters (**Fig. 5c; Supplemental Figure 7; Methods**). As a comparison to differential expression within a cell type or subset, we highlight the power of ARBOL to deconstruct cell clusters that are often unified through strong cell-state signatures, such as proliferation, into identifiable, diverse, and disease-associated cell subsets (**Supplemental Figure 8; Methods**). This cluster-level analysis identified concerted changes when comparing FRs and PRs to NOAs, especially within the T/NK/ILC compartment, with PRs relative to NOAs having additional significant changes within the myeloid compartment (**Fig. 5c; Supplemental Figure 7; Methods**).

As noted above, higher disease severity was inversely related with the PC loadings in pediCD-TIME. Amongst the top negative PC loadings for pediCD-TIME (**Fig. 5a,b**), enriched in FRs—and further enriched in PRs—compared to NOAs, included both helper and cytotoxic T cell clusters (CD.T.MAF.CTLA4; CD.T.CCL20.RORA; CD.T.GNLY.CSF2), NK cell clusters (CD.NK.GNLY.FCER1G; CD.NK.GNLY.IFNG; CD.NK.GNLY.GZMB); proliferating T cells and NK cells (CD.T.MKI67.FOXP3; CD.T.MKI67.IFNG; CD.T.MKI67.IL22; CD.NK.MKI67.GZMA), and monocytes, macrophages, DCs and pDCs (CD.cDC2.CD1C.AREG; CD.Mac.C1QB.CD14; CD.Mono.CXCL3.FCN1; CD.pDC.IRF7.IL3RA; CD.Mono/Mac.CXCL10.FCN1) (**Figure 5d; Supplemental Table 16**). The top positive loadings for pediCD-TIME encompassing the NOA-enriched clusters included several epithelial cell subsets such as Tuft cells (CD.EC.GNAT3.TRPM5) and those with specialized metabolic features including retinol-binding, bile binding and export, fatty-acid and cholesterol metabolism, fructose and glucose metabolism, starch metabolism, glutathione metabolism, sulfation, and the terminal degradation of peptides (CD.EC.RBP2.CYP3A4; CD.EC.FABP6.PLCG2; CD.EC.FABP1.ADIRF; CD.EC.GSTA2.TMPRSS15) (**Figure 5d; Supplemental Table 16**) (Lampen et al., 2000; Mårtensson et al., 1990; Martínez-Augustin and de Medina, 2008; Sullivan et al., 2021; Wen and Rawls, 2020). Furthermore, clusters also enriched in NOA-pediCD-TIME such as CD.EC.ADH1C.RPS4Y1 and CD.EC.ADH1C.GSTA1, clustered in a separate branch together and expressed several enzymes responsible for steroid hormone and dopamine biosynthesis (**Figure 4d, 5d**) (Cima et al., 2004; Magro et al., 2002). Importantly for the regenerative potential of the epithelium, CD.EpithStem.LINC00176.RPS4Y1 were also defining of the NOA-pediCD-TIME direction.

In order to determine the relative contributions and stability of each cell cluster to the pediCD-TIME vector, we performed leave-one-out cross-validation (LOOCV) on the top-weighted pediCD-TIME clusters based on their frequency within total cells (**Supplemental Figure 9a**). This identified the stability of cell clusters such as CD.T.MAF.CTLA4, CD.Mono.CXCL3.FCN1, CD.T.GNLY.CSF2 and other top PR-pediCD-TIME-negative subsets. Furthermore, we also re-calculated PC weights as frequencies of clusters within their major cell type, with the standard deviation of PC loadings stabilizing even further (**Supplemental Figure 9b**). This suggests that compositional scRNA-seq analysis provides more robust signatures within major cell types than amongst all total cells, and both methods can recover a disease trajectory.

Together, these discoveries underscore the multiple collective changes in the composition and/or state of T/NK/ILC cells, myeloid cells, and epithelial cells at pediCD diagnosis that could stratify pediCD patients by disease severity, and may influence anti-TNF responsiveness.

### Cluster-based and cluster-free approaches contextualize pediCD with FGID

As we had generated independent cellular atlases for FGID and pediCD to mitigate aspects of disease batch effects obscuring the ability to resolve cell clusters, we next sought to compare pediCD and FGID cell subsets using label-retaining (Random Forest, ARBOL) and label-free (diffusion maps) methods. We applied these models to each cell type individually, and here focus our discussion on Myeloid cells and T/NK/ILC cells as two cell types prominently associated with pediCD disease severity (**Supplemental Figures 10-12)**. As newer methods are developed, more refined integration is likely to be possible. Comparing across myeloid cells between pediCD and FGID, we could identify strong correspondence of specific cell subsets such as cDC1s or pDCs (**Supplemental Figure 10a**). We also identified strong correspondence between several cDC2 clusters. We identified a gradient of monocyte and macrophage correspondence of 31 clusters in pediCD to 2 FGID clusters, likely reflective of inflammatory monocyte to macrophage differentiation in pediCD (Blériot et al., 2020; Dutertre et al., 2019; Guilliams et al., 2018). For T/NK/ILC cells, we identified more discrete patterns relative to Myeloid cells based on comparison of the RF result. Within the two FGID cytotoxic T cell clusters, we identified correspondence by 18 pediCD clusters, representing ILC3s, and cytotoxic NK cells and T cells (**Supplemental Figure 11**). The cluster of naïve T cells in FGID had correspondence with the majority of pediCD non-cytotoxic T cell clusters, illustrating a substantial activation and specialization to several discrete T cell states that were specific for pediCD.

We next performed an analysis over a shared gene expression space of FGID and pediCD of the monocytes/macrophages (**Supplemental Figure 13**) and T/NK/ILCs (**Supplemental Figure 14**). Within FGID monocytes/macrophages, we identified that the majority of clusters occupied the periphery of the UMAP space, including chemokine-expressing clusters (FG.Mac.CCL3.HES1; FG.Mac.CXCL8.IL1B) and metabolic clusters (FG.Mac.APOE.PTGDS) (**Supplemental Figure 13a,c**). This was in stark contrast to the pediCD monocytes/macrophages, where we identified that now many of the clusters occupied the central region of the UMAP (**Supplemental Figure 13a,c**). We highlight several of the clusters that are significantly different in frequency between the pediCD groups which were found in this central region (**Supplemental Figure 13b**), and how NOA, FR and PR pediCD clusters had significantly different distributions within this space as measured by the Hellinger Distance (**Supplemental Figure 13c-e; Methods**). Of note, the frequency of *TNF*+ macrophages and its expression level of *TNF* was significantly increased in FRs relative to all other groups (**Supplemental Figure 13f;** permutation test shuffling anti-TNF response variable, FR had significantly more TNF+ cells than expected by chance with p approximating 0). Despite the fine-grained tiered clustering approach used, the majority of clusters had high Simpson’s Diversity indices representing cell states found in several patients (**Supplemental Figure 13g**).

Within T/NK/ILCs, we identified that FGID cells were more uniformly mixed with the pediCD cells relative to monocytes/macrophages (**Supplemental Figure 14a**). FGID cells occupied naïve and quiescent states, showed some signatures of activation, and also specialization towards helper and cytotoxic states (**Supplemental Figure 14a,c**) (Sallusto et al., 1999). The most notable changes in the Hellinger Distance distribution occurred between FGID and FR, rather than between FGID and PR (**Supplemental Figure 14d,e**). Similar to the monocytes/macrophages, the main area which gained density with increased disease severity was the central region: characterized by proliferation of several clusters increased in frequency within FRs and PRs, including T cells such as CD.T.MKI67.FOXP3 and CD.T.MKI67.IL22 (**Supplemental Figure 14b-d**). Taken together with several cell clusters associated with pediCD proliferation overlapping with existing areas found in FGID, this indicates activation and more extreme diversification from existing T cell states driving the T cell clustering that defined pediCD. This is distinct from the recruitment and failed differentiation towards homeostatic cell states between FGID and pediCD that we discovered in monocytes/macrophage clusters. Both joint projections confirm and extend our random forest correspondence probabilities, and provide a clustering-independent view of how cellular density is shifted between FGID and pediCD (Dann et al., 2022).

**Joint ARBOL atlas of activated T/NK/ILC’s in FGID and pediCD**

Building on our Random Forest and Hellinger Distance distribution analyses, we sought to formally cluster pediCD and FGID T/NK/ILC, Myeloid, and Epithelial cells together to place the pediCD patients within context of FGID, taking advantage of the high-resolution view afforded by ARBOL (**Fig. 6a**). We tested whether performing ARBOL on a joint FGID and pediCD dataset would also discern a disease severity vector. We identified that JOINT-PC1 in this analysis captures a disease severity vector, with the positive direction enriched in FGID patients, and the negative direction enriched in FR and PR pediCD patients (**Fig. 6b**; r= −0.74, p<0.0001). We focused on a branch of 7,366 activated T/NK/ILCs, annotated all end clusters as T, NK or ILC (**Supplemental Figure 6**), and selected 2 genes from the top-10 rank-scored list (**Methods**) that provide a descriptive name for each cluster (**Fig. 6c,d**). We visualized at each split of the binary tree representation the fractional composition of cells weighted towards FGID or pediCD cells.

Our tree recovered primary splits of memory T cells, cytotoxic cells, helper cells, and intraepithelial cells, with proliferative cells interspersed within these groups. Intriguingly, this analysis reinforces our flow cytometric and initial two-tiered manual clustering approach whereby up to a certain level of the tree, FGID and pediCD do not show significant differences in composition. However, when following the additional splits leading to each end cluster, we now note several clusters which are pediCD or FGID enriched. Furthermore, despite this disease-enrichment, the end clusters are still high in patient diversity, with the majority falling above 0.5 Simpson’s Index (**Figure 6d,e**). Importantly, our top-ranked PR-PC1-negative T cells contributing to the disease severity vector are all high diversity clusters even when clustered jointly with FGID (**Fig. 6; Supplemental Figure 15**). We again recover a majority of proliferating T and NK cell clusters contributing to the JOINT-PC1 disease severity vector (NK.MKI67.CD38, T.MKI67.FABP5, T.MKI67.IL17A, T.MKI67.IL26, T.MKI67.FOXP3, and others).

The joint FGID-pediCD atlas showed an overlap between less severe pediCD patients (NOA) and FGID samples. To understand what causes the alignment between these groups, we used an automated naming method to name the 802 clusters in the joint ARBOL, and plotted the subsets with the most positive loadings in JOINT-PC1 (**Supplemental Figure 15).** We used alluvial diagrams to match the joint cell states with their corresponding curated names from the CD atlas to contextualize the FGID samples in terms of the NOA samples. Doing so revealed subsets of *CCR7+* T cells and *CLEC10A+* cDC2’s are shared between these groups that are less prominent in severe CD (**Supplemental Figure 15e)**. However, in order to determine if these cell states are distinct from healthy T and DC states, we would need to directly integrate healthy data.

While further experiments would need to be conducted to isolate and determine the stable or transient nature of each cell cluster, their conservation across multiple patients from distinct genetic and environmental backgrounds suggests that these end clusters may be reliably found across most patients sampled within a disease category, and that the primary disease-associated signatures remain robust when placed in context of another pediatric gastrointestinal disease. As we had now generated and annotated three atlases: FGID, pediCD, and joint FGID/pediCD, we took the opportunity to calculate how considering atlases separately or jointly influenced the coherence of each end cluster **(Supplemental Figure 15; Methods)**. We anticipate that as atlases scale in cell number, patient number, cohort number, and meta-analyses, that multiple methods of clustering will be important to fully-resolve the cellular heterogeneity present within each sample, while retaining community-level labels for facile integration and comparison for meta-analyses.

### Pair-Wise Correlation of Cell Clusters and Histology

In order to provide spatial quantification for key cell types in our disease signature, we also utilized multiplex immunofluorescence to identify the absolute numbers and locations of B cells, cytotoxic, helper and regulatory T cells, and macrophages in FGID and pediCD samples. Using an automated mask for epithelial and lamina propria identification, we were able to resolve the location of these cell types within the whole section or within the epithelial or lamina propria layers of the tissue (**Figure 7a**). We identified no significant differences in the whole sections for total numbers of T cells (CD3+), B cells (CD20+), myeloid cells (CD68+), and cytotoxic T cells or regulatory T cells between pediCD (n=8) and FGID (n=10) participants (**Figure 7b**). Strikingly, we did find a significant difference in total CD3+ cells within the epithelium, and specifically of CD3+CD8-FOXP3-(CD4+ effector T cells) using our multiplex immunofluorescence panel (**Figure 7c**). This indicates that CD4+ cells infiltrating into the epithelial layer may have an early and outsized role in driving pediCD.

In order to understand the impact of which CD4+ T cell states that were associated with anti-TNF response may be influencing specific epithelial states, we correlated proliferating T and NK cell clusters with epithelial cell clusters (**Figure 7d**). We used the single-cell data for this, where we are powered to make comparisons between response groups. We found that CD.T.MKI67.FOXP3 were strongly positively associated with CD.Goblet.RETNLB.ITLN1 and CD.EC.NUPR1.LCN2 secretory cell states. Conversely, we found that CD.T.MKI67.IL22 were significantly negatively correlated with CD.EC.GNAT3.TRPM5 and EC.FABP1.ADIRF cells, and that CD.T.CCL20.RORA were significantly negatively correlated with EC.ADH1C.GSTA1 and EpithStem.LINC00176.RPS4Y1. This indicates the potential for CD4 T cells present within the epithelial layer to significantly alter epithelial cell states away from nutrient sensing and homeostatic metabolic function in pediCD.

### Contextualization to published Crohn’s Disease Studies

To determine whether the pediCD-TIME disease severity gene signature that we discovered in the PREDICT study can be found in other cohorts with published bulk RNA-seq data, we selected the top 92 markers of the 25 cell states associated with disease severity and treatment outcomes (**Methods; Supplemental Table 17**) and performed a gene-set enrichment analysis (GSEA) (**Figure 8a; Supplemental Figure 16**). We used the cell clusters ranked by cell frequency of total cells, as this would be most representative of bulk-analyzed tissue. In the PREDICT cohort, we were able to validate the 92-gene signature as significantly enriched within FR and PR patients relative to FGID or NOA (all p<9.3×10^-05^). These 92 genes were also enriched in PR relative to FR patients (p<4.3×10^-05^). Notably, in the two independent treatment-naïve cohorts that we analyzed (the pediatric RISK cohort, n = 69, and the adult E−MTAB−7604 cohort, n = 43) this 92-gene signature was significantly enriched in illeal, but not colonic, mucosal biopsies from patients who did not respond to anti-TNF therapy compared to those who responded (**Figure 8a**; p= 0.0046 vs. p=0.78, respectively) (Kugathasan et al., 2017; Verstockt et al., 2019). Thus, these 92 genes (including *TNFAIP6, GZMB, S100A8, CSF2, CLEC4E, S100A9, IL1RN, FCGR1A, CLIC3, CD14, PLA2G7, FAM26F, IL3RA, NKG7, IL32, CCL3, OLR1, LILRA4, APOC1, MYBL2* and others; **Supplemental Table 17**) nominate a signature of anticipatory markers of anti-TNF therapy outcome in newly-diagnosed patients and are validated in one internal (bulk RNA-seq from PREDICT patients) and two external (RISK and E-MTAB-7604) bulk RNA-seq cohorts.

### Anti-TNF treated repeat pediCD biopsies are closer to adult treatment-refractory CD

We sought to test how anti-TNF treatment shifts the cellular ecosystems of the pediatric ileum. The availability of repeat biopsy specimens from PREDICT patients enabled us to perform a joint ARBOL containing treatment-naïve pediCD biopsies (n=14), 8 follow-up endoscopy samples (from 2 NOA and 6 on-anti-TNF PREDICT patients), and 11 adult CD patients from the (Martin et al., 2019) study which identified the GIMATS (T, B, Myeloid, Fibroblast Plasma) cellular module (**Figure 8b**).

Due to the utilization of several 10X 3’ kit versions, we added the integration method Harmony into the ARBOL pipeline (**Figure 8b,c; Methods**). Of note, pediCD atlas curated clusters were preserved with a similar distribution of Shannon entropy in the integrated atlas compared to the joint atlas (**Supplemental Figure 15**). This analysis illustrated that we continue to recover a gradient of disease severity based on our patients’ cell clusters from NOA, to untreated FR and PR. Intriguingly, the repeat anti-TNF treated samples begin to occupy the area between the baseline PREDICT samples and the on-treatment GIMATS adult samples (**Figure 8d; diamonds are repeat samples**). Meanwhile, the two NOA patients with repeat biopsies remain at the beginning of the trajectory overlaid next to their corresponding baseline samples.

From the resultant integrated ARBOL, we calculated a pseudotime trajectory of patients informed by cell clusters. By calculating the cell clusters most auto-correlated along the pseudotime trajectory, this revealed that T/NK/ILC, Myeloid, and Epithelial cell clusters (i.e. the same cell types defining the pediCD-TIME signature) were enriched in those tracking with the trajectory (**Figure 8e,f**). By using the automated naming function within ARBOL, we were able to provide identities for these cell clusters, and present those for T/NK/ILC cells (**Figure 8g**). Our analysis revealed that T.KLF2.FAM65B.CCR7.SELL (expressing markers of naïve T cells), T.GZMK.CCL5.HLA-B.TRAC (expressing markers of CD8 cytotoxic T cells) and T.S100A4.CCR6.TMSB4X.PFN1 (expressing markers of Th17 cells) were significantly decreased with anti-TNF treatment in pediatric and adult patients. Correspondingly, T.IL7R.TNFAIP3,KLRB1 (with signatures of TNF activation), T.CCL5.CD8A.CD8B.TXNIP (effector CD8 T cells), and T.KIT.PCDH9.LINC00299.AREG; T.LST1.AREG.CXCL2.LINC00299 (innate lymphoid cell clusters) increased.

To further characterize which cell states exhibited consistent trends across this disease-associated pseudotime, we conducted additional validation analysis using the TradeSeq package (**Supplemental Figure 17).** Through leave-one-out cross-validation and we assessed the robustness of the association between each cell state and the pseudotime vector. TradeSeq’s trajectory clustering method enabled us to classify the robustly associated cell states into distinct pseudo-temporal patterns (e.g. downward-hill, downward-slope, quick-climb, and upward-hill; **Supplemental Figure 17b).** Notably, bootstrap analysis confirmed that subsets of T, Myeloid, and Epithelial cells consistently exhibited strong associations with pseudotime. This analysis identifies those clusters within the terminal ileum which are most associated with compensation to TNF inhibition, and illustrates that while our NOA patients exhibited remarkable consistency in the pseudotime location of their biopsies within this trajectory, anti-TNF treatment consistently “pushed” a patient’s cells closer to the location of the anti-TNF refractory disease observed in adult CD.

## Discussion

This study addresses a critical unmet need in the fields of IBD and systems immunology: the creation of an atlas of newly-diagnosed untreated diseased tissue, coupled with detailed clinical follow-up to link diagnostic cell types and states with disease trajectory. This is especially true for GI autoimmune disease, and other diseases which affect tissues not easily accessible without operative or endoscopic intervention, and where tissue-specific immune pathology dictates disease severity and trajectory. Likewise, cross-sectional studies, as have been the norm for most previous scRNA-seq studies of IBD, are not able to overlay disease trajectory and treatment response onto the topography of a complex multi-cellular atlas, thus limiting the mechanistic and predictive inferences that can be drawn from the generated atlas (Corridoni et al., 2020b, 2020a; Drokhlyansky et al., 2019; Elmentaite et al., 2020; Huang et al., 2019; Kinchen et al., 2018; Martin et al., 2019; Parikh et al., 2019; Smillie et al., 2019; Uzzan et al., 2022).

Furthermore, mouse models of CD, and of IBD more broadly, may not be the most appropriate models for understanding treatment resistance in pediCD (Neurath, 2019). To surmount these limitations, we created a prospective clinical study, and enrolled patients requiring a diagnostic biopsy for possible IBD, prior to diagnosis. This allowed us to capture a tremendously valuable control group: those patients with FGID, who experience GI symptoms, but for whom the endoscopy does not reveal evidence of GI inflammation or autoimmunity. These uninflamed controls served as a critical comparator to contextualize the evidence of immune pathology that we observed in patients with pediCD across the severity spectrum. With these detailed clinical phenotypes as our foundation, we developed an automated iterative tiered clustering algorithm for scRNA-seq data, ARBOL, which defined pediCD-TIME, a vector of T cells, myeloid cells and epithelial cells that stratifies both pediCD disease severity and response to treatment. We also relate our treatment-naïve samples to repeat samples from PREDICT patients on anti-TNF, and to an adult on-treatment Crohn’s scRNA-seq atlas, identifying a continuum between pediatric and adult Crohn’s disease that may be bridged by the effects of treatment (Martin et al., 2019).

The availability of comprehensive clinical, flow cytometric and scRNA-seq data from patients with pediCD and from uninflamed FGID controls created an unprecedented opportunity for comparative atlas creation. We developed a methodical, unbiased, approach to cell state discovery, ARBOL, released alongside our manuscript in both R (https://github.com/jo-m-lab/ARBOL) and python (https://github.com/jo-m-lab/ARBOLpy). ARBOL iteratively explores axes of variation in scRNA-seq data by clustering and subclustering until specific stop conditions are met. The philosophy of ARBOL is that multiple axes of variation could be biologically meaningful at each tier, and that axes of variation are relative to the comparative outgroup, meaning that similar cell states may arise at distinct tiers. Once these possibilities are explored, curation and a statistical interrogation of resolution are used to collapse clusters into the elemental transcriptomes and co-varying gene expression of the dataset. ARBOL inherently builds a tree of subclustering events. As data is separated by major axes of variation in each subset, later rounds capture less pronounced variables. This comes with some caveats: variation shared by all cell types (for example, cell cycle stage) can make up one of the major axes of variation in the first round of clustering. Cell types can split up at the beginning, so the same splitting of B and T cells, for example, may happen further down in separate branches. The resulting tree of clustering events (**Supplemental Figure 5a**) is therefore neither indicative of true distances between end clusters nor a tree of unique groupings. We address this problem by calculation of a binary tree of manually and computationally curated end clusters. Using a standardized method of end cluster naming, which we describe in ARBOL’s tutorial (https://jo-m-lab.github.io/ARBOL/ARBOLtutorial.html), we found the resulting binary tree assorted end clusters into appreciated cell types and subsets (**Figures 3 and 4**), and also reveals further previously unappreciated granularity that will serve as the foundation for future work into the cellular composition of the gastrointestinal tract.

One of the primary remaining challenges going forward will be to identify which clusters are truly patient-unique, or are simply patient-unique at the cohort size to which we are currently limited to. We calculate a diversity metric for each end cluster to highlight those which are largely conserved between patients, and provide complete cluster-defining gene lists for both FGID and pediCD at three levels of clustering. We also provide links to our data visualization portal to enable cross-atlas comparisons: https://singlecell.broadinstitute.org/single_cell/study/SCP1422/predict-2021-paper-fgid and https://singlecell.broadinstitute.org/single_cell/study/SCP1423/predict-2021-paper-cd.

ARBOL can be deployed in the generation of cell atlases from multiple study designs to yield, for example, separate atlases for distinct diseases, joint atlases of diseases within a study, or integrated larger meta-atlases that merge multiple studies. Recent work from the integrated Human Lung Cell Atlas highlights that under-clustering of cells continues to be a large hurdle in many individual studies, but that higher clustering power must be balanced with the use of more stringent integration methods to merge control and disease groups without obscuring known subsets within lymphoid cells such as regulatory T cells and ILCs (Sikkema et al., 2022). One of the chief advantages of enrolling patients at diagnosis, and prior to any therapeutic intervention, was that we were able to relate their diagnostic immune landscape with disease trajectory. In the pediCD group, we identified 3 clinical subgroups. The first distinction was made by treating physicians, and classified patients with milder versus more severe clinical disease characteristics at diagnosis. The milder patients were not placed on anti-TNF agents (NOA), while the more severe patients were treated with monoclonal antibodies that neutralize TNF including infliximab and adalimumab. The second distinction between patient groups could not be made at diagnosis, but rather, was based on clinical and biochemical response to anti-TNF agents. Thus, of those patients treated with anti-TNF therapeutics, some were FRs, and some were only PRs, with PRs requiring anti-TNF dose modifications and the addition of other agents, and with ongoing, uncontrolled disease signs and symptoms. While differences in anti-TNF pharmacokinetics have been partially implicated in the need to dose-escalate anti-TNF agents in some pediCD patients, our study identifies foundational differences in the immune state at diagnosis in PR patients compared to the NOA and FR subgroups (D’Haens and Deventer, 2021; Ordás et al., 2012; Yarur et al., 2016). Although standard flow cytometry was not able to distinguish the immune phenotype of NOA versus treated patients, scRNA-seq identified significant differences. The contextualization of our scRNA-seq derived predictive cellular vector, pediCD-TIME, with two other treatment-naïve bulk RNA-seq studies of Crohn’s disease, and a reference adult scRNA-seq study, underscores the broader applicability of our findings (Kugathasan et al., 2017; Verstockt et al., 2019).

We noted significant cell state changes at diagnosis underlying clinically-appreciated disease severity that impacted the clinical decision to treat or not to treat with anti-TNF agents. These occurred within multiple clusters of T, NK, fibroblast, epithelial, monocyte, macrophage, and dendritic cells. For anti-TNF response, very few clusters exhibited significantly differential composition between FR and PR individuals. This suggests that multiple collective changes in several cell types may conspire to lead to differences in treatment outcomes. Indeed, when we jointly considered a cellular principal component vector comprising epithelial cells, T/NK/ILCs, and myeloid cells, we identified several clusters that together could delineate the full spectrum of NOA, FR, and PR. This cellular vector (pediCD-TIME) indicated that multiple T cell subsets, NK cells, monocytes, macrophages, and epithelial cells were altered in disease.

Epithelial cells involved in chemosensation (Tuft.GNAT3.TRPM5) and absorption of metabolites (EC.GSTA3.TMPRSS15), as well as stem cells (Banerjee et al., 2020; Sido et al., 1998; von Moltke et al., 2016) were enriched in NOA individuals. That pediCD severity is not uniquely predicted by a singular cell subset or gene is reflective of the complex genetics and environmental factors that have been implicated, along with the rich literature that has found significant changes by histology, flow cytometry, or mass cytometry in CD relative to control tissue (Buisine et al., 2001; Leeb et al., 2003; Leonard et al., 1995; Lilja et al., 2000; Mitsialis et al., 2020; Müller et al., 1998; Souza et al., 1999; Stappenbeck and McGovern, 2017; Takayama et al., 2010). However, with the PREDICT study, we have discovered precisely which changes in CD cellular composition collectively form a vector that anticipates both disease severity and treatment response. Intriguingly, the quantification and visualization of this response vector predicted a later escalation of one of our patients (p022; who appeared as an outlier FR in **Figure 5d**) from FR to PR, which occurred after our database lock. Furthermore, our approach also leverages co-variation of genes across single cells to identify gene sets which are not found by differential expression testing (which ignores associations between genes across cells) yet give rise to recurrent cell states found in multiple patients that are differentially abundant.

When considering the relationships between T cells and NK cells along with epithelial cells, we captured that proliferating cytotoxic NK cell subsets like CD.NK.MKI67.GZMA were significantly negatively correlated with critical metabolic and progenitor epithelial cell subsets in pediCD. Conversely, proliferating regulatory CD.T.MKI67.FOXP3 were positively associated with secretory epithelial cells in pediCD, but did not appear related to the decrease in metabolic or progenitor cells. In addition, we have found that pediCD samples have a significant increase in the number of CD4 T cells localized to the epithelial lining relative to FGID. How T cell-derived cytokines impact intestinal regeneration and differentiation has recently been the focus of several studies, but the relationship of these fine-grained T cell subsets with specific epithelial cell states observed in the human intestine remained unknown (Biton et al., 2018; Lindemans et al., 2015). Furthermore, our study builds on work from other groups who have recently started to describe specific features of intestinal intraepithelial T cells in inflammation and infection (Hoytema van Konijnenburg et al., 2017; Jaeger et al., 2021; Parsa et al., 2022). Our work suggests that there is further complexity to understand, particularly as it pertains to specific subsets of cytotoxic NK cells and T cells, and their impact on epithelial cell homeostasis and regeneration in pediCD.

The mapping of these disease severity-associated cell networks identifies a host of new potential therapeutic targets for pediCD, for many of which there are clinical-stage therapeutics that could be investigated. These include CD40L-blocking antibodies, IL-22 agonists, and targeted anti-proliferation agents (Betts et al., 2017; Furlan et al., 2015; Lindemans et al., 2015; Miura et al., 2021; Ramanujam et al., 2020; Sootome et al., 2020). A case can also be built for targeting inflammatory cytokines such as IL-1β, and for interrogating agents aimed at mucosal healing including new anti-GM-CSF antibodies, given that several prominent cell subsets marked by CSF2 were enriched in the PR patients (Ai et al., 2021; Aschenbrenner et al., 2021; Castro-Dopico et al., 2020; Mehta et al., 2020; Mitsialis et al., 2020; Muro and Mrowiec, 2015). This atlas therefore provides a rigorous evidence-based rationale for proposing new therapeutic interventions, as well as a mechanism for interrogating the impact of new agents on the longitudinal immune landscape of pediCD patients. It also identifies those clusters which respond to anti-TNF treatment and may present new therapeutic targets.

Our analysis underscores the power in the disease-specific clustering employed with this dataset, which revealed principled end clusters in pediCD which could then be related back to a non-inflammatory reference atlas of FGID, and contextualized with published adult CD atlases; while still maintaining fine granularity through distinct computational methods. Recent work on COVID-19 has also highlighted the challenges faced by systems approaches to capture baseline cell states that predict disease trajectory (Kaczorowski et al., 2017; Lucas et al., 2020; Mathew et al., 2020; Schulte-Schrepping et al., 2020; Su et al., 2020). In a disease of known infectious etiology with SARS-CoV-2, monocytes, macrophages, granulocytes, T cells, B cells, antibodies, and interferon state have all independently been associated with disease outcomes. Enabled by larger numbers of participant biopsies profiled at single-cell resolution, recent work in cancer builds on analytical frameworks using dimensional reduction techniques on flow cytometry data to understand collective cellular changes in multi-cellular tissues (Combes et al., 2022; Kaczorowski et al., 2017; Nieto et al., 2021). This work has helped establish the paradigm of tissue “archetypes” or “immunotypes” that represent the collective cellular system. Here, we have built on this foundation to consider how multiple collective changes at baseline may influence outcome, yet are likely more reflective of the disease. With the complex and protracted presentation of a multifactorial disease like Crohn’s disease, our data suggest that multiple concerted effects are required to dictate both the severity (NOA vs FR/PR) and the treatment-response (FR vs PR). Future work will further consider which cell subsets are recovered during mucosal healing, and how closely the treated state reflects each individual patient’s baseline presentation. Our work taken together with our colleagues’ points to the emergent opportunity to place a patient’s tissue in the broader cellular context of health and disease.

### Limitations of the Study

Our study has several limitations that are important to consider. Despite having profiled more treatment-naïve patients than any other studies at single-cell resolution, we are still limited by the cohort size to the strict inclusion/exclusion criteria used, follow-up period, and the ability to capture untreated patients early during the diagnosis process. As a result, we are underpowered to directly correlate in our cohort genetic and environmental influences such as the enteric microbiome, infections, and dietary factors (Dovrolis et al., 2020; Rajca et al., 2014; Yilmaz et al., 2019). Second, we are limited in our ability to capture full spatial transcriptomic and multiplexed immunofluorescence information from our initial cohort. It will also be critical to understand how disease progresses in larger numbers of patients from our cohort, and the reason for persistent NOA, FR, or PR patient states in some individuals which will require retention in the trial and additional repeat biopsy procurement from larger numbers of participants. Third, our cohort is representative of the demographics of FGID and IBD in Seattle, WA. It will be essential to understand how both FGID and IBD may present differently across the world, and multiple cohorts will be required to understand location-unique and generalizable findings. Fourth, we also recognize the number of pediCD patients within this cohort limits our ability to subdivide CD phenotype into location and behavior of disease (via the Paris/Montreal classification systems) (Levine et al., 2011). Fifth, we also highlight the rapidly-evolving field of approaches to integrate single-cell datasets towards a larger gut cell atlas and the trade-offs in resolution that occur as additional datasets are brought in that require more stringent integration methods. We recognize that integration methods, as well as the use of UMAP to represent multiple PC’s in two dimensions, which, while helpful to jointly visualize studies together, may lower the resolution, and in some cases even make cell subsets like T, NK, or ILCs challenging to discern (Sikkema et al., 2022). Future work will need to understand specific platform differences that drive integration to help specifically separate batch from biology. Finally, as we are working with small pediatric IBD biopsies, our ability to functionally test and validate the stability of these end cell clusters was limited in this study. Based on our study, we will seek to expand our efforts to validate and extend our finding in ongoing work.

## ACKNOWLEDGMENTS

We thank the patients and their families for helping and contributing to our study. We thank Kathy McConville and Sarah Mbonde for identifying and recruiting patients for PREDICT. We thank Carly G.K. Ziegler for discussions and advice related to iterative tiered clustering, and all members of the Kean, Ordovas-Montanes and Shalek labs for thoughtful discussions. J.O.-M is a New York Stem Cell Foundation – Robertson Investigator. J.O.-M was supported by the Richard and Susan Smith Family Foundation, the AGA Research Foundation’s AGA-Takeda Pharmaceuticals Research Scholar Award in IBD – AGA2020-13-01, the HDDC Pilot and Feasibility P30 DK034854, the Food Allergy Science Initiative, the Leona M. and Harry B. Helmsley Charitable Trust, The Pew Charitable Trusts Biomedical Scholars, The Broad NextGen Award, The Mathers Foundation, The Manton Foundation, and The New York Stem Cell Foundation. L.S.K is supported by NIH P01 1P01HL158504, R01 5R01HL095791, U19 U19AI051731, and by the Helmsley Charitable Trust. V.N. was supported by the International mobility of research, technical and administrative staff of research organizations (CZ.02.2.69/0.0/0.0/18_053/0016981). A.K.S. was supported, in part, by the Searle Scholars Program, the Beckman Young Investigator Program, a Sloan Fellowship in Chemistry, and the NIH (5U24AI118672, 2R01HL095791). V.M. reports research support from Novartis. V.T. Is supported by ASTCT New Investigator Award and CIBMTR/Be the Match Foundation Amy Strelzer Manasevit Research Program Award. S.B.S. is supported by NIH grants P30 DK034854 and RC2 DK122532, and the Helmsley Charitable Trust.

## AUTHOR CONTRIBUTIONS

Conceptualization: G.W., A.Y., A.K.S., L.S.K., H.B.Z., J.O.-M

Methodology: B.A.D., K.K., A.Y., L.A., G.W., D.L.S., D.L., V.N., A.K.S., L.S.K., V.T., H.B.Z., J.O.-M

Formal analysis: B.A.D., K.K., A.Y., V.N., X.D., V.T., H.B.Z., M.F., M.S., J.S.C., J.O.-M

Investigation: B.A.D., K.K., L.A., K.C., R.F., P.K., A.A., L.C., C.M., A.Y., G.W., D.L.S., D.L., M.F., V.N., L.S.K., X.D., G.D., B.B., K.B., V.T., L.V.C., V.M., N.F., B.K., S.C., S.J., Y.Y., M.D., M.W., S.H., C.K., H.B.Z., S.B.S., J.O.-M

Resources: B.A.D., K.K., A.K.S. L.S.K., H.B.Z., J.O.-M

Data Curation: B.A.D., K.K., A.Y., P.K., M.F., V.N., V.T., H.B.Z., J.O.-M

Writing – Original Draft: B.A.D., K.K., V.N., A.K.S., L.S.K., H.B.Z., M.S., J.S.C., J.O.-M

Writing – Review & Editing: B.A.D., K.K., L.A., G.W., D.L.S., A.Y., A.A., P.K., K.C., R.F., B.B., K.B., L.C., C.M., D.L., R.vE., A.C.K., N.F., B.K., S.C., S.J., Y.Y., M.D., M.W., S.H., G.D.K., A.H., W.K.L., S.H., Y.W., M.F., V.N., G.D., A.K.S., L.S.K., V.M., V.T., L.V.C., H.B.Z., M.S., J.S.C., F.T., S.B.S., J.O.-M

Visualization: B.A.D., K.K., V.N., A.K.S. H.B.Z., M.S., J.O.-M

Supervision: A.K.S., L.S.K., J.O.-M

Project Administration: A.K.S., L.S.K., B.B., G.D.K., A.H., W.K.L., S.H., Y.W., H.B.Z., F.T., J.O.-M

Funding Acquisition: A.K.S., L.S.K., J.O.-M

## ETHICS DECLARATIONS

J.O.-M. reports compensation for consulting services with Cellarity and Tessel Biosciences. A.K.S. reports compensation for consulting and/or SAB membership from Merck, Honeycomb Biotechnologies, Cellarity, Repertoire Immune Medicines, Hovione, Third Rock Ventures, Ochre Bio, FL82, Senda Biosciences, Relation Therapeutics, Empress Therapeutics, IntrECate Biotherapeutics, and Dahlia Biosciences unrelated to this work. A.K.S. has received research support from Merck, Novartis, Leo Pharma, Janssen, the Bill and Melinda Gates Foundation, the Moore Foundation, the Pew-Stewart Trust, Foundation MIT, the Chan Zuckerberg Initiative, Novo Nordisk and the FDA unrelated to this work. Dr. Kean is on the scientific advisory board for HiFiBio and Mammoth Biosciences. She reports research funding from Kymab Limited, Magenta Therapeutics, BlueBird Bio, and Regeneron Pharmaceuticals. She reports consulting fees from Equillium, FortySeven Inc, Novartis Inc, EMD Serono, Gilead Sciences, Vertex Pharmaceuticals, and Takeda Pharmaceuticals. Dr. Kean reports grants and personal fees from Bristol Myers Squibb that are managed under an agreement with Harvard Medical School. N.F., B.K., S.C., S.J., Y.Y., M.D., M.F.W., S.H., G.D.K., A.H., W.K.L., S.H., Y.W. are employees and shareholders of Regeneron Pharmaceuticals, Inc. L.A. is a consultant for Takeda Pharmaceuticals. G.W. reports Research funding from Abbvie, Jansen, Takeda, Allakos; SAB for Abbvie, Bristol Myers Squibb; DSMB for Abbvie. D.L.S., is the co-founder, CM, president of NiMBAL Health. S.B.S. declares the following interests: Scientific advisory board participation for Pfizer, Lilly, IFM therapeutics, Merck, Pandion, and Takeda Inc., and grant support from Pfizer, Novartis, Amgen, Takeda Consulting for Hoffman La Roche and Amgen. A.K.S., L.S.K., J.O.-M., H.B.Z., K.K. and B.A.D., are co-inventors on a provisional patent application relating to methods of stratifying and treating IBD.

## Supplemental Tables

**Supplemental Table 1: Flow cytometry panels**

**Supplemental Table 2: scRNA-seq sample filtering thresholds**

**Supplemental Table 3: scRNA-seq quality control metrics for all single-cells**

**Supplemental Table 4: Traditional joint clustering for broad cell types and differential expression testing by Wilcoxon to determine CD or FGID-enriched genes.**

**Supplemental Table 5: FGID cell type markers**

**Supplemental Table 6: FGID cell subset markers**

**Supplemental Table 7: FGID cell state (i.e. end cell cluster) markers**

**Supplemental Table 8: CD cell type markers**

**Supplemental Table 9: CD cell subset markers**

**Supplemental Table 10: CD cell state (i.e. end cell cluster) markers**

**Supplemental Table 11: FGID end cell cluster descriptive names and short curated names look up table**

**Supplemental Table 12: CD end cell cluster descriptive names and short curated names look up table Supplemental Table 13: CD cell type markers**

**Supplemental Table 13: Number of cells per patient per end cell cluster**

**Supplemental Table 14: PCA Loadings for joint Epithelial, Myeloid, T/NK/ILC vectors**

**Supplemental Table 15: Differential composition testing for NOA vs FR, NOA vs PR and PR vs FR categories of anti-TNF response with CD patients**

**Supplemental Table 16: pediCD-TIME top positive and negative cell clusters**

**Supplemental Table 17: 92 markers derived from pediCD-TIME used for bulk RNA seq extension**

## METHODS

### DATA AVAILABILITY

#### Data and Code Availability

Single-cell RNA-seq data can be visualized via the Single Cell Portal links for each atlas of FGID https://singlecell.broadinstitute.org/single_cell/study/SCP1422/predict-2021-paper-fgid and pediCD https://singlecell.broadinstitute.org/single_cell/study/SCP1423/predict-2021-paper-cd. The cell-by-gene matrices will be available with the peer-reviewed version of this manuscript. The raw human FASTQ files will be available from the Broad controlled access repository DUOS with the peer-reviewed version of this manuscript. Please contact the authors for further information.

All original code has been assembled as the ARBOL package and deposited at GitHub and is publicly available together with this manuscript for both R and Python versions at: https://github.com/jo-m-lab/ARBOL and https://github.com/jo-m-lab/ARBOLpy and https://jo-m-lab.github.io/ARBOL/ARBOLtutorial.html.

### PATIENT COHORT DETAILS

#### Study Population and Clinical Parameters

Pediatric patients less than 20 years of age with suspected inflammatory bowel disease were enrolled on the PREDICT Study (ClinicalTrials.gov# NCT03369353). Enrollment took place between November 9, 2017 to December 21, 2018 in accordance with the Fred Hutch Institutional Review Board (Protocol #9730, ethical approval given) with written informed consent and assent when applicable. Patients were enrolled at diagnostic visit and we considered those diagnosed with Crohn’s Disease (CD) and patients without gut inflammation on endoscopy and histology, and who were diagnosed with Functional GI Disease (FGID), were included on this study. Terminal ileum biopsies and blood samples were taken during the diagnostic endoscopy procedures prior to initiation of therapy. Patients diagnosed with other inflammatory or infectious etiologies on endoscopy and biopsy were excluded from the analysis. Clinical course and variables were monitored at the time of enrollment and for 3 years after initial endoscopy, with median follow up for CD being 32.5 months and FGID being 31 months at the time of clinical database lock (December 1, 2020). Medical management was dictated by clinicians. Clinical variables obtained included sex, race, age at diagnosis, weight z-score, height z-score, BMI z-score, clinical disease severity using the Pediatric Crohn’s Disease Activity Index (PCDAI), and disease location and phenotype using the Montreal Criteria (Hyams et al., 1991; Silverberg et al., 2005). Laboratory evaluation included C-reactive protein, ESR, hemoglobin, albumin, white blood cell count, and platelet count.

#### Response to Anti-TNF therapy

Early anti-TNF or immunomodulator therapy was defined as initiation of immunosuppression within 90 days of diagnostic endoscopy. Anti-TNF monoclonal antibody was started in 10 patients with CD. All patients were followed prospectively and categorized as full responders (FR), partial responders (PR), or not on anti-TNF (NOA). Full response to anti-TNF is defined as clinical symptom control and biochemical response with wPCDAI score of <12.5 on maintenance anti-TNF therapy and partial response defined as lack of clinical symptom control and biochemical response with documented escalation of anti-TNF therapy at 2 years after baseline visit.

#### Clinical Statistical Analysis

Clinical variables are expressed as median (lower and upper confidence interval; range) and compared using the Mann-Whitney U test. Categorical variables were described as frequencies and percentages and compared using the chi-square test. Clinical laboratory values are represented by mean and standard error of the mean (range) and compared with the Mann-Whitney U test. Significance is indicated by a P value of <0.05. Clinical statistical analysis was performed using GraphPad Prism version 8.3.0.

### EXPERIMENTAL METHOD DETAILS

#### Tissue Dissociation into Single-Cell Suspensions

##### Human Ileum

Single-cell suspensions were collected from intestinal biopsies using a modified version of a previously published protocol (Persson et al., 2013) as described in (Smillie et al., 2019). One biopsy from the terminal ileum was received directly in hand and processed with an average time from patient to loading on the 10X Chromium platform of 2.5 total hours, and never exceeding 3.5 hours. While intact, biopsy bites were handled using a P1000 pipette applying gentle suction, and all centrifugation steps done in a temperature controlled 4°C centrifuge.

Biopsy bites were first rinsed in 30 mL of ice-cold PBS (ThermoFisher 10010-049) and allowed to settle. Each individual bite was then transferred to 10 mL epithelial cell solution (HBSS Ca/Mg-Free [ThermoFisher 14175-103], 10 mM EDTA [ThermoFisher AM9261], 100 U/ml penicillin [ThermoFisher 15140-122], 100 μg/mL streptomycin [ThermoFisher 15140-122], 10 mM HEPES [ThermoFisher 15630-080], and 2% FCS [ThermoFisher 10082-147]) freshly supplemented with 200 μL of 0.5M EDTA. Separation of the epithelial layer from the underlying lamina propria was performed for 15 minutes at 37°C with rotation at 120RPM. The tube was then removed and placed on ice immediately for 10 minutes before shaking vigorously 15 times. Visual macroscopic inspection of the tube at this point yielded visible epithelial sheets, and microscopic examination confirmed the presence of single-layer sheets and crypt-like structures.

The remnant tissue bite was carefully removed and placed into a large volume (40mL) of ice-cold PBS to rinse before transferring to 5mL of enzymatic digestion mix (Base: RPMI1640, 100 U/ml penicillin [ThermoFisher 15140-122], 100 μg/mL streptomycin [ThermoFisher 15140-122], 10 mM HEPES [ThermoFisher 15630-080], 2% FCS [ThermoFisher 10082-147], & 50 μg/mL gentamicin [ThermoFisher 15750-060]), freshly supplement immediately before with 100 μg/mL of Liberase TM [Roche 5401127001] and 100 μg/mL of DNase I [Roche 10104159001]), at 37°C with 120 rpm rotation for 30 minutes. During this 30-minute lamina propria (LP) digestion, the epithelial (EPI) fraction was spun down at 400g for 7 minutes and resuspended in 1 mL of epithelial cell solution before transferring to a 1.5mL Eppendorf tube in order to minimize time spent centrifuging and provide a more concentrated cell pellet. Cells were spun down at 800g for 2 minutes and resuspended in TrypLE express enzyme [ThermoFisher 12604013] for 5 minutes in a 37°C bath followed by gentle trituration with a P1000 pipette. Cells were spun down at 800g for 2 minutes and resuspended in ACK lysis buffer [ThermoFisher A1049201] for 3 minutes on ice to remove red blood cells, even if no RBC contamination was visibly observed in order to maintain consistency across samples. Cells were spun down at 800g for 2 minutes and resuspended in 1 mL of epithelial cell solution and placed on ice for 3 minutes before triturating with a P1000 pipette and filtering into a new Eppendorf tube through a 40 μM cell strainer [Falcon/VWR 21008-949]. Cells were spun down at 800g for 2 minutes and then resuspended in 200 μL of epithelial cell solution and placed on ice while final steps of LP dissociation occurred. After 30 minutes, the LP enzymatic dissociation was quenched by addition of 1ml of 100% FCS [ThermoFisher 10082-147] and 80 μL of 0.5M EDTA and placing on ice for five minutes. Samples were typically fully dissociated at this step and after gentle trituration with a P1000 pipette filtered through a 40μM cell strainer into a new 50 mL conical tube and rinsed with PBS to 30 mL total volume. This tube was spun down at 400g for 10 minutes and resuspended in 1 mL of ACK and placed on ice for 3 minutes. LP cells were spun down at 800g for 2 minutes and resuspended in 1 mL of epithelial cell solution and spun down at 800g for 2 minutes and resuspended in 200 μL of epithelial cell solution and placed on ice. Following centrifugation, the cells from both EPI and LP fractions were counted and prepared as a single-cell suspension for scRNA-seq. Since the full EPI isolation was not performed on all patients limiting sample sizes, here we focus our analysis on LP fractions that still retain ∼20% of deeper crypt epithelial cells.

#### Flow Cytometry

Multicolor flow cytometry was performed on tissue samples to examine the immune composition for enrolled patients and acquired on a BD Fortessa SORI. Antibodies used include: CD3 APC, SP34-2 (BD Biosciences); CD3 BUV661, UCHT1 (BD Biosciences); CD3 BV711, OKT3, (Biolegend); CD3 PE, SP34 (BD Biosciences); CD4 BV785, OKT4 (Biolegend); CD8a BUV395, RPA-T8 (BD Biosciences); CD8b FITC, REA715 (Miltenyi Biotec); CD11b APC-Cy7, ICRF44 (BD Biosciences); CD11c APC-eFlour 780, BU15 (Fisher Scientific); CD11c BUV661, B-ly6 (BD Biosciences); CD14 APC-eFluor 780, 61D3 (Fisher Scientific); CD14 BUV737, M5E2 (BD Biosciences); CD20 APC-eFluor 780, 2H7 (Fisher Scientific); CD20 PE-Cy7, L27 (BD Biosciences); CD38 APC, HIT2 (BD Biosciences); CD45 PerCP/Cy5.5, HI30 (Biolegend); CD45RA BV605, HI100 (Biolegend); CD56 (NCAM) FITC, TULY56 (Fisher); CD94 APC-Vio770, REA113 (Miltenyi Biotec); CD117 (c-kit) BV421, 104D2 (Biolegend); CD123 BV711, 9F5 (BD Biosciences); CD127 Biotin, HIL-7R-M21 (BD Biosciences); CD161 BV711, DX12 (BD Biosciences); CD197 (CCR7) BV421, GO43H7 (Biolegend); CD294 (CRTH2) BV605, BM16 (Biolegend); CD326 (Epcam) APC, HEA-125 (Miltenyi Biotec); HLA-DR APC-H7, L243 (G46-6) (BD Biosciences); TCR PAN γδ PE-Cy7, IMMU510 (Beckman Coulter); α4-β7 integrin (Act-1), (NIH AIDS Reagent Program); Streptavidin BUV737 (Fisher); Live/dead Fix Aqua (Fisher); R-PE Antibody Labeling Kit (300 mcg) (Abcam). All antibodies used are found in **Supplemental Table 1**. Flowjo software was used to phenotypically define cell populations and compared in patients using two-way ANOVAs (or non-parametric equivalent) using GraphPad PRISM.

#### Methods to Generate Single-Cell RNA-seq Libraries and Sequencing

##### 10X v2 3’

Single cells were loaded onto 3’ library chips as per the manufacturers protocol for Chromium Single Cell 3’ Library (v2) (10X Genomics). The LP fraction was captured in its own channel of the 10X Chromium Single Cell Platform, in order to recover sufficient numbers of cells for downstream analyses. An input of 10,000 single cells was added to each channel with a recovery rate of 9,514 cells per sample based on median across samples. Briefly, single cells were portioned into Gel Beads in Emulsion (GEMs) in the Chromium controller with cell lysis and barcoded reverse transcription of RNA, followed by cDNA amplification, enzymatic fragmentation and 5’ adaptor and sample index attachment. Libraries were sequenced on a HiSeq or NovaSeq flow cell. The read structure was paired end with length of read 1 26bp, length of read 2 91bp, and the length if index 1 (i7 primer) 8bp. Quality-filtered base calls were converted to demultiplexed FASTQ files.

#### Alignment and Filtering

FASTQ files were aligned to GRCh38 using Cellranger v2.2 pipeline on the Cumulus/Terra cloud pipeline https://portal.firecloud.org/?return=firecloud#methods/cumulus/cellranger_workflow/10 generating 27 cell-by-gene matrices (13 FGID, 14 CD), one for each patient. We used default parameters of the 10^th^ snapshot version of the pipeline, aside from requiring that it use cellranger v2.2.0.

Every sample was first filtered excluding genes measured in fewer than 3 cells and cells with fewer than 200 unique genes. To control for doublets and low-quality cells we then further filtered individually, attempting to match the approximate 10,000 cells loaded onto the sample lane and balancing the thresholds to not cut out dense regions of a Ncounts by Nfeatures scatter plot. Pre-filtering, we looked for outlier samples, based on proportion of percent mitochondrial genes, number of counts, and number of features, none fell beyond the 1.5 times the IQR threshold.

**Exact thresholds used for each sample:**

| Sample ID | Diagnosis | min_cells | min_genes | max_genes | max_count | N_cells_post |
| --- | --- | --- | --- | --- | --- | --- |
| p011_T0D_ILE_LPS_3p | CD | 3 | 200 | 3000 | 20000 | 11087 |
| p014_T0D_ILE_LPS_3p | CD | 3 | 200 | 3000 | 20000 | 7285 |
| p016_T0D_ILE_LPS_3p | CD | 3 | 200 | 3000 | 20000 | 7713 |
| p018_T0D_ILE_LPS_3p | CD | 3 | 300 | 4000 | 20000 | 15469 |
| p019_T0D_ILE_LPS_3p | CD | 3 | 200 | 5000 | 20000 | 11200 |
| p022_T0D_ILE_LPS_3p | CD | 3 | 200 | 3000 | 20000 | 7798 |
| p024_T0D_ILE_LPS_3p | CD | 3 | 200 | 3000 | 20000 | 9705 |
| p026_T0D_ILE_LPS_3p | CD | 3 | 300 | 4000 | 30000 | 10847 |
| p027_T0D_ILE_LPS_3p | CD | 3 | 200 | 4000 | 40000 | 8124 |
| p031_T0D_ILE_LPS_3p | CD | 3 | 200 | 3000 | 20000 | 12177 |
| p032_T0D_ILE_LPS_3p | CD | 3 | 200 | 3000 | 20000 | 9052 |
| p041_T0D_ILE_LPS_3p | CD | 3 | 200 | 4000 | 40000 | 7508 |
| p042_T0D_ILE_LPS_3p | CD | 3 | 200 | 4000 | 30000 | 12219 |
| p048_T0D_ILE_LPS_3p | CD | 3 | 200 | 4000 | 40000 | 9158 |
| p009_T0D_ILE_LPS_3p | FGID | 3 | 200 | 3000 | 20000 | 11426 |
| p017_T0D_ILE_LPS_3p | FGID | 3 | 200 | 3000 | 20000 | 11107 |
| p023_T0D_ILE_LPS_3p | FGID | 3 | 200 | 3000 | 20000 | 11262 |
| p028_T0D_ILE_LPS_3p | FGID | 3 | 200 | 4000 | 20000 | 10293 |
| p030_T0D_ILE_LPS_3p | FGID | 3 | 200 | 4000 | 30000 | 9590 |
| p033_T0D_ILE_LPS_3p | FGID | 3 | 200 | 3000 | 20000 | 9514 |
| p034_T0D_ILE_LPS_3p | FGID | 3 | 200 | 3000 | 20000 | 8142 |
| p035_T0D_ILE_LPS_3p | FGID | 3 | 200 | 3000 | 20000 | 5668 |
| p037_T0D_ILE_LPS_3p | FGID | 3 | 200 | 3000 | 20000 | 4210 |
| p043_T0D_ILE_LPS_3p | FGID | 3 | 200 | 5000 | 30000 | 5045 |
| p044_T0D_ILE_LPS_3p | FGID | 3 | 350 | 4000 | 30000 | 12727 |
| p049_T0D_ILE_LPS_3p | FGID | 3 | 200 | 3000 | 20000 | 8003 |
| p050_T0D_ILE_LPS_3p | FGID | 3 | 200 | 4000 | 30000 | 8582 |

Post filtering, we merged sample matrices using an outer join to create an FGID dataset (115,569 cells) and a pediCD dataset (139,342 cells).

### COMPUTATIONAL AND STATISTICAL ANALYSIS

#### Preprocessing & Clustering of scRNA-seq Data

##### 1^st^ Approach – Classical Methods on Combined Dataset

We began initial analysis following traditional clustering and annotation techniques; however, these methods using manual and at times subjective metrics scaled poorly to the size and scope of our dataset and moreover did not give clear distinction between disease specific cell states and compositional shifts within cell states across disease.

For our first pass at analysis, we grouped the FGID and pediCD datasets together (254,911 cells) and proceeded with the standard Seurat v3.1.5 pipeline (Stuart et al., 2019). We used manual heuristics of gene marker specificity to choose cluster resolution and isolate 9 major cell types (T, B, plasma, epithelial, endothelial, fibroblast, myeloid, mast cell, and glial) and 1 aggregate cluster of T, B, myeloid, and epithelial cells with a strong proliferation signature. We then subclustered the proliferating group and manually merged the proliferating cells with their corresponding cell type based on marker gene expression, and separately re-preprocessed and clustered each cell type annotating based on one vs. rest differential expression (Wilcoxon, fdr < 0.05) within the cell type.

We found several disadvantages to this approach. First, we found it difficult to determine for each cluster whether we should be looking for changes in compositional frequency or gene expression. Particularly within the myeloid major cell group we found extremely disease biased sub-clusters, as much as a 9:1 ratio between pediCD to FGID. It was unclear whether there was massive compositional shift within a conserved cell state or if instead a base cell state was split into multiple clusters based on phenotypic differences in disease and we should perform a differential expression test between it and neighboring FGID biased clusters. Second, after two rounds of manual clustering, some cell types could be clustered more deeply based on pre-existing knowledge, whereas other types that have been less well-resolved may have heterogeneity that has yet to be well-appreciated. Third, having partitioned over 100 distinct clusters, individually supervising each subsets processing and sub-clustering was infeasible. We needed a more systematic method to address these challenges.

#### 2^nd^ approach – ARBOL: Iterative Tiered Clustering (ITC) on Separated Disease Conditions

To be able to choose the appropriate future analyses and comparisons, we need a highly accurate representation of cell identity and state. The underlying issue in our first pass at clustering was that in combining the disease conditions together, the variable genes selected at each stage represented a combination of differences between cell identity & disease. This combination could have been manageable if either disease or cell identity were consistently more variable. We could isolate one factor at a specific tier in the hierarchy before sub-clustering to isolate the other. In our case, disease and cell identity both had many overlapping scales of variation. To address this effect, we isolated cell identity by separating our dataset by disease and clustering for cell identity within each disease set (FGID 115,569 cells, pediCD 139,342 cells). This approach required us to perform an additional stage of analysis to find corresponding clusters between the two datasets, but allowed us to more effectively distinguish type, scale, and specificity of disease differences.

Within each disease set we still needed a method to ensure we were reaching the bottom level of biological heterogeneity, and preferably an automated method as our first pass had shown the potential for isolating hundreds of cell states. To efficiently cluster and isolate these cell states we wrote a cloud-based pipeline to systematically optimize parameter selection and stop when biological heterogeneity is exhausted. Homogenous cell subsets were isolated by recursively normalizing, selecting variable genes, and clustering based on silhouette score. We stopped recursing into sub-clusters once we reached one of four end conditions defined as:

1. Having a group of less than 100 cells (though we did partition many clusters smaller than 100 cells after clustering groups just larger than that cutoff).
2. Isolating an optimized clustering of only one cluster.
3. Finding two clusters that have fewer than 25 genes (fdr< 0.05 & |log fold change| > 1.5 & percent expression >= 20% in at least one cluster) differentially upregulated between each cluster using a bimodal test developed in (Shekhar2016 10.1016/j.cell.2016.07.054). For this last condition, if reached, we reject the clustering and return back the cells as a single end cluster.
4. Having reached a max tier limit. Setting this value to 10, we never triggered this condition with either FGID or pediCD datasets, but included it to prevent runaway recursion.

Code for generating this tree of cell clusters is currently available here: (https://jo-m-lab.github.io/ARBOL/ARBOLtutorial.html. Within each recursion, the established steps were processed using Seurat version 3.1.5 (https://github.com/satija.lab/seurat). Normalization and variable gene selection were processed with SCTransform (https://github.com/ChristophH/sctransform) (Hafemeister and Satija, 2019). Clustering for major cell types was performed using Louvain clustering on dimensionally reduced principal components.

Parameters depend on the size of the dataset, and thus must be adjusted based on how many cells are being partitioned for each recursive step. When calculating nearest neighbor graphs, and clustering we set the K parameter to ‘ceiling(0.5*sqrt(N))’. We chose the number of principal components based on the top 15 percentile of calculated improvement of variance explained. For subsets less than 500 cells we used Jackstraw to calculate significant principal components. If neither method succeeded, we chose the first two principal components. This only occurred rarely at late stages where Jackstraw was unable to find significance. We set clustering resolution via a grid search optimizing for maximum average silhouette score (silhouette measures the ratio of intra-cluster distance to inter-cluster distance, where a high score means highly distinct clusters). For stages where we were clustering more than 500 cells a randomized subsample of N cells / 10 was used to calculate the average silhouette score.

Additionally, at each recursive step we output quality metrics and basic plots, such as 1:rest differential expression from the optimal partitioning at each stage and UMAP representations painted by sample metadata (sample ID, cluster number). Our pipeline, saved output as a directory structure matching the tree discovered by this recursive clustering. This tree represents the lower levels of variance of discovered at each tier. At any tier level we are able to extract the cell’s partitioning. Due to the intermixing of patient and cell identity effects at multiple levels of the tree (a fraction of a single patient’s cells might separate out at a high level, but then continue to separate into identifiable cell types, or vice versa), we found the most meaningful levels at the top and bottom of the tree. The clustering tree is useful for understanding the levels of variance in the dataset, but we found it contains too much noise to be easily interpretable. Thus, we later generated a hierarchical clustering of the bottom level clusters based on pairwise differential expression, which is displayed in figures (**Figure 3**: FGID atlas, **Figure 4**: pediCD atlas). See the hierarchical clustering of cell subsets section for details.

#### Cell Type and Subset Annotation from Tiered Clustering

After running the iterative tiered clustering pipeline we manually curated the generated tree of clusters. Specifically, tree generation was reinitiated for the B Cells within the FGID dataset as it had stopped at the first tier on two clusters with < 50 genes differentially expressed, however we could see in this case that there was additional biological stratification based on strong differential expression of *CXCR4* (Wilcoxon; logFC=1.22860917, Bonferroni.p=1.0E-300) *CD69* (Wilcoxon; logFC=1.27527652, Bonferroni.p=2.99E-151), *HMGN1* (Wilcoxon; logFC=-1.1688612, Bonferroni.p=1.62E-227) and *HMGA1* (Wilcoxon; logFC=-1.28838294, Bonferroni.p= 1.06E-209) among others. This formed a clear divide between non-proliferating and proliferating B Cells, further validated by a clear separation within the UMAP based on PCA reduced variable genes within the B cells. We further examined each branching point of the tree to determine its splitting cause, noting splits based on spillover, doublet, and singular patient effects. Splits at higher tiers based on doublets often split again allowing us to recover cells that did not have the dual expression profile. Splits that only had patient splits below (measured by having only clusters of single patients) were manually marked as end clusters, thereby merging all clusters below that split. With these manual steps made, we performed pairwise differential expression to ensure each partitioned subset is distinct from its neighbors.

We annotated these final clusters with four methods attempting to balance descriptiveness, ease of understanding, and ease of name generation: The first method, is generated during the hierarchical tiered clustering by following the path from the end cluster up to the original tier. An example annotation is *T0C0.T1C3.T2C3.T3C5* marking an end cluster that split at tier 1 into cluster 0 and at tier 2 into cluster 3. These annotations do not provide any biological information to the reader, but do provide a unique ID for the end cluster.

Our second method is far more descriptive, where we manually annotate the main reason for each particular split. As several cell types contained readily identifiable and meaningful cell subsets, we utilized curation of literature-based markers to provide further guidance within each cell type (Blériot et al., 2020; Cherrier et al., 2018; Dutertre et al., 2019; Guilliams et al., 2018; Robinette and Colonna, 2016). For example, within Tier 1 T cells, we could identify T cells, NK cells and ILCs; within Tier 1 myeloid cells, monocytes, cDC1, cDC2, macrophages and pDCs; within Tier 1 B cells, germinal center, germinal center dark zone and light zone cells; within Tier 1 endothelial cells, arterioles, capillaries, lymphatics, mural cells and venules; and so forth for other cell types. To illustrate this process for one cluster, upon automated hierarchical tiered clustering of T cells, we identified a cluster that was Tier 0: pediCD, Tier 1: T cells, Tier 2: cytotoxic, Tier 3: IEL_FCER1G_NKG7_TYROBP_CD160_AREG. Upon inspection of CD3 genes (*CD247, CD3D,* etc.), TCR genes (*TRAC, TRBC1*, etc.), and NK cell genes (*NCAM1, NCR1*), it became readily apparent these cells were NK cells (**Supplemental Figure 6**). This still follows the original ranking of variation as found by the hierarchical tiered clustering, while also providing biological interpretation, as an example: *CD.Mloid.macrophage_chemokine.S100A8_S100A9_CXCL9_CXCL10_TNF_inflamonocyte*.

This method of annotation was particularly useful during analysis as we were immediately able to see how early or late two clusters had split from each other, as well as seeing a number of the subset defining genes. Unfortunately, as is apparent this method also produces extremely long names that are difficult to display and to which to refer. It is also a highly manual process, and difficult to reproduce precisely. To better present our findings and aid others in reproducing our results, our third method automates this annotation. This method is performed by taking each major cell type, which in our case matched the tier one splits, and performing 1:rest differential expression testing (Wilcoxon; adj.p<0.05, only.pos=True) within each major cell type. We then ranked the genes based on the product of ‘-log(Bonferroni.p)’, ‘avg_logFC’, and ‘pct.exp.1/pct.exp.rest’ and took the top 5, forming a name like *CD.Mloid_CCL3_CCL4_CCL3L3_TNF_TNFAIP6*. This scheme, again was useful, but did not quite meet our demands of recognizability and brevity.

#### Rank-Score gene selection naming function

To account for genes that might be highly expressed in just a few cells we ranked the marker genes by a score combining their significance, the fold change in expression, and fold change of percent gene positive cells in the subset versus the percent of gene positive cells outside the subset. The collected metrics were multiplied together to provide a single score by which the genes were ranked: **(-log(sig+1) * avg_logFC * (pct.in / pct.out))**. For most subsets we selected the top 2 of these marker genes. For T/NK/ILC cells and myeloid cells we occasionally chose a slightly lower ranking gene from the top 10 if it was well supported and recognized by the literature. Thus, for T and Myeloid cells we adjusted these names to a finer degree of specificity by visualizing the expression profiles of each subset with a dotplot of canonical marker genes based off of current literature, and limiting to the top 2 genes based off our method 3 rankings and the dotplot of canonical markers, thereby producing the fourth and final annotations in the form: *CD.Mono.CXCL10.TNF.* Due to the limited nature of current characterization of stromal and epithelial cells we were unable to match the same degree of specificity as the T and Myeloid cells, however we did where possible adjust from the major cell type, to the most specific that we could be confident of. For instance, adjusting “Epith” to “Goblet” based on marker expression of *TFF3* and *MUC13*.

#### Hierarchical Clustering of Subsets on Unified Gene Space and Removal of Doublets

At this point we had generated a hierarchical representation of the datasets from the top down showing the splits of highest variation at every level. By necessity that means that each level is controlled by and represents different selections of genes, which may have no relation to the genes selected in another branch. To understand the relations of cell subsets and compare across cell type we needed a unified set of genes. For each dataset (FGID and pediCD) we performed pairwise differential expression (Wilcoxon; Bonferroni.p<0.01, max.cells.per.ident=500) and selected the top 50 most significant genes from each test. Gene lists were merged as a union, finding 4445 unique genes for FGID and 1844 unique genes for pediCD that best differentiate the subsets. Subset centers were calculated from these selected genes as the median expression of cells grouped by subset. The resulting table was then hierarchically clustered using correlation distance and complete linkage. Clustering was performed in R using the pvclust package (https://github.com/cran/pvclust).

The resulting tree shows from the bottom up the relationships between cell subsets, and allows cell subsets that were potentially misclassified at a high split in hierarchical tiered clustering to find their biological neighboring subsets. As previously mentioned within our description of hierarchical tiered clustering we did not find any end cluster subsets that met our thresholds for merging. This does not mean that we did not observe shuffling from the initial tiered splits. While overall there was good agreement between the two methods, we noted subsets jumping between major cell types as defined by the first splits of our tiered clustering. We identified the majority of these jumping subsets as doublet clusters by exploring their differential gene results at multiple levels of the tiered clustering tree. We removed these doublet subsets and others based on flipping expression programs at different tiers. For instance, looking like T cells expressing *TRAC*, *IL7R* within an epithelial cluster, then at the next tier having significantly higher expression of *KRT18* and *PIGR* than neighboring clusters. After removing doublets, we recalculated subset distances and dimensional reductions, as presented in the main figures.

#### FGID Atlas Generation and Curation

Within B cells, we identified a strong division between non-cycling and cycling B cells, with those found in the cycling compartment readily identifiable by germinal center markers and further dark zone (*AICDA*) and light zone (*CD83*) genes resulting in FG.B/DZ.AICDA.IGKC and FG.B/LZ.CD74.CD83 clusters (**Figure 3d**) (Victora et al., 2010).

Within myeloid cells, we identified, and confirmed using extensive inspection of literature curated markers, cell subsets corresponding to monocytes (*CD14, FCGR3A, FCN1, S100A8, S100A9*, etc.), macrophages (*CSF1R, MERTK, MAF, C1QA*, etc.), cDC1 (*CLEC9A, XCR1, BATF3*), cDC2 (*FCER1A, CLEC10A, CD1C, IRF4* etc.), and pDCs (*IL3RA, LILRA4, IRF7*) (**Figure 3d; Supplemental Table 6**) (Blériot et al., 2020; Dutertre et al., 2019; Guilliams et al., 2018). We highlight selected cell states including a migratory dendritic cell state (FG.DC.CCR7.FSCN1), extensive cDC2 heterogeneity relative to cDC1 heterogeneity, and a main distinction between macrophage clusters expressing *C1Q\**, *MMP\**, *APOE, CD68* and *PTGDS* (FG.Mac.C1QB.SEPP1, FG.Mac.APOE.PTGDS) and a series of clusters expressing various chemokines including *CCL3, CXCL3,*and *CXCL8* (FG.Mac.CCL3.HES1, FG.Mac.CXCL3.CXCL8, FG.Mac.CXCL8.IL1B).

Within T cells, we followed a similar approach as utilized for Myeloid cells and identified principal cell subsets of T cells (joint expression of *CD247, CD3D, CD3E, CD3G* with *TRAC, TRBC1, TRBC2*, or *TRGC1, TRGC2* and *TRDC*), and a combined cluster of cytotoxic cells (FG.T/NK/ILC.GNLY.TYROBP) likely including T cells, NK cells (lower expression of TCR-complex genes with *NCAM1*, *NCR1* and *TYROBP*), and some ILCs (*KIT, NCR2, RORC* and low expression of CD3-complex genes) (**Figure 3d, Supplemental Table 6**) (Cherrier et al., 2018; Robinette and Colonna, 2016). We note that the numerical majority of CD4 T cells (FG.T/NK/ILC.MAF.RPS26) and CD8 T cells (FG.T/NK/ILC.CCR7.SELL) expressed *SELL* and *CCR7* thus identifying them as naïve T cells. However, regardless of clusters expressing *CD4* (FG.T.GZMK.GZMA) or *CD8A/CD8B* (FG.T.GZMK.IFNG, FG.T.GZMK.CRTAM, etc.), most activated T cells were characterized by expression of granzymes (Sallusto et al., 1999).

Within epithelial cells, most cells expressed high levels of *OLFM4*, identifying them as crypt-localized cells (Moor et al., 2018). We readily identified subsets of stem cells (*LGR5*), proliferating cells (*TOP2A*), goblet cells (*SPINK4*, *ZG16*, various *MUCs*), enteroendocrine cells (*SCG3, ISL1*), Paneth cells (*ITLN2, PRSS2, LYZ*), tuft cells (*GNG13, SH2D6, TRPM5*) and enterocytes (*APOC3, APOA1, FABP6,* etc.) (**Figure 3d, Supplemental Table 6**) (Barker et al., 2007; van der Flier and Clevers, 2009).

Within endothelial cells, we readily identified vascular and lymphatic endothelial cells (*LYVE1, PROX1*), with the vascular cells able to be further identified as capillaries (*CA4*) or venular endothelial cells (*ACKR1, MADCAM1*) (Brulois et al., 2020). We also identified a subset of cells (FG.Endth/Peri.FRZB.NOTCH3) expressing high levels of *FRZB* and *NOTCH3*, which, rather than being arterioles, likely represent arteriole-associated pericytes or smooth muscle cells given the absence of *EFNB2, SOX17, BMX,* and *HEY1,* and the presence of *ACTA2* and *MYL9*, as cluster-defining genes (**Figure 3d, Supplemental Table 6**) (Travaglini et al., 2020; Whitsett et al., 2019). We highlight that the FG.Endth/Ven.ACKR1.MADCAM1 cluster is characterized by expression of markers for postcapillary venules specialized in leukocyte recruitment (Thiriot et al., 2017).

Within fibroblasts, we identified principal subsets characterized by their structural roles (*COL3A1, ADAMDEC1, FBLN1, LUM,* etc.), myofibroblasts (*MYH11, ACTA2, ACTG2,* etc.), and organization of lymphoid cells (*CCL19, CCL21* etc.) (**Figure 3d, Supplemental Table 6**) (Buechler et al., 2021; Davidson et al., 2021). Within the lymphoid-organizing fibroblasts, we draw attention to the FG.Fibro.C3.FDCSP, FG.Fibro.CCL19.C3, and FG.Fibro,CCL21.CCL19 subsets, which appear to have some characteristics of follicular dendritic cells and variable expression of *CCL19/CCL21* (T-cell or migratory dendritic cell chemoattractants) and *CXCL13* (B-cell chemoattractant) (Das et al., 2017; Heesters et al., 2013). We also identified a separate Tier 1 cluster of glial cells characterized by *CRYAB* and *CLU*. Intriguingly within the glial cell Tier 1 cluster, we then recovered a cell subset expressing *FDCSP, CXCL13,* and *CR2,* a key complement receptor which allows for complement-bound antigens to be recycled and presented by follicular dendritic cells (Das et al., 2017; Heesters et al., 2013). This highlights the power of iterative tiered clustering to recover discrete cell states that may, through the process of traditional clustering, not be fully resolved. Furthermore, the presence of these discrete cell clusters within larger parent cell clusters will alter the gene expression signatures of the higher-level cell types. For example, the FG.Glial/fDC.FDCSP.CXCL13 in the hierarchical cluster tree then assorts within the lymphoid-organizing stromal cells.

The mast cells recovered did not further sub-cluster in an automated fashion, and were largely marked by *TPSB2* and *TPSAB1* (>97%), with minimal *CMA1* (<20%) expressing cells, suggesting they are largely classical MC-T cells in FGID intestine (Dwyer et al., 2021).

We identified four Tier 1 clusters for plasma cells, which are characterized by their strong expression of IGH* immunoglobulin heavy-chain genes together with either an IGK* (kappa light chain) or IGL* (lambda light chain) genes (Cyster and Allen, 2019; James et al., 2020). This resolved IgA IgK plasma cells, IgA IgL plasma cells, IgM plasma cells, and IgG plasma cells. Iterative tiered clustering identified further heterogeneity within all clusters of IgA and IgG plasma cells, though given the 3’-bias of this dataset, we note that a principled investigation of these clusters would ideally use 5’ sequencing with targeted VDJ amplification.

Together, our treatment-naïve cell atlas from 13 FGID patients captures 138 cell clusters from a non-inflammatory state of pediatric ileum which we annotated and named in a principled fashion.

We note that p044 was overrepresented with more terminally differentiated epithelial cells, likely from incomplete EDTA separation, and thus omit the p044 unique cell clusters from further analyses of composition.

#### pediCD Atlas Generation and Curation

Within B cells, we also identified a strong division between non-cycling and cycling B cells, with those found in the cycling compartment readily identifiable by germinal center markers and further dark zone (*AICDA*) and light zone (*CD83*) genes, as in FGID (Victora et al., 2010). Within cells expressing germinal centers markers, a highly-proliferative branch including clusters such as CD.B/LZ.CCL22.NPW, CD.B/GC.MKI67.RRM2, and CD.B/DZ.HIST1H1B.MKI67 emerged (**Figure 4d**). The CD.B/LZ.CCL22.NPW was characterized by high levels of *MYC*, which has been shown to allow for further rounds of germinal center affinity maturation (Dominguez-Sola et al., 2012). More numerous B cell clusters included ones characterized by expression of *GPR183*, such as CD.B.CD69.GPR183 (also expressing *IGHG1*) and CD.B.RPS29.RPS21. GPR183 has been shown to regulate the positioning of B cells in lymphoid tissues (Pereira et al., 2009).

Within myeloid cells, we identified, and confirmed using the same extensive inspection of literature curated markers as in FGID, cell subsets corresponding to monocytes (*CD14, FCGR3A, FCN1, S100A8, S100A9*, etc.), macrophages (*CSF1R, MERTK, MAF, C1QA*, etc.), cDC1 (*CLEC9A, XCR1, BATF3*), cDC2 (*FCER1A, CLEC10A, CD1C, IRF4* etc.), and pDCs (*IL3RA, LILRA4, IRF7*) (**Figure 4d; Supplemental Figure 6, Supplemental Table 9**) (Blériot et al., 2020; Dutertre et al., 2019; Guilliams et al., 2018). We highlight selected cell states including a migratory dendritic cell state (CD.DC.CCR7.FSCN1), extensive cDC2 heterogeneity relative to cDC1 heterogeneity, and a main distinction between macrophages expressing *C1Q\**, *MMP\**, *APOE, CD68* and *PTGDS*, (CD.Mac.APOE.PTGDS) and a series of clusters expressing various chemokines including *CXCL2, CXCL3,* and *CXCL8* (CD.Mac.SEPP1.CXCL3, CD.Mono.CXCL3.FCN1, CD.Mono.CXCL10.TNF). Several of the end cell clusters initially clustering with macrophages also expressed monocyte markers (*S100A8, S100A9*), and expressed detectable, but lower levels of *MERTK* or *AXL* relative to *bona fide* macrophages, potentially indicative of the early stages of the trajectory of monocyte-to-macrophage differentiation (Blériot et al., 2020; Dutertre et al., 2019; Guilliams et al., 2018). We also noted a substantial expansion of clusters characterized by expression of *CXCL9, CXCL10,* and *STAT1,* canonical interferon-stimulated genes, observed in clusters such as CD.Mono/Mac.CXCL10.FCN1 (Ziegler et al., 2021, 2020). Moreover, we identified a cluster of inflammatory monocytes, CD.Mono.S100A8.S100A9, characterized by both *CD14* and *FCGR3A* expression.

Within T cells, we followed a similar approach as utilized for FGID T cells and identified cell subsets of T cells (joint expression of *CD247, CD3D, CD3E, CD3G* with *TRAC, TRBC1, TRBC2*, or *TRGC1, TRGC2* and *TRDC*), but in pediCD also identified several discrete clusters of NK cells (lower expression of TCR-complex genes with *FCGR3A* or *NCAM1*, *NCR1* and *TYROBP*), and ILCs (*KIT, NCR2, RORC* and low expression of CD3-complex genes) (**Figure 4d, Supplemental Figure 6, Supplemental Table 9**) (Cherrier et al., 2018; Robinette and Colonna, 2016). We note that T cells and NK cells with a shared expression of *GNLY*, *GZMB* and other cytotoxic effector genes cluster almost indistinguishably from each other through iterative tiered clustering and visualization of the hierarchical tree, but that careful inspection of literature-curated markers helped resolve NK cells (CD.NK.CCL3.CD160; CD.NK.GNLY.GZMB) from *CD8A*/*CD8B* T cells (CD.T.GNLY.GZMH; CD.T.GNLY.CTSW) (**Figure 4d, Supplemental Figure 6, Supplemental Table 9**) (Cherrier et al., 2018; Robinette and Colonna, 2016). One of the specific challenges in distinguishing between T cells and NK cells in scRNA-seq data is that NK cells can express several CD3-complex genes, particularly *CD247*, as well as detectable aligned reads for *TRDC* or *TRBC1* and *TRBC2*, and thus lower-resolution clustering approaches or datasets with lower cell numbers may miss these important distinctions (Björklund et al., 2016; Renoux et al., 2015). NK cell clusters also expressed the highest levels of *TYROBP*, which encodes DAP12 and mediates signaling downstream from many NK receptors (French et al., 2006; Lanier, 2001; Lanier et al., 1998). ILC clusters such as CD.ILC.LST1.AREG or CD.ILC.IL22.KIT were characterized by an apparent ILC3 phenotype, with expression of *KIT*, *RORC* and *IL22*, though they also expressed detectable transcripts of *GATA3* in the same clusters (Cherrier et al., 2018; Robinette and Colonna, 2016). We detected several clusters expressing *CD4* and lacking *CD8A/CD8B*, including regulatory T cells (CD.T.TNFRSF18.FOXP3), and *MAF*- and *CCR6*-expressing helper T cells (CD.T.MAF.CTLA4). Perhaps most strikingly, we resolved multiple subsets of proliferating lymphocytes, including regulatory T cells (CD.T.MKI67.FOXP3), *IFNG*-expressing T cells (CD.T.MKI67.IFNG), and NK cells (CD.NK.MKI67.GZMA).

Within epithelial cells we identified substantial heterogeneity in CD. Most cells expressed high levels of *OLFM4*, identifying them as crypt-localized cells (Moor et al., 2018). We readily identified subsets of stem cells (*LGR5*), proliferating cells (*TOP2A*), goblet cells (*SPINK4*, *ZG16*, various *MUCs*), enteroendocrine cells (*SCG3, ISL1*), Paneth cells (*ITLN2, PRSS2, LYZ*), tuft cells (*GNG13, SH2D6, TRPM5*) and enterocytes (*APOC3, APOA1, FABP6,* etc.) (**Figure 4d, Supplemental Table 9**) (Barker et al., 2007; van der Flier and Clevers, 2009). Amongst several clusters characterized by *CCL25* and *OLFM4* expression, we identified a subset marked by *LGR5* expression, characteristic of intestinal stem cells (CD.EpithStem.LINC00176.RPS4YA1) (Barker et al., 2007). We identified several subsets expressing *CD24*, indicative of crypt localization, with expression of *REG1B* (CD.Secretory.GSTA1.REG1B; CD.Secretory.REG1B.REG1A) (Moor et al., 2018). We also identified early enterocyte cluster CD.EC.ANPEP.DUOX2, characterized by *FABP4* and *ALDOB* and expressing *DUOX2 and MUC1*. We resolved several clusters of enteroendocrine cells, including CD.Enteroendocrine.TFPI2.TPH1 and CD.Enteroendocrine.NEUROG3.MLN. We also found two clusters we labeled as M cells based on expression of *SPIB* (CD.Mcell.CCL23.SPIB; CD.MCell.CSRP2.SPIB) (Beumer et al., 2020; Mabbott et al., 2013). Paneth cells did not further sub-cluster despite forming an independent Tier 1 cluster (CD.Epith.Paneth). Most strikingly, we identified a diversity of goblet cells recovered across multiple patients including CD.Goblet.HES6.COLCA2 expressing *REG4* and *LGALS9*, and CD.Goblet.TFF1.TPSG1 expressing *TFF1* and *ITLN1*, amongst others. We also identified a cluster of Tuft cells: CD.EC.GNAT3.TRPM5.

Within endothelial cells, we also readily identified vascular and lymphatic endothelial cells (*LYVE1, PROX1*), with the vascular cells able to be further identified as capillaries (*CA4*) or venular endothelial cells (*ACKR1, MADCAM1*) (**Figure 4d, Supplemental Table 9**). We also identified a subset of cells (CD.Endth/Mural.HIGD1B.NDUFA4L2) expressing high levels of *FRZB* and *NOTCH3*, which, rather than being arterioles, likely represent arteriole-associated pericytes or smooth muscle cells given the absence of *EFNB2, SOX17, BMX,* and *HEY1,* and the presence of *ACTA2* and *MYL9*, as cluster-defining genes. In pediCD, we also identified a cluster of arteriolar endothelial cells, CD.Endth/Art.SEMA3G.SSUH2, identified by expression of *HEY1, EFNB2,* and *SOX17*. We also highlight that the endothelial venules characterized by expression of markers for postcapillary venules specialized in leukocyte recruitment, such as CD.Endth/Ven.ADGRG6.ACKR1 and CD.Endth/Ven.POSTN.ACKR1, exhibited greater diversity than in FGID with multiple end cell clusters identified (Thiriot et al., 2017).

Within fibroblasts, we identified principal subsets characterized by their structural roles (*COL3A1, ADAMDEC1, FBLN1, LUM,* etc.), myofibroblasts (*MYH11, ACTA2, ACTG2,* etc.), and organization of lymphoid cells (*CCL19, CCL21* etc.) (**Figure 4d**) (Buechler et al., 2021; Davidson et al., 2021). The principal hierarchy in fibroblasts in pediCD was between *FRZB-*, *EDRNB*- and *F3*-expressing subsets such as CD.Fibro.LY6H.PAPPA2 and CD.Fibro.AGT.F3, which were also enriched for *CTGF* and *MMP1* expression, and *ADAMDEC1*-expressing fibroblasts, which were enriched for several chemokines such as *CXCL12,* and in some specific clusters *CXCL6, CXCL1, CCL11,* and other chemokines. Amongst three fibroblast subsets marked by *C3* expression, we identified follicular dendritic cells (CD.Fibro/fDC.FCSP.CXCL13), along with fibroblasts expressing *CCL21*, *CCL19*, and the interferon-stimulated chemokines *CXCL9* and *CXCL10* (CD.Fibro.CCL21.CCL19; CD.Fibro.TNFSF11.CD24) (Das et al., 2017;

Heesters et al., 2013). Distinct from the FGID atlas, within the pediCD atlas, glial cells clustered within fibroblasts, but were also marked by *S100B, PLP1* and *SPP1* expression.

The mast cells recovered in pediCD did further sub-cluster in an automated fashion, were largely marked by *TPSB2* (>90%), with minimal *CMA1* (<16%) expressing cells, suggesting they are largely classical MC-T cells in pediCD intestine (**Figure 4d**) (Dwyer et al., 2021). Intriguingly, some subsets (CD.Mstcl.AREG.ADCYAP1) were enriched for *IL13*-expression. We also detected a small cluster of proliferating mast cells from several patients (CD.Mstcl.CDK1.KIAA0101).

We also identified four Tier 1 clusters for plasma cells, which are characterized by their strong expression of IGH* immunoglobulin heavy-chain genes together with either a IGK* (kappa light chain) or IGL* (lambda light chain) genes. This resolved IgA IgK plasma cells, IgA IgL plasma cells, IgM plasma cells, and IgG plasma cells. Iterative tiered clustering identified further heterogeneity within all clusters of IgA plasma cells, though given the 3’-bias of this dataset, we note that a principled investigation of these clusters would ideally use 5’ sequencing with targeted VDJ amplification.

Together, our treatment-naïve cell atlas from 14 pediCD patients captures 305 cell clusters from an inflammatory state of the pediatric ileum suggesting an increase in the number and diversity of cell states present in the intestine during overt inflammatory disease.

#### Association of Cell Subsets to anti-TNF Response

Compositional differences are an important metric for understanding the baseline differences that prognose a patent’s response to treatment. We measure these differences with proportional enrichment of particular cell subsets within each patient, and finding the significantly reproducible enrichments across disease. As an extreme toy example, we might find that subset A cells comprise as much 80% of cells sampled in one condition whereas they might only comprise 30% in a different condition. This type of compositional analysis is highly affected by the number and choice of subsets included, and the sampling depth per patient (how may cells are collected). The first factor is controlled by the confidence in our clustering and using computationally optimized parameters. We further control this factor by limiting analysis of compositional shifts of cell states to within major cell types. This isolates the chance of error from affecting the entire analysis and allows us to gain a more direct biological insight of the rise and fall of particular cell states in the context of similar subsets. We control the second factor of sampling depth differences by computing a normalized cell count score per patient of the form (ncells in subset / ncells in patient’s major cell type) * 1e6. This score provides us with the number of cells expected per million.

#### Mann-Whitney tests

We input our cells per million score into a two-sample Wilcoxon test in base R, which is equivalent to the Mann-Whitney rank score test. We set a significance threshold of p_value < 0.05. We made 5 different pairwise comparisons (FGID vs FR, FGID vs PR, NOA vs FR, NOA vs PR, FR vs PR). Comparisons between FGID and pediCD groups were determined by finding maximum correspondence between the disease conditions for each subset.

#### Fisher’s Exact tests

A similar compositional analysis to that done with the Mann-Whitney was performed with a Fisher’s Exact test. We input for each subset the number of cells for that subset against the number of cells not of that subset within the major cell type split on rows by pairwise comparisons (NOA vs FR, NOA vs PR, FR vs PR). We computed FDR correction of p_values at major cell type and entire dataset levels and found significance subsets at both levels. But, most interestingly in comparing the two tests we found that the Mann-Whitney discovered as significant (pval < 0.05) the portion of cell subsets with largest effect sizes. Understanding our limited patient number at these within pediCD comparisons and wanting to only report results most likely to be reproducible biology, we determined to only follow those subsets reported as significant within both Mann-Whitney and Fisher’s exact tests.

#### Discrete cell cluster changes across the pediCD clinical severity and response spectrum

When comparing FR/PRs to NOAs, two subsets with significantly increased frequency in FR/PR patients amongst T cells, NK cells, and ILCs were identified. These were CD.NK.MKI67.GZMA and CD.T.MKI67.IL22 (**Figure 5c; Supplemental Figure 7b; Supplemental Table 15**). Beyond the strong proliferation signature, CD.NK.MKI67.GZMA were enriched for genes such as *GNLY, CCL3, KLRD1, IL2RB* and *EOMES*, and CD.T.MKI67.IL22 were enriched for *IFNG, CCL20, IL22, IL26, CD40LG* and *ITGAE*. This indicates that with increasing pediCD clinical severity, there is increasing local proliferation of cytotoxic NK cells and proliferation of tissue-resident T cells with the capacity to express anti-microbial and tissue-reparative cytokines, and molecules to interface with antigen-presenting cells and B cells. Alongside this increase, there was a significant decrease amongst fibroblasts of CD.Fibro.CCL19.IRF7, and amongst epithelial cells of CD.EC.SLC28A2.GSTA2 clusters in the FR/PR patients compared to NOA (**Supplemental Figure 7b**). The CD.Fibro.CCL19.IRF7 were enriched for *CCL19, CCL11, CXCL1, CCL2,* and very specifically for *OAS1* and *IRF7*. The CD.EC.SLC28A2.GSTA2 cluster was characterized by its two namesake markers, involved in purine transport and glutathione metabolism (Moor et al., 2018).

We also detected significant decreases in FRs relative to NOAs in certain cell types, particularly within Epithelial cells including CD.EpithStem.LINC00176.RPS4Y1, CD.MCell.CSRP2.SPIB, CD.EC.FABP6.PLCG2, and CD.EC.FABP1.ADIRF (**Supplemental Figure 7c; Supplemental Table 15**). We note that the relative decrease in M cells is in stark contrast to the “ectopic” M-like cells that were detected in adult ulcerative colitis (Smillie et al., 2019).

We next focused on those cell subsets that were significantly changed only between PRs and NOAs (**Figure 5c; Supplemental Figure 7d; Supplemental Table 12**). Here we note several distinct clusters within the lymphocyte cell type, including increases in the PR patients compared to NOA patients of CD.T.MKI67.IFNG, CD.T.MKI67.FOXP3, CD.T.GNLY.CSF2, and CD.NK.GNLY.FCER1G. The two MKI67 clusters again highlighted an increase in proliferative cells, specifically cells enriched for *IFNG, GNLY, HOPX, ITGAE* and *IL26* (CD.T.MKI67.IFNG), and *IL2RA, BATF, CTLA4, TNFRSF1B, CXCR3,* and *FOXP3* (CD.T.MKI67.FOXP3), the latter of which may be indicative of proliferating regulatory T cells. The two GNLY clusters emphasized cytotoxicity as they were both enriched for *GNLY, GZMB, GZMA, PRF1* and more specifically for *IFNG, CXCR6,* and *CSF2* (CD.T.GNLY.CSF2), or *AREG, TYROBP,* and *KLRF1* (CD.NK.GNLY.FCER1G). Amongst myeloid cells, there was an increase in CD.Mac.CXCL3.APOC1, CD.Mono/Mac.CXCL10.FCN1, and CD.Mono.FCN1.S100A4 in PR versus NOA. The CD.Mac.CXCL3.APOC1 cluster was enriched for a variety of chemokines including *CCL3, CCL4, CXCL3, CXCL2, CXCL1, CCL20,* and *CCL8*. It was also enriched for *TNF* and *IL1B*. The CD.Mono/Mac.CXCL10.FCN1 cluster was enriched for *CXCL9, CXCL10, CXCL11, GBP1, GBP2, GBP4, GBP5,* suggestive of activation by IFN, and more specifically Type II IFNγ, based on the GBP gene cluster (Ziegler et al., 2020). CD.Mono.FCN1.S100A4 was characterized by *S100A4, S100A6,* and *FCN1* expression. These two immune clusters were paralleled by increases in certain clusters within endothelial cells in PR versus NOA patients (CD.Endth/Ven.LAMP3.LIPG) and epithelial cells (CD.Goblet.TFF1.TPSG1).

Several clusters of cells were decreased in PR versus NOA, including CD.T.LAG3.BATF, CD.T.IFI44L.PTGER4, and CD.T.IFI6.IRF7 amongst lymphocytes (**Supplemental Figure 7d**) (Roncarolo et al., 2018). Amongst myeloid cells, CD.cDC2.CLEC10A.FCGR2B were decreased, and amongst fibroblasts CD.Fibro.IFI6.IFI44L were decreased. In epithelial cells, CD.Tuft.GNAT3.TRPM5 cells were decreased. Alongside the decrease in Tuft cells amongst epithelial cells, two more clusters closely related to the aforementioned CD.EC.GSTA2.SLC28A3 cluster, also marked by *GSTA2* expression, were significantly decreased (CD.EC.GSTA2.CES3, and CD.EC.GSTA2.TMPRSS15).

We assessed the compositional differences between FRs and PRs and only identified one cell cluster which was significantly increased in PRs: CD.B/DZ.HIST1H1B.MKI67, which are proliferating dark zone B cells. CD.T.EGR1.TNF T cells were significantly decreased in PR versus FR (**Supplemental Figure 7e; Supplemental Table 15**). These data suggest that at the time of diagnosis of pediCD, there are a series of changes in multiple cell types that encapsulate the distinctions between NOA and FR or PR patients.

We provide an example of specific tiers, subclusters, and modules of co-expressed genes with proliferating T cells as a case study. We compare our approach of iterative clustering within a cell state (such as proliferating T cells), and then testing for significant differences in the composition between disease severity groups (**Supplemental Figure 8a**) vs. performing differential expression within the proliferating T cell state between disease severity groups (**Supplemental Figure 8b**). Importantly, differential expression at this level fails to recover the critical substructure present in our dataset (**Supplemental Figure 8a,b**). This is further validated through the additional use of topic modeling: an orthogonal method to clustering for cell program identification (**Supplemental Figure 8c**) (Bielecki et al., 2021). For example, while *GZMA* alone is not differentially expressed, the cell subset CD.NK.MKI67.GZMA proliferating cytotoxic NK cells, identified by ARBOL and Topic modeling, is significantly increased by compositional (i.e. percentage of the parent cell subset, as typically performed in flow cytometry) analysis with increasing disease severity (**Figure 5c**). This highlights the need to examine how co-variation in multiple genes defines cell clusters beyond traditional differential expression at a coarser level of clustering(Shalek et al., 2014, 2013).

#### Principal component analysis of cell frequencies and correlation to clinical metadata

Cell frequencies were calculated per patient for cell subsets (i.e. end clusters) within parent cell types and cell subsets (i.e. end clusters) within all cells as CPM = ((count/sum(count)) * 1e6.

Principal component analysis (PCA) was performed on the resulting patient x CPM matrices using the R package *stats*::prcomp(., scale=TRUE). Variance explained per PC was calculated as std^2/sum(std^2). PCA loadings per patient and per cell subset were extracted from the prcomp() result. PCA1 and PCA2 from the total PCA x patient and from each celltype’s PCA x patient matrix were correlated with clinical metadata using Spearman rank correlation as calculated by the R package *stats*::cor.test(., method=’spearman’). P values were recorded from the cor.test() call, and FDR was calculated using R’s *fdrtool*::fdrtool(p.values, statistic=”pvalue”). For combined celltype PCA’s, patient x CPM tables were concatenated before PCA.

#### Finding Corresponding Cell Subsets Between Diseases

Separating the data on disease condition into two datasets was important as it allowed us to isolate the axis of cell identity within each disease and be confident in the homogeneity of each subset. But does present the problem of how to make comparisons across the disease condition:

#### 1^st^ Approach – KNN classifier

Our first attempt to find corresponding clusters followed the methods of *Tasic et al. 2018*. We used the best differentiating genes sets created for our unified gene space clustering to as the mapping space for a nearest-neighbor classifier. For each cell within the disease condition, we could map it to the nearest cell subset within the other disease condition. As a trial run, we created this gene space for each major cell type of the FGID disease condition and performed 5-fold cross-validation. Unfortunately, we could only achieve accuracies of up to 55%.

#### Rethinking approach

From this trial run we realized that we needed a more automated system to choose genes as the most significantly differentially expressed genes did not create enough separation between cluster centers to effectively classify new cells. We chose to use an RF classifier as it allowed us to train for the optimal selection of genes, required little to no preparation of data, and provided probabilities of each cell being predicted to each class. These probabilities for each class proved particularly useful do to our second realization. Because the number of subsets differs between disease conditions, we could not assume that there was a one-to-one relationship between conditions. Nor could we assume that the many-to-one relationships were unidirectional with one base subset splitting into many states only from FGID toward CD. A single classifier would not allow us to distinguish between these many types of relationships. However, we realized that by creating a classifier for both directions (FGID to pediCD & pediCD to FGID) we could take advantage of the difference in confidence between the two classifications to discover the direction and type of relationship. In a perfect world of 1-to-1 relationships, we would expect all cells of subset A in condition X to match with 100% confidence to subset B in condition Y. In that particular case the summed probability equal 2 and there would be zero difference in confidence of one classifier to its matching classifier. In our imperfect world we might instead see 90% of cells of subset A in condition X to matching with > 85% confidence to subset B in condition Y, and only 30% of subset B in condition Y matching with > 85% confidence. From this discrepancy we can infer that subset A may be a cell state in condition X that is layered on top of a base state B in condition Y. Low confidence in both directions indicates subsets unique to a particular condition.

#### 2^nd^ Approach – Training Random Forest Model

We trained these models in scikit-learn with 5-fold cross validation and params: min_samples_leaf=1, oob_score=True, criterion=“gini”, max_depth=200, n_estimators=700, max_features=“sqrt” The training set (but not the test set) was sampled with replacement such that all classes contained as many samples as the maximum proportioned class. This up-sampling procedure provided the largest gain to our test accuracy, sensitivity, and specificity scores, increasing accuracy ∼7-10% across each cell type.

We trained random forest classifiers for each cell type in each disease condition using SciKit-Learn v0.22.2, with the intent to classify each cell to the subset in the opposed dataset the cell is most similar to (Pedregosa et al., 2011). For each cell type we optimized a classifier for accuracy using grid-based search tuning number of trees, depth, number of features, criterion, and min samples per leaf with 5-fold cross validation for each set of tuning parameters. We never observed full overfitting where the accuracy on test folds began to drop with increased size of model, but we did quickly find diminishing returns as we increased model size. For simplicity and because optimal tuning parameters were robust to overfitting, we chose to use the same largest model parameters for all models (number of trees = 500, depth = 200, number of features = sqrt, criterion = gini, min samples per leaf = 1, oob_score=True) (Pedregosa et al., 2011). Our initial training rounds found accuracies in the mid 60%. A definite increase from the NN classifier, but not high enough for us to be confident in the results. The main issue we eventually determined to be the uneven class distributions (far more cells in subset A than subset B). This caused the smaller subsets to be under trained. To compensate we up-sampled with replacement each subset within the training fold to contain at least the 75th quantile number of cells. This single change improved accuracy on the unmodified test fold the most, on average providing a ∼7-10% improvement of accuracy, precision and recall across each cell type, and provided accuracies ranging from high 70 to low 90 percent per major cell type.

#### Applying Random Forest Model

Satisfied with the validation results within each disease condition, we ran the random forest model across the disease conditions. We trained each random forest model with optimized parameters on all folds of its dataset, then proceeded to get probability predictions for each cell from the disease condition to the trained disease condition. With these class probabilities per cell we could aggregate for each disease condition by taking the mean class probabilities for each group, leaving us with 2 *n* by *m* table where *n* equals the number of subset groups and *m* equals the number of subset classes in the opposing disease condition. Using the mean probabilities for the group allowed more information from the cell level to rise to the aggregated levels than using the individual class prediction alone (computed as the class with max confidence of cell membership). These tables also provide confidences to all classes which is important for understanding the transverse confidence in both directions.

It is especially important to understand the many-to-one relationships between disease conditions and find where a base cell state becomes layered in additional expression profiles, as these are the exact cases where we can infer the underlying signaling patterns that diversify or concentrate cell state profiles. In diverse splitting of a subset across disease we can start to understand the heterogeneity of patient response to treatment as it becomes clear which particular cell profiles are correlated with strong and poor response. To gain insight to these changes, we care about where there is strong confidence in both directions and where there is strong confidence in only one direction. The simplest method to calculate these is to separately take the sum of the pairwise prediction confidences and the difference. We call the sum of confidences the correspondence of a subset, and the difference the bias.

#### Visualizing Correspondence and Bias

We plot these metrics on a dot plot where each possible connection is laid out on a grid. For each dot we set the size to match the correspondence, and color the dot based on the bias, such that a perfect match would appear as a large white circle. A more unidirectional match would be tinted darker in the color matching the disease condition with more confidence. Matches with more bias tend to indicate a subset matching a base cell state but also expressing some additional gene modules. To aid the human eye on picking up the major patterns we filter to only show the top 10% highest correspondences. This parameter was chosen after looking at the distribution of correspondence scores and selecting the majority of the right tail of the distribution. It keeps the strongest matches in both ways and keeps the strongest in highly biased matches. To also aid the human eye we perform a hierarchical clustering using cosine distance and complete linkage on the prediction confidences and compute an optimal ordering based on the cosine distances using the “cba” package in R: https://cran.r-project.org/web/packages/cba/index.html. This allows us to sort subsets on the rows and columns such that subsets that get predicted similarly are next to each other. From this visualization we are able to easily discern which are the subsets FGID that split into many phenotypes within pediCD from high correspondence and bias, which subsets don’t change phenotype much at all based on high correspondence and low bias, and which are the subsets are potentially unique to a disease condition based on very low correspondence and bias.

#### Random Forest classifiers relative to joint clustering and integration approaches

While integration methods continue to improve (Hao et al., 2021; Hie et al., 2019; Korsunsky et al., 2019; Pliner et al., 2019), even the latest methods are still benchmarked largely on broad cell type or subset integration, and often struggle (<50% classification accuracy) with fine cell states (Sikkema et al., 2022). Thus, we employed a random forest (RF) classifier-based approach, which has been applied successfully in work to identify correspondence in fine sub-clusters in the mammalian retina (Peng et al., 2019; Shekhar et al., 2016). Specifically, we employed paired RF models (one trained on FGID, the other trained on pediCD) to obtain cross dataset predictions per cell using an up-sampling procedure that enhanced accuracy ∼7-10% across each cell type (**Supplemental Figures 10-12**). With our final model, we attained cross-dataset predictions (pediCD to FGID & FGID to pediCD) for each cell, giving a probability score of a cell belonging to a subset in the other disease condition (**Supplemental Figure 5c; Methods**).

Some macrophage clusters characterized by *STAT1* activation did not demonstrate significant correspondence to any FGID cluster. We also generally noted substantially increased cluster diversity in pediCD end clusters relative to their correspondence in FGID. This emerged from more patient-specific clusters found in pediCD, and an overall decrease in Simpson’s index of diversity considering the patient composition of each end clusters (**Supplemental Figure 10b**).

Importantly, when we jointly clustered macrophages from FGID and pediCD together, we identified that several of the original end clusters identified through ARBOL in pediCD were divided across the UMAP: being split into distinct clusters of cells (**Supplemental Figure 10c-f**). This reinforces the need to quantitatively approach the choice of clustering parameters and number of iterations used. We also employed the STACAS package to integrate T cells between FGID, confirming the higher percentage of proliferating T cells in pediCD patients compared to FGID (**Supplemental Figure 11b**) (Andreatta and Carmona, 2021). Integration of T cells from the FGID and pediCD datasets (n = 29640 and 38031, respectively) was performed using the STACAS package (v1.1.0) (Andreatta and Carmona, 2021) Sankey plot was created using RAWGraphs 2.0 beta (https://github.com/rawgraphs) (Mauri et al., 2017). However, as compared to ARBOL, the integration approach on T cells resulted in lower heterogeneity, thus masking important disease-associated differences revealed by the ARBOL approach.

#### Pseudotime analysis of expression landscape on cells

The micrograin structure found through hierarchical tiered clustering is vital for being able to directly compare like cells across disease conditions, and find significant changes in phenotype and composition within individual subsets. It is also vital to understand how those like subsets relate to each other within a disease condition and how the larger macrograin structure differs across conditions. This macrograin structure can be explored through the gradients of gene expression among cells of a major type. Pseudotime and RNA-velocity are both excellent tools for exploring these gradients. For both tools, the choice of genes directly determines the structure found within the dimensional reduction, and thus what genes are chosen as significantly location specific within the resulting landscape of cells. for our purposes, as we knew we would be exploring a single cell lineage, and exploring the relationships of cell states within that space, we required for our dimensional reduction the genes common to that space. We selected genes by performing differential expression between the major cell type and all other cell types within that disease. We took the outer union of those genes. Then removed genes from the list found to be differentially expressed between disease conditions at the major cell type level. From these genes we performed PCA to 50 principal components and then computed a UMAP reduction to 2 components. This selection process allows the dimensional reduction to find smooth gradients between cells and provided a common space for cells of multiple disease conditions to exist.

From this common expression landscape we utilized Monocle3 https://cole-trapnell-lab.github.io/monocle3/ (Cao et al., 2019) to extract a best estimate linear path through the space, which we calculated a diffusion pseudotime on allowing use to numerically estimate the distribution of cells within the expression landscape. We utilized a list of genes that were cell-type defining genes in either FGID or pediCD, but removed genes that were differentially expressed between FGID and pediCD, to allow for cell type/subset to drive placement on the UMAP (Luo et al., 2022; Ordovas-Montanes et al., 2018). This allowed us to place our fine-grained clusters within a joint gene-expression space related to underlying cell types in FGID and pediCD. We note that caution must be taken in interpreting these findings as UMAP distances represent non-linear distances unlike PCA space. To compute the significance of changes in that distribution we used a permutation test of Hellinger distance between distributions. At each of 10,000 permutations we shuffled the group ordering within the comparison pair. We performed this test five times for comparisons between FGID vs FR, FGID vs PR, NOA vs FR, NOA vs PR and FR vs PR. Our threshold was set as Bonferroni corrected p_value < 0.05.

#### Bulk RNA-seq library generation from PREDICT patients

Population RNA-seq was performed as follows. In brief, tissue was collected directly into RLT Plus lysis buffer (Qiagen) and stored at −80°C until RNA isolation. RNA was isolated from 30 patients using a Qiagen Qiashredder protocol followed by total RNA isolation with the RNeasy Plus Mini Kit (Qiagen). Total RNA was quality controlled via Agilent high-sensitivty RNA Tapestation assay (Agilent). Total cDNA synthesis was performed using Takara SMART-Seq v4 Ultra Low input RNA kit with 10ng of RNA input from samples with RIN values >8 (Takara). cDNA was amplified and cleaned using a 0.6x SPRIselect protocol (Beckman Coulter). Sequencing libraries were generated following the Illumina NXTR XT DNA SMP Prep Kit and Index Kit (Illumina). Libraries were pooled post-Nextera and cleaned using Agencourt AMPure SPRI beads with successive 0.7X and 0.8X ratio SPRIs. Sequencing was performed on Illumina HiSeq®2500 (Illumina) by multiplexed single-end read run with 80 cycles (split from HiSeq SBS Kit V4 250 cycle kit, FC-401-4003). Samples were sequenced at an average read depth of 15 million reads per sample.

#### Tissue bulk RNA-seq Data Analysis

Tissue and sorted basal cell samples were aligned to the GRCH38-2020-A genome and transcriptome using STAR and RSEM. One sample (p022) was excluded as an outlier based on PCA space so we retained 26 samples for further analyses. Differential expression analysis was conducted using DESeq2 package for R. Genes regarded as significantly differentially expressed were determined based on an adjusted p-value using the Benjamini-Hochberg procedure to correct for multiple comparisons with a false discovery rate <0.05.

#### 92-gene signature derived from pediCD-TIME

In order to generate a gene signature from pediCD scRNA-seq data to apply to bulk RNA-seq data, we took the 25 end clusters associated with FR and PR to anti-TNF in the pediCD-TIME-negative (PC2 negative) direction (Figure 5), and for each of those clusters we selected the top 10 best-scoring markers using the rank-score function (described above).

Duplicate hits were only accounted once, and genes that appear as hits in both the positive and negative directions were filtered out. This gene signature is in **Supplemental Table 17**.

#### Gene set enrichment analysis

Fold changes between patients responding or not responding to anti-TNF therapy from RISK and E-MTAB-7604 cohorts were calculated with Seurat (v4.0.3) (Haberman et al., 2014; Hao et al., 2021) and DESeq2 (v1.30.1) (Love et al., 2014) packages, respectively. GSEA analysis was performed using the fgsea R package (v1.16.0) (Korotkevich et al., 2021). Genes with similar fold changes were preranked in a random order. The code for this analysis can be found in the GitHub repository https://jo-m-lab.github.io/3p-PREDICT-Paper/4_GSEA/PREDICT_GSEA_final.html.

#### Differential expression testing

To calculate differential expression between FR and PR groups, for each subset with a least 50 cells in each condition we used a Wilcoxon test thresholded to 0.05 Bonferroni corrected p-value and down sampled using the “max.cells.per.ident” argument within Seurat’s ‘FindMarkers’ function to a maximum of 10000 cells. The limits on minimum and maximum number of cells were chosen to mitigate issues with comparisons between disproportionate populations and computational efficiency. There does still exist 2 orders of magnitude between the minimum and maximum; however, the subsets most of interest and reported through the manuscript are within the same order of magnitude There are noted spillover effects within the expression tests. We observe ubiquitous contamination of genes such as *IGHA1*, *IGHG1* and *DEFA5* across all cell types and subsets. These genes are routinely found as enriched within more severe inflammation, beyond even this dataset. This is a real effect, but less than useful for understanding driving factors within individual cell subsets. So, we focus on significant differentially expressed genes that also have a high pct.cells.expressing.in/ pct.cells.expressing.out ratio.

#### Understanding how additional outgroups influences cluster diversity and stability through entropy testing

By nature of clustering over two distinct diseases in **Figure 6**, the specific Simpsons’ Diversity for each cluster is less than what was observed for pediCD and FGID alone, but still highlights that despite these clusters representing ∼0.05-0.1% of cells, they are remarkably composed of cells sampled from most patients within a disease category (**Supplemental Figure 15**). In particular, we decided to measure the degree to which end clusters maintain their composition, or if cells are split or shuffled into different clusters across trees. To answer this question, we calculate the log base 2 Shannon entropy per curated end cluster to compare the joint ARBOL to the original separate ARBOL’s to determine how many groups each end cluster is split into in an alternate tree. A lower entropy corresponds a majority of the same cells composing a similar cluster in the alternate tree, providing a quantification of the preservation of label-level information, per label, between two trees. These entropies are compared to simulations of randomly scrambled end clusters of the same sizes mapping to the same number of end clusters in the new tree, with a fixed percentage mapping to a single new cluster, while the other cells map randomly.

To calculate representative label preservation, Shannon entropy was calculated as 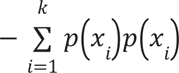 where *x*_1_…*x*_k_ are the set of end cluster labels in a new ARBOL, and *p*(*x*) is the fraction of cells of that curated cluster in each of the *k* new labels. Simulations of 75% label preservation, 50% label preservation, and random shuffling were performed 100 times per pediCD atlas T/NK/ILC cluster size with 57 possible labels. In simulations with preservation, unpreserved labels are shuffled randomly among the other 56 new labels. The average of the 100 simulations per cluster size are displayed in **Supplemental Figure 15**. Entropies are calculated per pediCD atlas cluster to determine preservation of CD severity vector cell states in new atlases, which contain a higher number of labels. We fixed cluster size and n possible labels to pediCD atlas sizes in simulations, as maximum Shannon entropy scales with both cluster size and number of new labels, such that calculation of joint atlas labels to pediCD alone finds a lower distribution of entropies than the reverse, as there are fewer possible labels in the curated pediCD atlas than the raw joint FGID-pediCD ARBOL.

Simulation of 75% label retention finds entropies between 1 for an end cluster of 8 cells, and 2.1 for an end cluster of >2000 cells. Similarly, simulation of 50% label retention finds entropies of 1.9 to 3.9. Random shuffling of cell labels finds an average of 5.7. We first mapped the pediCD atlas end clusters to the joint atlas and identified that half of end clusters fall under an entropy of 2 with most clusters below the maximum 50% label retention simulation of 3.8 (**Supplemental Figure 15**). We then took the joint atlas end clusters and mapped them to the pediCD atlas end clusters and recovered a similar distribution. Importantly, these values are all significantly less than when labels are randomly shuffled, whereby entropy values rapidly approach 5.8 with increasing label size. We find that our high-resolution pediCD clusters vary between near perfect label preservation, as many did not mix with FGID cells (**Supplemental Figure 15 right** NK.MKI67.CD38), to a state where they are combined into one larger cluster (**Supplemental Figure 15 right** T.STMN1.RRM2), and below 50% label retention, as some larger clusters were broken up and mixed with FGID (**Supplemental Figure 15 left** T.CCL4.GZMK). It is of interest to note how an “outgroup” condition may influence the resulting entropy of clusters. In these cases, recombination of end clusters based on sample diversity may reduce entropy between ARBOL results.

#### Joint FGID-CD ARBOL cluster frequencies analysis

We performed analysis of the compositional PCA space of cell type abundances of T, Myeloid, and Epithelial subsets in the joint ARBOL to determine key similarities between the NOA anti-TNF response category and FGID patients. Celltypes were named using our automated naming method. The top 20 cell types in the positive direction of PC1 were selected and visualized using a heatmap of z-score normalized per-patient frequencies. Samples were manually ordered according to disease and antiTNF_response (NM [Not Measured / FGID], NOA, FR, PR) to facilitate direct comparisons across groups. This method enabled clear visual inspection of compositional similarities and differences among patient categories based on hierarchical clustering, revealing subsets which overlap between NOA and FGID patients.

The automated joint-ARBOL names were then compared to the original curated celltype names in both the FGID and CD atlases separately with alluvial plots to give context to the overlapping clusters.

#### Pediatric-to-adult disease trajectory analysis in integrated atlas cell-state composition space

We performed Harmony integration on adult CD, pediCD, and pediCD longitudinal samples using these 3 categories as the harmony variable. Harmony and SCTransform normalization was performed at each iteration of subclustering to create the resulting integrated ARBOL atlas. Compositions of T, B, Myeloid, Fibroblast, and Plasma cells were calculated per sample by normalizing ARBOL cluster abundance per sample by total number of these celltypes combined in that sample. PCA was performed on the resulting sample x composition matrix, resulting in **Figure 8d** (left). We plugged this matrix into a Seurat object and used Seurat functions to perform nearest neighbors embedding and UMAP, resulting in **Figure 8d** (right). This Seurat object was then converted to a Monocle CellDataSet object with a negBinomial.size() expression family. A pseudotime graph was then learned in UMAP space with geodesic_distance_ratio 0.5 and minimal_branch_len 5, with no partitions. The graph was rooted in the NOA cells to begin the pseudotime trajectory at the earliest timepoint in disease trajectory.

To discover cell states varying along the pseudotime trajectory, we used Monocle 3’s graph_test() algorithm, an implementation of the Moran’s I test of spatial autocorrelation. Fig 8f counts the top 50 autocorrelated cell states along pseudotime per celltype ranked by q-value. Cell states’ changes over pseudotime were then visualized using ggplot geom_smooth with a linear cubic model to display cell state presence patterns over pseudotime. This model is estimated with geom_smooth(method = “lm”, formula = y ∼ splines::bs(x)).

To develop a deeper understanding into which cell types varied concordantly and which were more sample or condition-dependent, we used the TradeSeq (https://github.com/statOmics/tradeSeq) package (**Supplemental Figure 17**). TradeSeq provides an association test which we used in a leave-one-out cross validation experiment, testing the association of each cell state with pseudotime across bootstraps. We then used TradeSeq’s trajectory clustering method to reveal trends in trajectories, named by trend (**Supplemental Figure 17b**), and overlaid the bootstrap results onto them. Bootstrap tests showed that T, Myeloid, and Epithelial (EC) subsets are robustly associated with the disease pseudotime, and that they generally follow downward-hill, downward-slope, quick-climb, and upward-hill trends.

#### Multiplex Immunohistochemistry

A fully automated Multiplex Immunohistochemistry assay was performed on the Ventana Discovery ULTRA platform (Ventana Medical Systems, Tucson, AZ), and as previously described(Marron et al., 2022). The assays were optimized for HCC. Optimal concentrations of each antibody were determined, and they were applied in the following sequence and detected with the indicated fluorophore.

Pan-Immune Panel:

1. Rabbit Anti-CD20 (Clone SP263, Abcam, ab64088) was detected with DISCOVERY Rhodamine 6G (Roche, Part number: 7988168001).
2. Rabbit anti-CD3 (Clone SP162, Abcam, ab135372) was detected with DISCOVERY DCC (Roche, Part number: 7988192001).
3. Rabbit anti-CD8 (Clone SP239, Abcam, ab178089) was detected with DISCOVERY RED 610 (Roche, Part number: 7988176001).
4. Rabbit Anti-CD68 (Clone SP251, Abcam, ab192847) was detected with DISCOVERY Cy5 (Roche, Part number: 7551215001).
5. Rabbit Anti-FOXP3 (Clone SP97, Abcam, ab99963) was detected with DISCOVERY FAM (Roche, Part number: 7988150001).

Following staining, the tissue was counter-stained and cover slipped with Invitrogen ProLong Gold Antifade Mountant with NucBlue. Whole slide imaging was performed on the Zeiss Axioscan which was equipped with a Colibri light source and appropriate filters for visualizing these specific fluorophores. Where applicable, an additional marker, Mouse anti-Pan Keratin (Clone AE1/AE3/PCK26, Roche, Part number: 5266840001) was added. Coverslips were removed by immersing the slides overnight in distilled water, staining with the PanCK antibody and detecting with Goat anti-mouse IgG (H+L)-Cy7 (AAT Bioquest, 16856). Slides were re-cover slipped and scanned as described above.

#### Quantitative image analysis

Quantitative image analysis was preformed using HALO Indica Labs Hyperplex module (IndicaLabs, Albuquerque, NM). For each sample, images from the pan-immune panel and the pan-CK were fused to generate one single image. A first classifier was applied to detect the tissue on each section and an automatic annotation was generated as “whole section”. A second classifier was applied using panCK and DAPI channels to define the epithelium layer (panCK positive) and the lamina propria (panCK negative). Automated annotations were generated as “epithelium” and as “lamina propria”. Immune cells were detected in each area of interest (whole section, epithelium, lamina propria). T cells were defined as CD3+, B cells as CD20+, myeloid cells as CD68+. For each T cell subset, CD8 T cells were defined as CD3+CD8+FOXP3-, CD4 as CD3+CD8-, Tregs as CD3+CD8-FOXP3+ and CD4 conventional as CD3+CD8-FOXP3-. Numbers of positive cells for each immune subset were counted and their density measured.

#### General Statistical Testing

Parameters such as sample size, number of replicates, number of independent experiments, measures of center, dispersion, and precision (mean +/- SEM) and statistical significances are reported in Figures and Figure Legends. A p-value less than 0.05 was considered significant. Where appropriate, a Bonferroni or FDR correction was used to account for multiple tests, as noted in the figure legends or Methods. All statistical tests corresponding to differential gene expression are described above and completed using R language for Statistical Computing.

## Supplemental Figure Titles and Legends

**Supplemental Figure 1.**
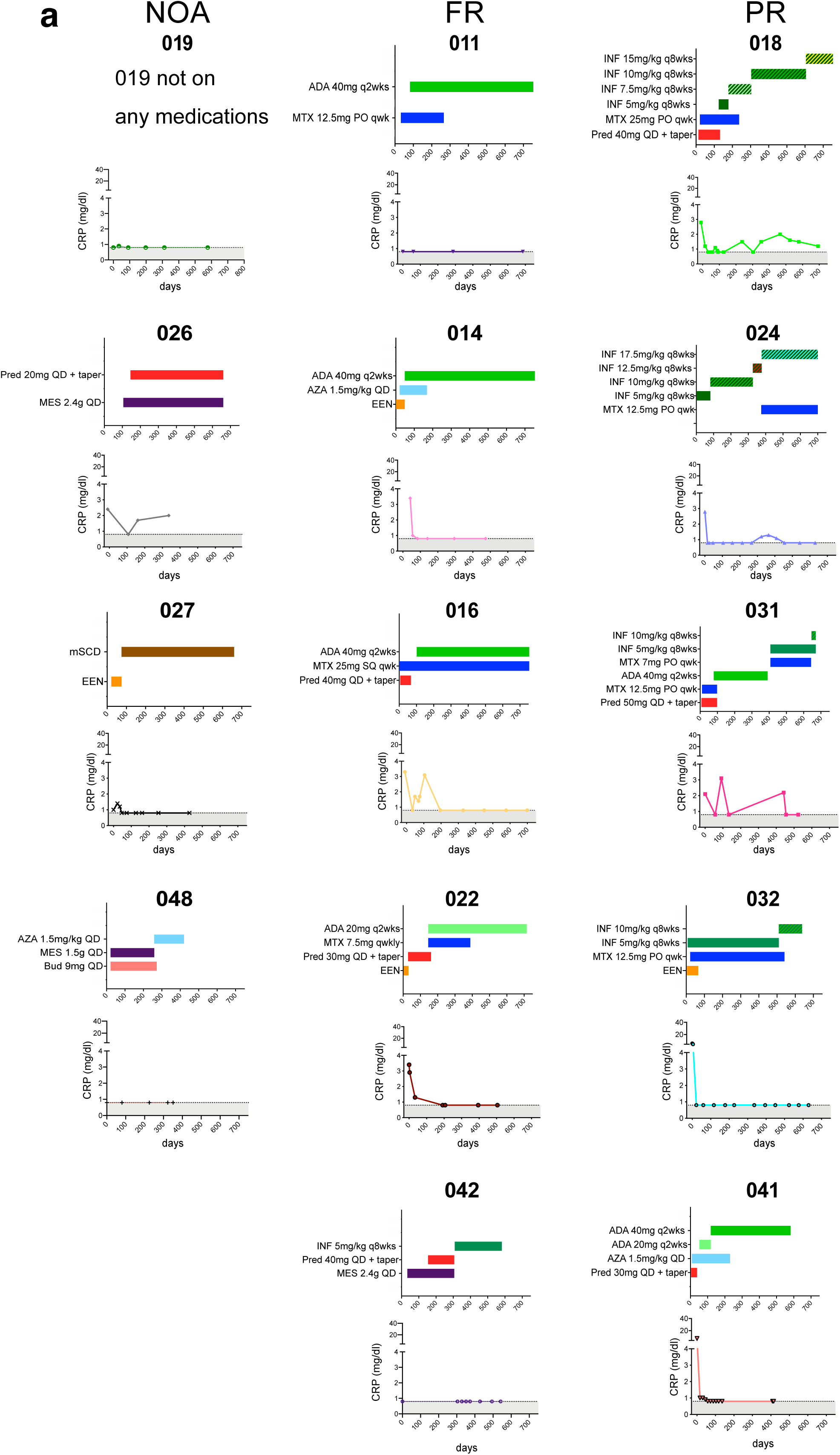
Clinical trajectory and treatments for all pediCD patients. a. Representative treatment history and clinical inflammatory parameters used for determination of NOA, FR and PR status for all pediCD patients (see **Methods**, **Table 1**, and **Figure 1**; ADA: adalimumab, INF: infliximab; MES: mesalamine MTX: methotrexate; Pred: prednisone; mSCD: modified specific carbohydrate diet; EEN: exclusive enteral nutrition).

**Supplemental Figure 2.**
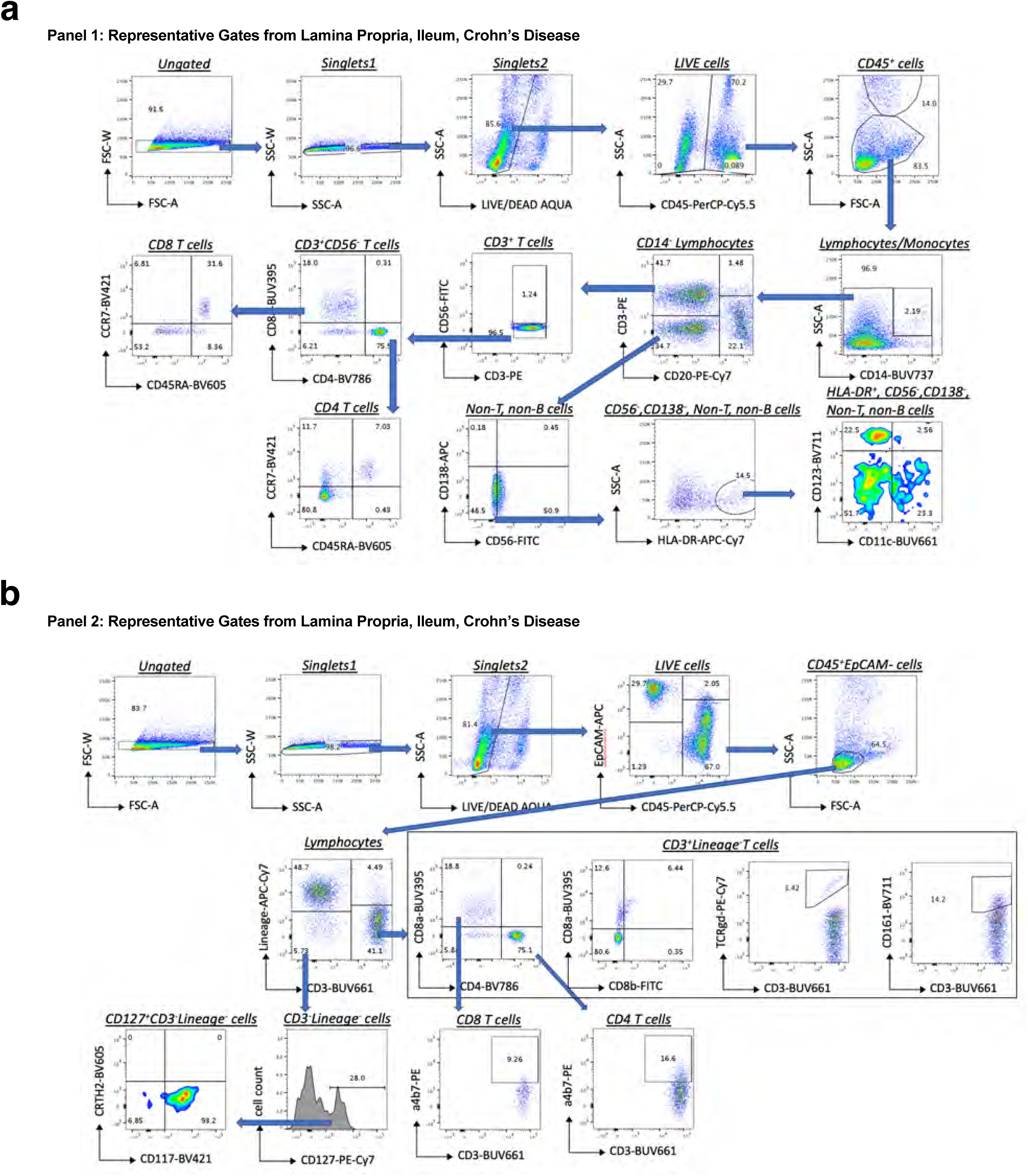
Representative gating strategies for flow cytometry. a. Representative gating strategy for Panel 1 focused on T cells and myeloid cells, for antibodies please see **Supplemental Table 1**. b. Representative gating strategy for Panel 2 focused on non-classical T cells and innate lymphoid cells (NB: Lineage = CD14, CD20, CD11c, CD11b, CD56), for antibodies please see **Supplemental Table 1**.

**Supplemental Figure 3.**
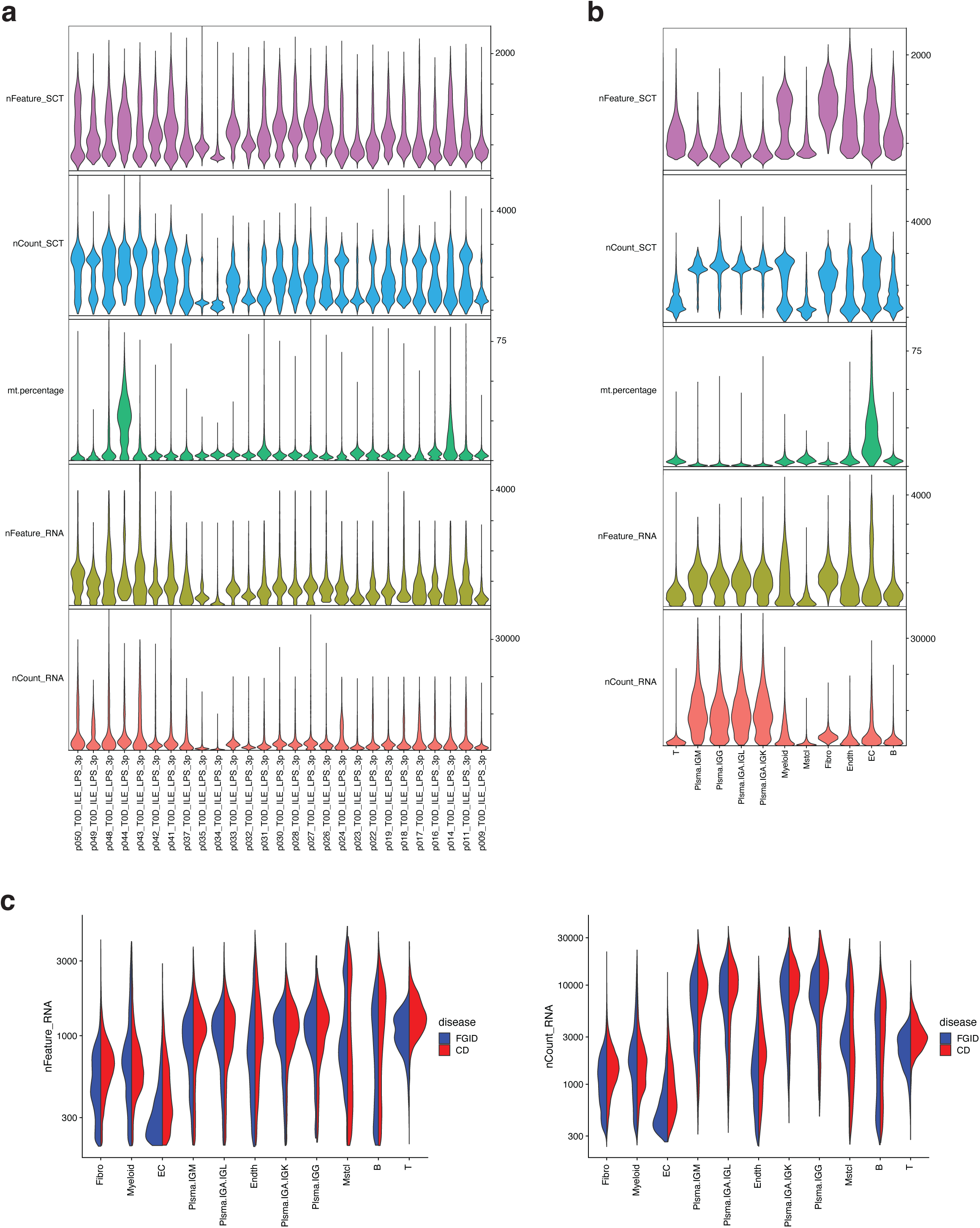
Comparison of quality control measures reveals similar sequencing depths and gene capture between FGID and pediCD. a. Quality control measures for scRNA-seq of ileal biopsies of 27 patients (13 FGID, 14 pediCD) included in the study. Top two graphs denote total genes (nFeature) and UMIs (nCount) after normalization with SCTransform. Lower graphs denote total genes (nFeature), UMIs (nCount) and mitochondrial read percentage (mt.percentage) of pre-processed 10X 3’ v2 single-cell RNA-sequenced samples. Cutoffs for individual samples are represented in **Supplemental Table 2**. b. Quality control measures as in **a** split by cell type. c. Comparison of total genes captured (nFeature, left) and total UMIs (nCount, right) between FGID (blue) and pediCD (red) split by cell type.

**Supplemental Figure 4.**
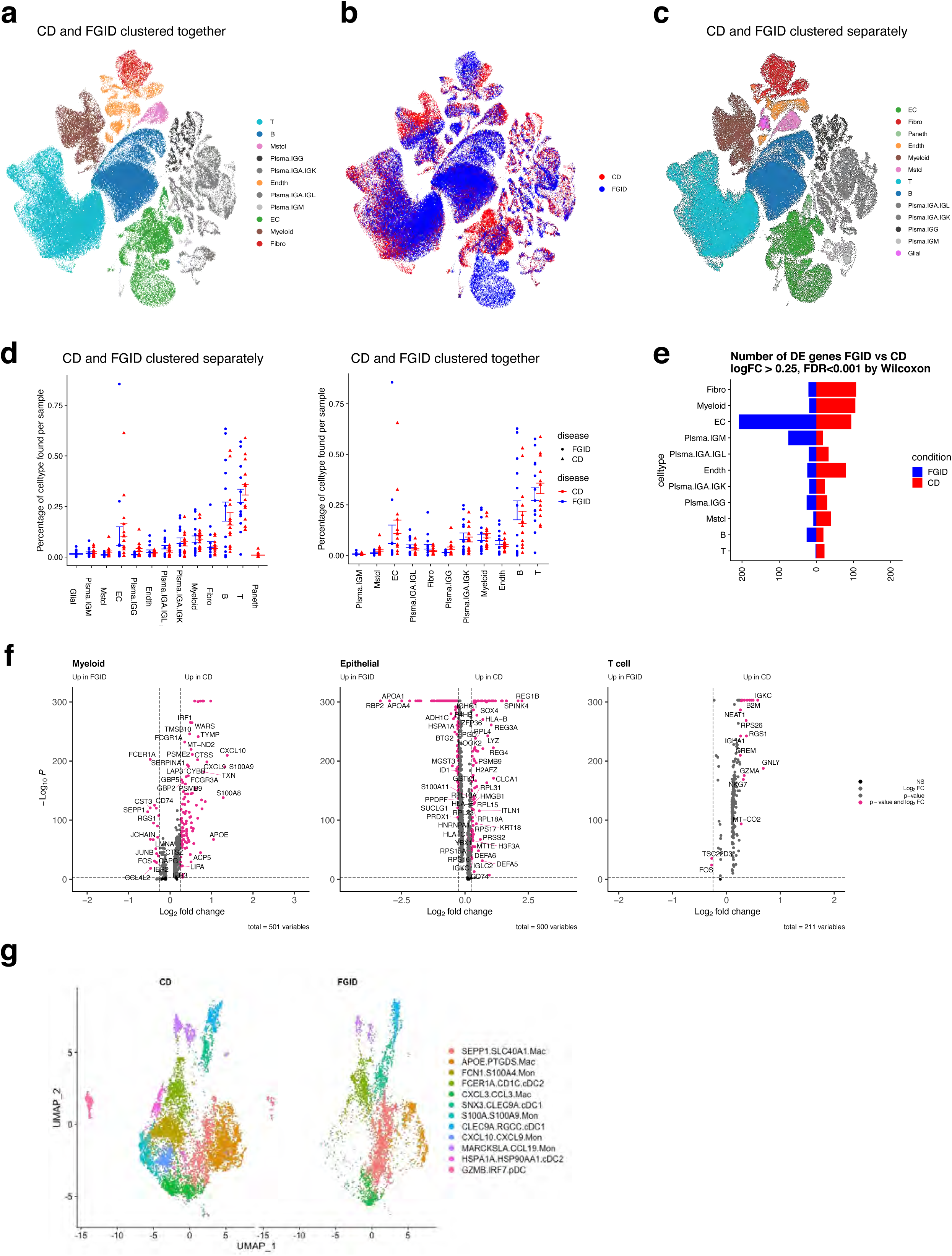
Traditional clustering with SCTransform normalization reveals similarities across cell types in FGID and pediCD. a. UMAP representing one round of clustering of 197,281 single-cells across FGID and pediCD samples. Traditional clustering performed on both diseases together. Colors represent major cell types determined by one round of clustering with Seurat RunUMAP parameters (PCs = 1:50, n.neighbors = 50, min.dist = 1). Cell types were assigned based on significantly upregulated marker genes (Wilcoxon; p.adj<0.05) obtained from comparison of specific cell type versus all other cell types. b. UMAP as in **a** colored to highlight FGID (blue) and pediCD (red) cells. c. UMAP as in **a** colored by Tier 1 ITC clusters performed separately for FGID and pediCD (NB: this was used for **Figure 3** and **Figure 4**). d. Comparison of cell cluster frequencies between FGID (blue) and pediCD (red) with cells clustered (left) separately or (right) together. Patient contributions denoted by circles (FGID) and triangles (pediCD). e. Differentially expressed genes across cell type in FGID vs pediCD determined to be significant by Wilcoxon test (logFC>0.25, FDR<0.001). f. Volcano plots for Myeloid, Epithelial, T-cell clusters denoting differentially expressed genes in FGID vs. pediCD. Those in pink are significant by Wilcoxon test; **Supplemental Table 4** for full gene lists. g. UMAPs of jointly clustered pediCD and FGID Myeloid cells.

**Supplemental Figure 5.**
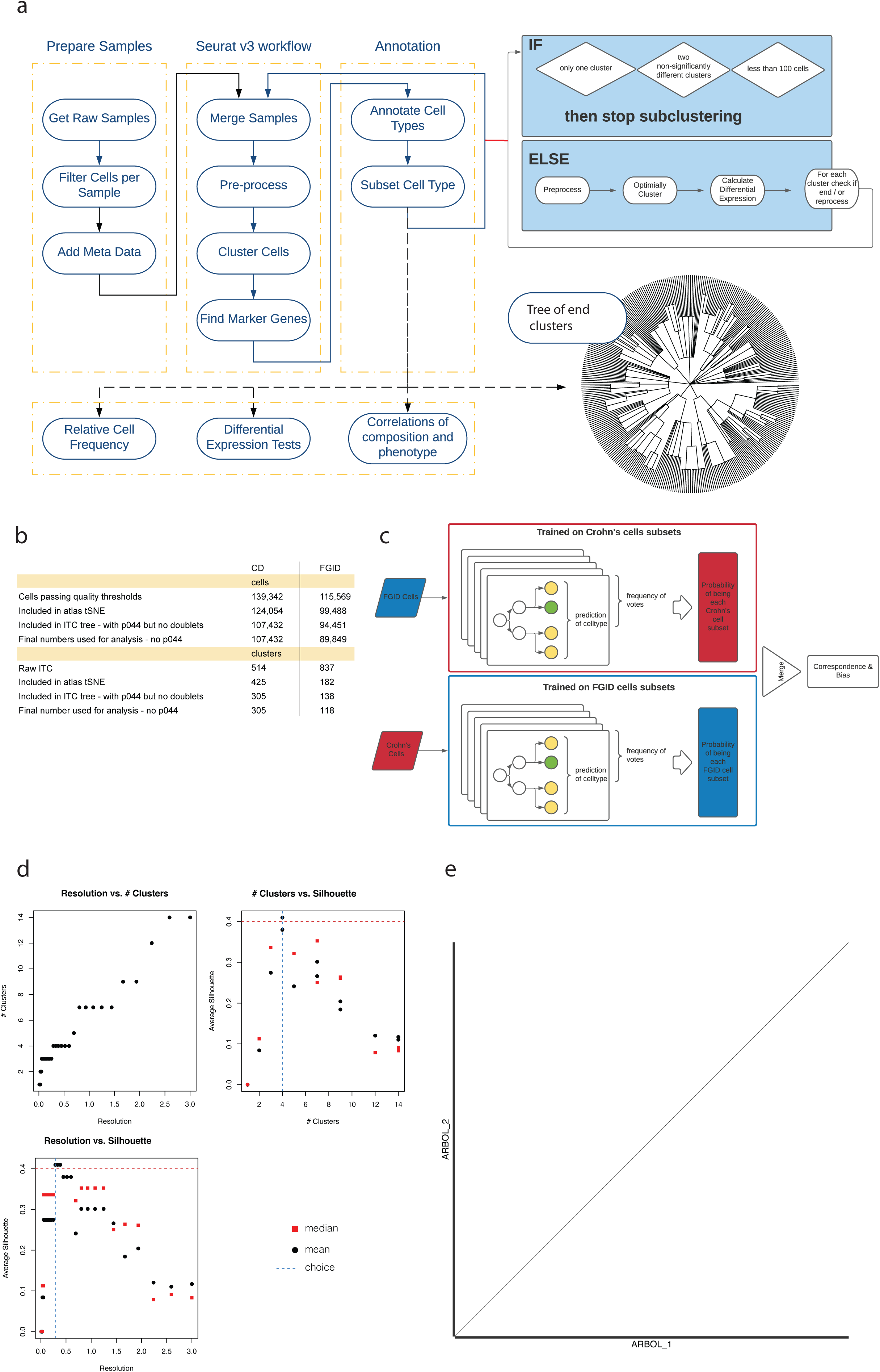
Schematic for iterative tiered clustering and random forest classifier approach. a. Flowchart depicting iterative tiered clustering (ITC) used for generating FGID and pediCD cellular atlases. After sequencing, cells underwent quality control and a cell by gene expression matrix was derived from the 27 ileal samples. Dimensionality reduction and graph-based clustering were performed using the standard Seurat workflow to annotate cell types. Resulting clusters were then iteratively processed through the same pipeline unless end conditions were met. Each cluster was checked for three end conditions which included: only one cluster remaining, two clusters remaining with no more than 5 up and down regulated genes as determined by Wilcoxon test (logFC > 1.5, FDR < 0.001), and/or less than 100 cells in the cluster. Iterative clustering stopped if any of the three conditions are met. Unlike traditional clustering, in ITC principal component and clustering resolution parameters are chosen automatically. Stop conditions are built in as parameters to the ITC pipeline, allowing customization to the dataset. From the results of ITC, we then build the cell cluster taxonomical tree through hierarchical clustering (**Figures 3 and 4**). b. Cell and cluster numbers after various processing steps tabulated c. Random forest classifier approach for integrating FGID and pediCD datasets. FGID and pediCD datasets were used as training datasets to create random forest predictors used in downstream sub-clustering of cell types and subsets. The opposing dataset was then tested by each algorithm independently to determine correspondence and bias as depicted in **Figure 6 and Supplemental Figure 8**. d. A visual representation of the parameter scanning method outputs used that illustrate resolution vs. clusters, numbers of clusters vs. silhouette, and resolution vs. silhouette. The resolution parameter is optimized to maximize the silhouette score. e. ARBOL was run independently in two distinct Google Cloud Platform Terra sessions by two distinct computationalists on the nasal polyp dataset from Ordovas-Montanes et al., returning a 1:1 correlation of clustering results (Ordovas-Montanes et al., 2018, p.).

**Supplemental Figure 6.**
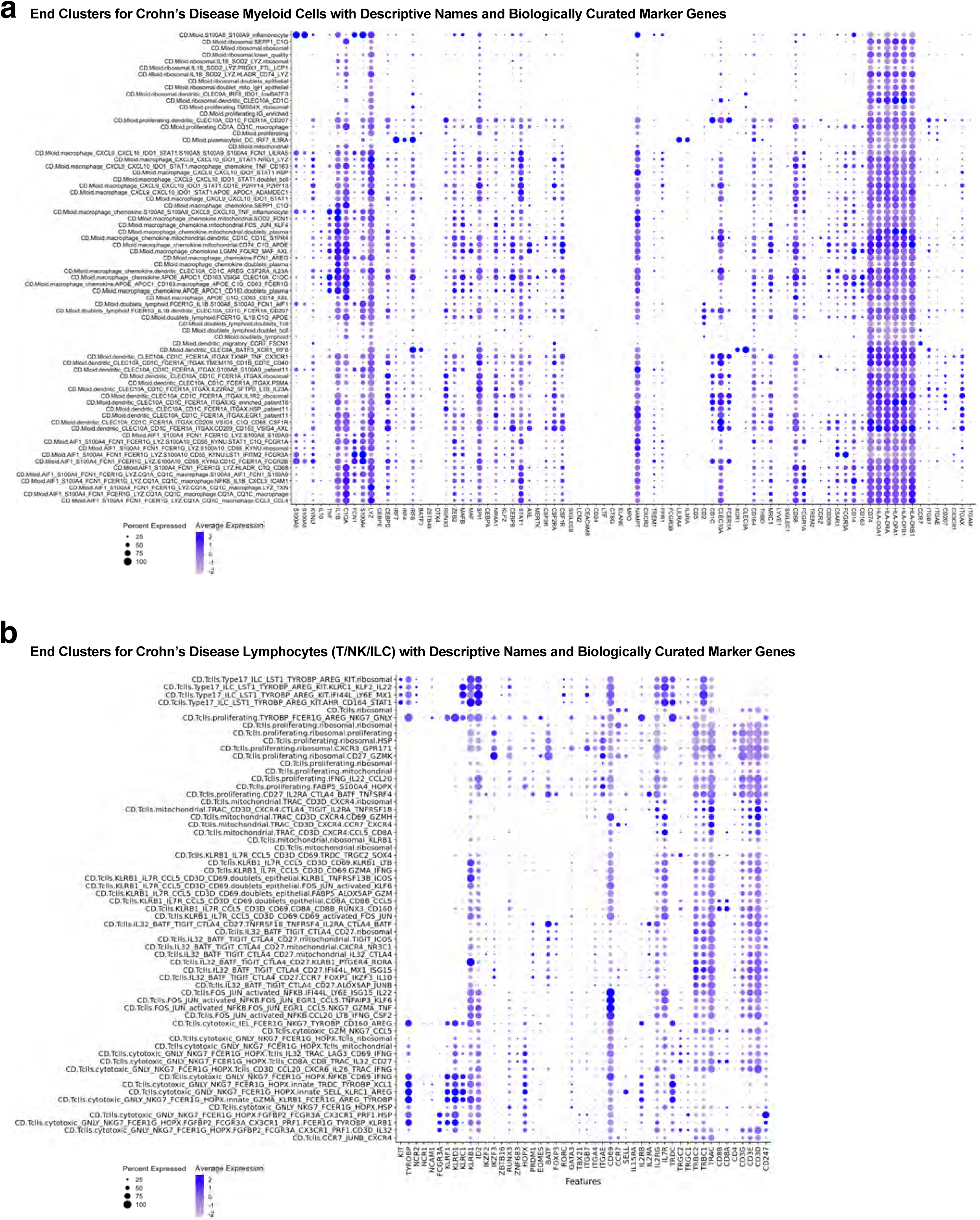
Representative marker genes for myeloid and T cells. a. Dot plot of curated genes related to myeloid biology. Dot size represents fraction of cells expressing the gene, and color intensity represents binned count-based expression level (log(scaled UMI+1)) amongst expressing cells. All cluster defining genes are provided in **Supplemental Table 7 and Supplemental Table 10**. Dot size is only plotted if more than 5% of cells are expressing the transcript. Names are descriptive names generated from inspection of ITC output which were then converted to standardized naming scheme as in **Methods**. b. Dot plot of marker genes related to T/NK/ILC lymphoid biology as in **a**.

**Supplemental Figure 7.**
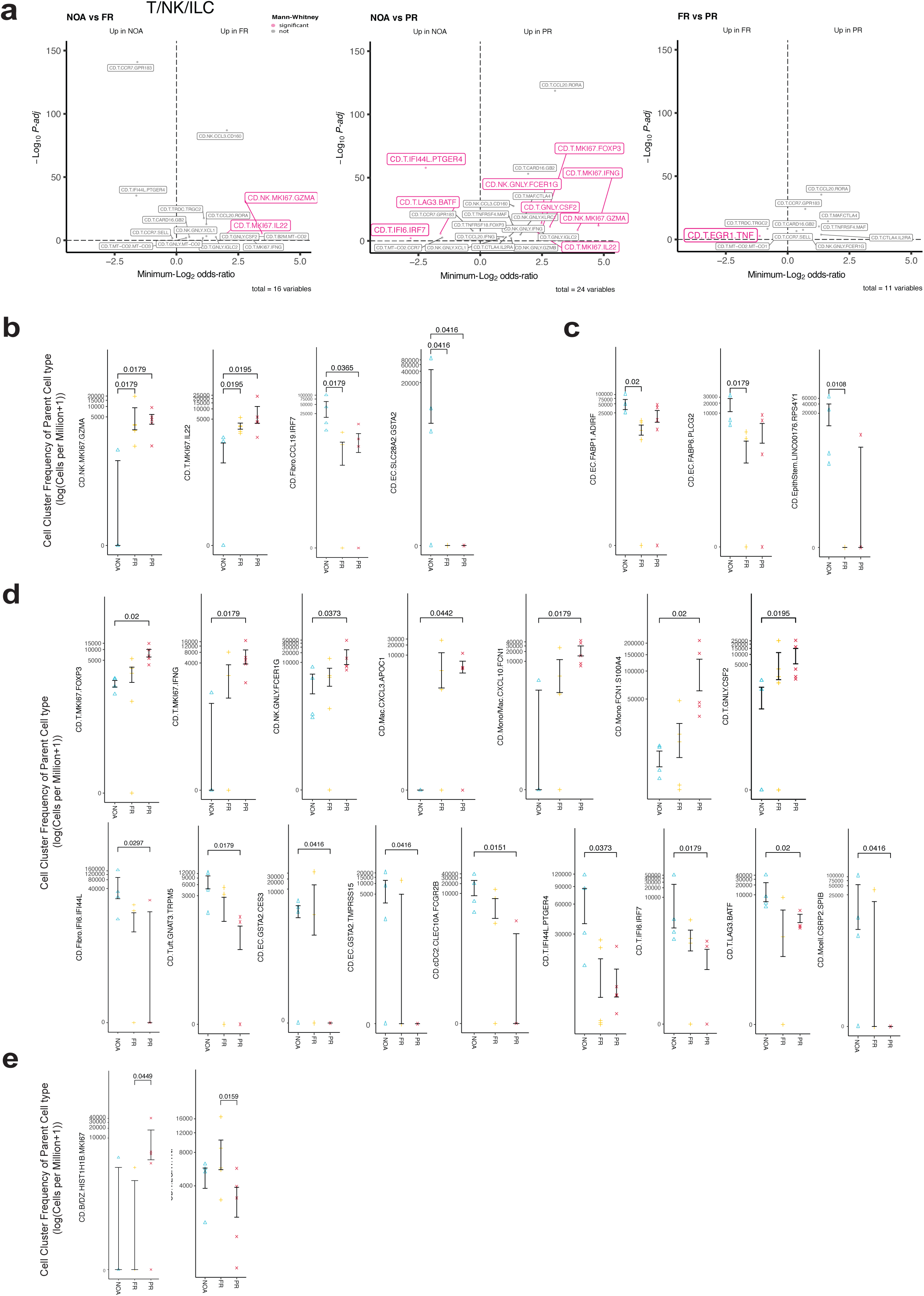
Cell types associated with pediCD severity after PCA analysis. a. Volcano plots for T/NK/ILC clusters between NOA, FR and PR, where named clusters are significant by Fisher’s exact test and those in pink are significant by Mann-Whitney U test. b. Cell cluster frequencies of the parent cell type found to be significant by Mann-Whitney U test between NOA and FR/PR. c. Cell cluster frequencies of the parent cell type between NOA and FR (as above). d. Cell cluster frequencies of the parent cell type between NOA and PR (as above). e. Cell cluster frequencies of the parent cell type between FR and PR (as above).

**Supplemental Figure 8.**
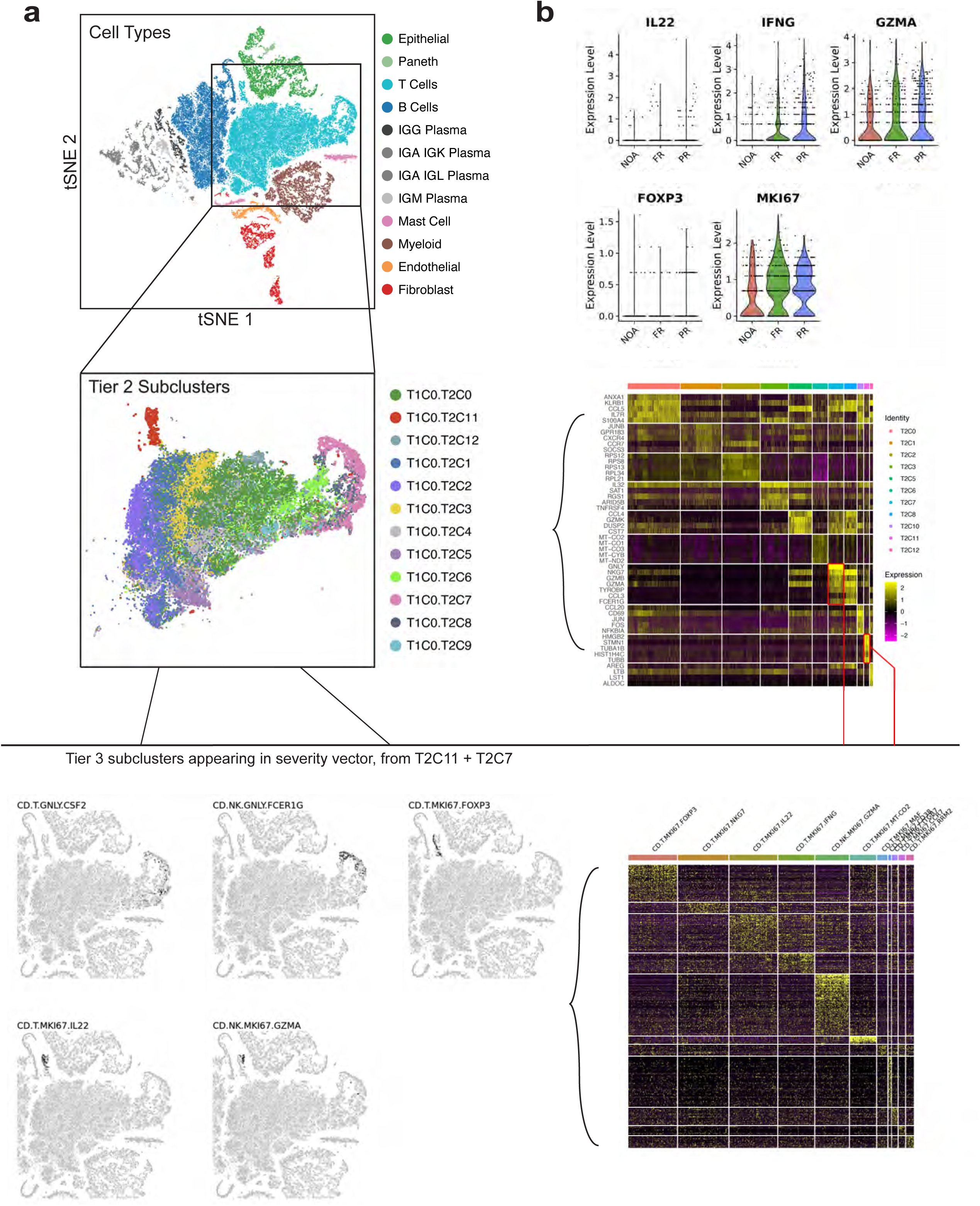
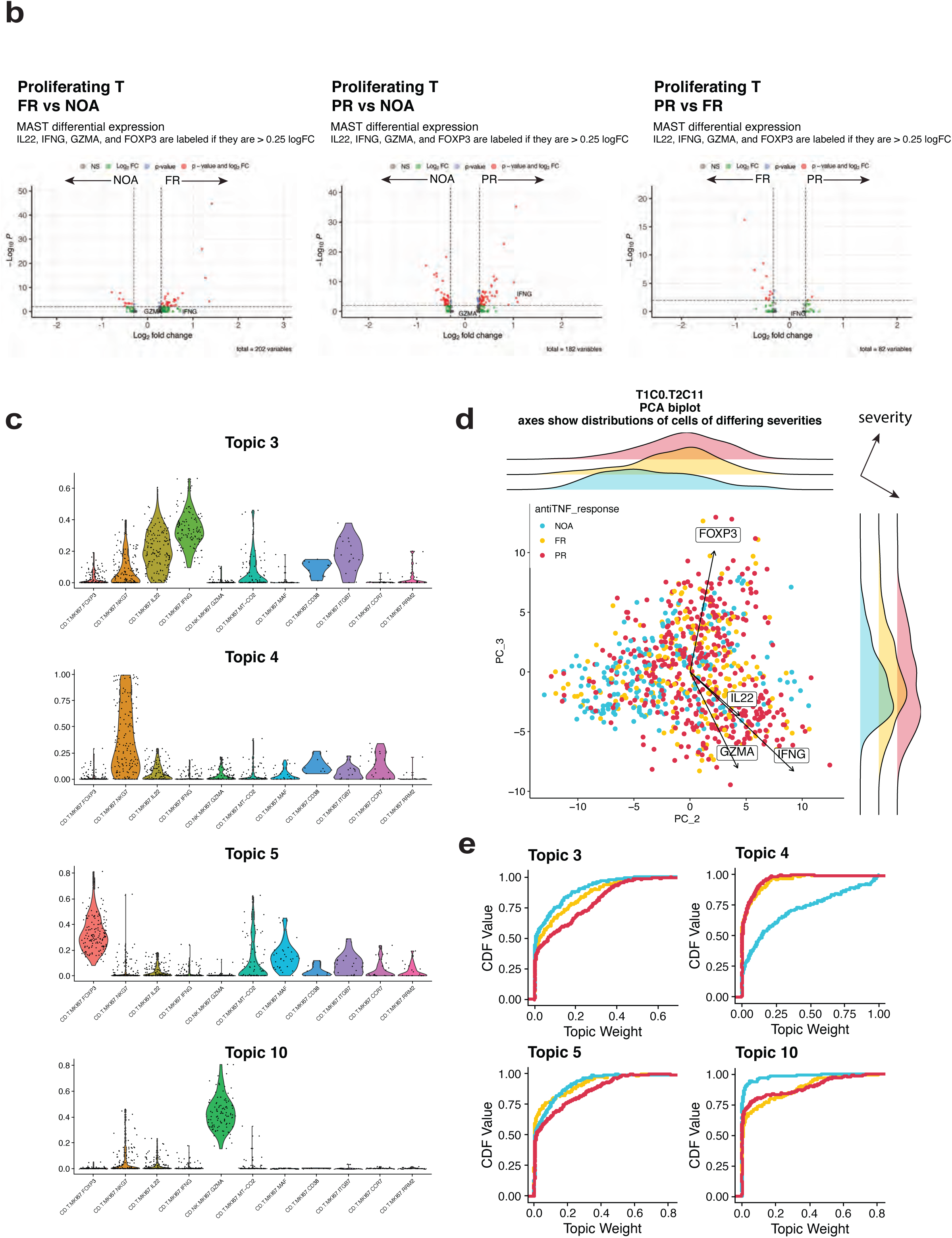
Illustrative example of ARBOL on proliferating T cells and comparison with differential expression and topic modeling. a. t-SNE representation of ARBOL clusters at the overarching cell type (top), tier 2 clustering of T/NK/ILC (middle), and end-cluster levels (bottom). ARBOL sub-clusters at each level based on gene modules with appreciable biological relevance, as shown by heatmaps of genes (rows) expressed in sub-clusters at both the tier 2 and end-cluster levels. Red box in tier 2 heatmap (middle) denotes ARBOL sub-clustering identification of proliferating T and NK clusters, of which several sub-clusters (bottom) are important for severity. Severity associated T/NK/ILC end-clusters (bottom) are found in multiple tier 2 clusters (T1C0.T2C11 + T1C0.T2C7). b. Violin plots of select genes important to ARBOL end-clusters and volcano plots of all genes at the level of proliferating T cells (T1C0.T2C11). MAST differential expression at this level between severity conditions finds *IFNG* significantly upregulated in PR vs. NOA, but not significant in other tests, nor are *FOXP3, GZMA,* or *IL22* in any comparison. c. Topic modeling performed on proliferating T cells (T1C0.T2C11) finds topics unique to ARBOL end-clusters, providing another avenue for identification of gene modules found by high-resolution ARBOL sub-clustering. d. Topics found in **(c)** and principal components used by ARBOL for sub-clustering T1C0.T2C11 display covariation with severity conditions. e. Topics found in (c) and their relationship to specific anti-TNF disease outcomes.

**Supplemental Figure 9.**
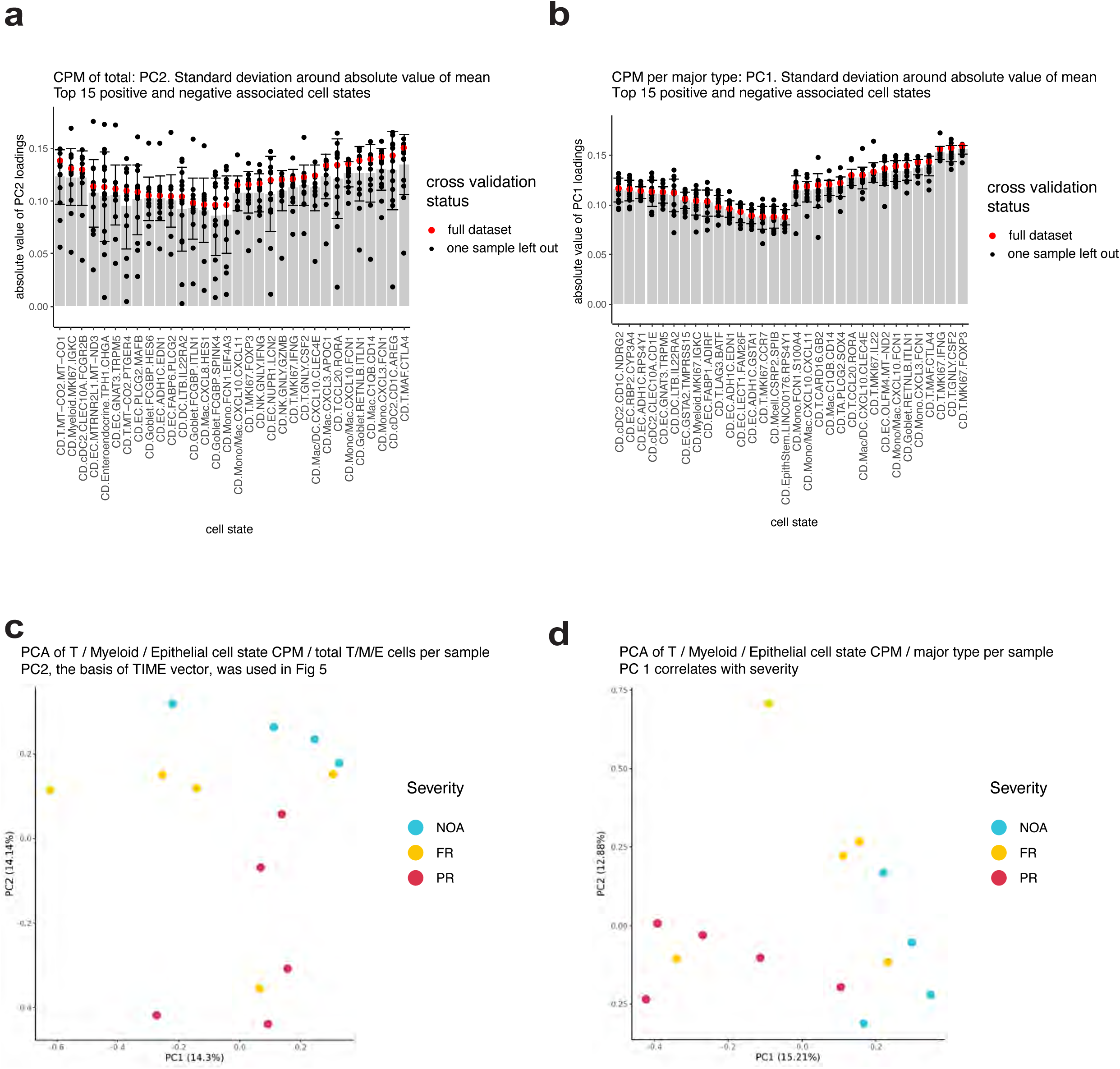
Leave-one-out cross-validation (LOOCV) to determine robustness of cluster contributions to PC loadings in pediCD severity vector. a. Top 15 positive and negative pediCD-TIME associated ARBOL cell clusters. Loadings of PC2 (pediCD-TIME) from PCA of CPM per total cell number per patient calculated with full dataset in red and grey bar showing mean of LOOCV. Error bars show standard deviation around the mean. b. As in **a,** but CPM calculated per cell type (e.g. T.MKI67.FOXP3 /(n T/NK/ILC) * 100000 per patient). PC1 of per-cell-type CPM matrix is associated with severity (R=0.72) and is more robust to LOOCV. c. Full dataset PCA used in **a (as in Figure 5).** d. Full dataset PCA used in **b.**

**Supplemental Figure 10.**
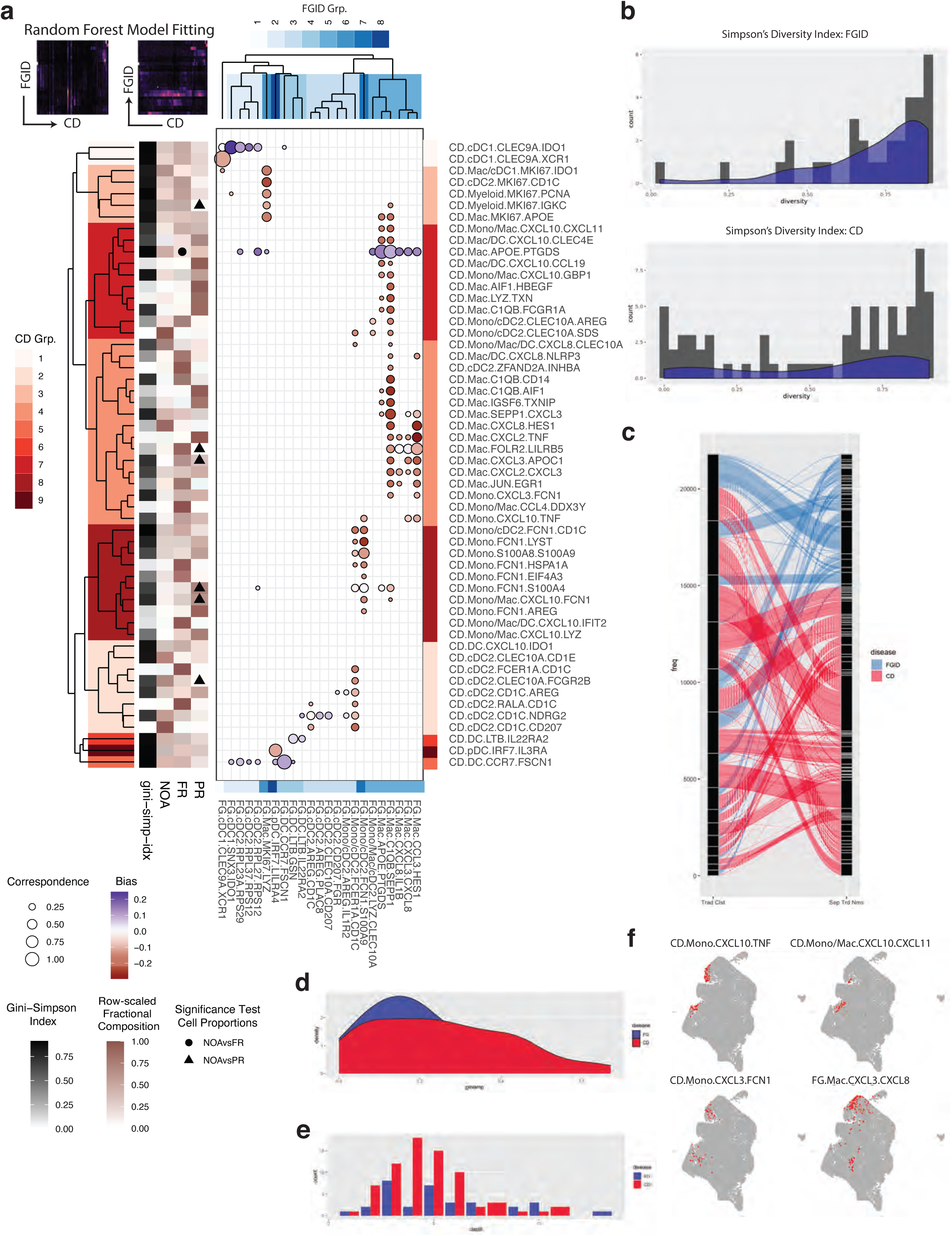
Random Forest (RF) Classifier Applied to Myeloid Cellular Taxonomies Identifies Correspondence between FGID and pediCD. a. Correspondence between cell subsets from FGID-to-pediCD and pediCD-to-FGID. <u>Top left heatmaps</u>: RF probabilities for each cell averaged over subset to gain probability of each FGID matching onto each pediCD subset (left), and pediCD onto FGID (right). <u>Bubble plot (center)</u>: size = sum(probability matrices) for confidence of predictions, marker color = diff(probability matrices) to show which direction the RF model is more confident on, e.g. more likely for FGID subset to belong to pediCD subset or pediCD subset to belong to FGID subset. Markers are filtered to show the top 10^th^ percentile of correspondence. <u>Dendrograms</u>: separated-tiered clustering on prediction probabilities of FGID (blue) and pediCD (red) using complete linkage with correlation distance metric, clusters are cut at height 0.7 (range 0-1). <u>Heatmap</u>: 1-Gini-Simpson index based on patient diversity, mono-patient clusters (white), full representation (black). Right 3 columns show row-normalized of frequency of NOA, FR, PR representation in each CD cell subset. Significant differences (Mann-Whitney, alpha=0.05) are marked, triangle NOA vs. PR and circle NOA vs. FR. b. Distribution of Gini-Simpson’s index of patient diversity in FGID (top) and pediCD (bottom) for myeloid cell clusters. c. Sankey plot comparing joined traditional single-level clustering (left) to disease-separated iterative tiered clustering (right). Each line follows each cell as it moves between in the two cluster sets (back bar split based on cluster identity). d. Gini-Simpson index on representation of traditional clusters in each of the separated tiered clusters (i.e., from how many of the higher-level clusters does the deep clustering pull). Calculated separately for FGID (blue) and pediCD (red). e. Similar to **d** but showing the total counts of how many traditional clusters are represented in a single tiered cluster per disease. f. UMAP of combined Myeloid cells: red shows example end clusters from ITC that are split across the traditional-clustering joint-disease UMAP.

**Supplemental Figure 11.**
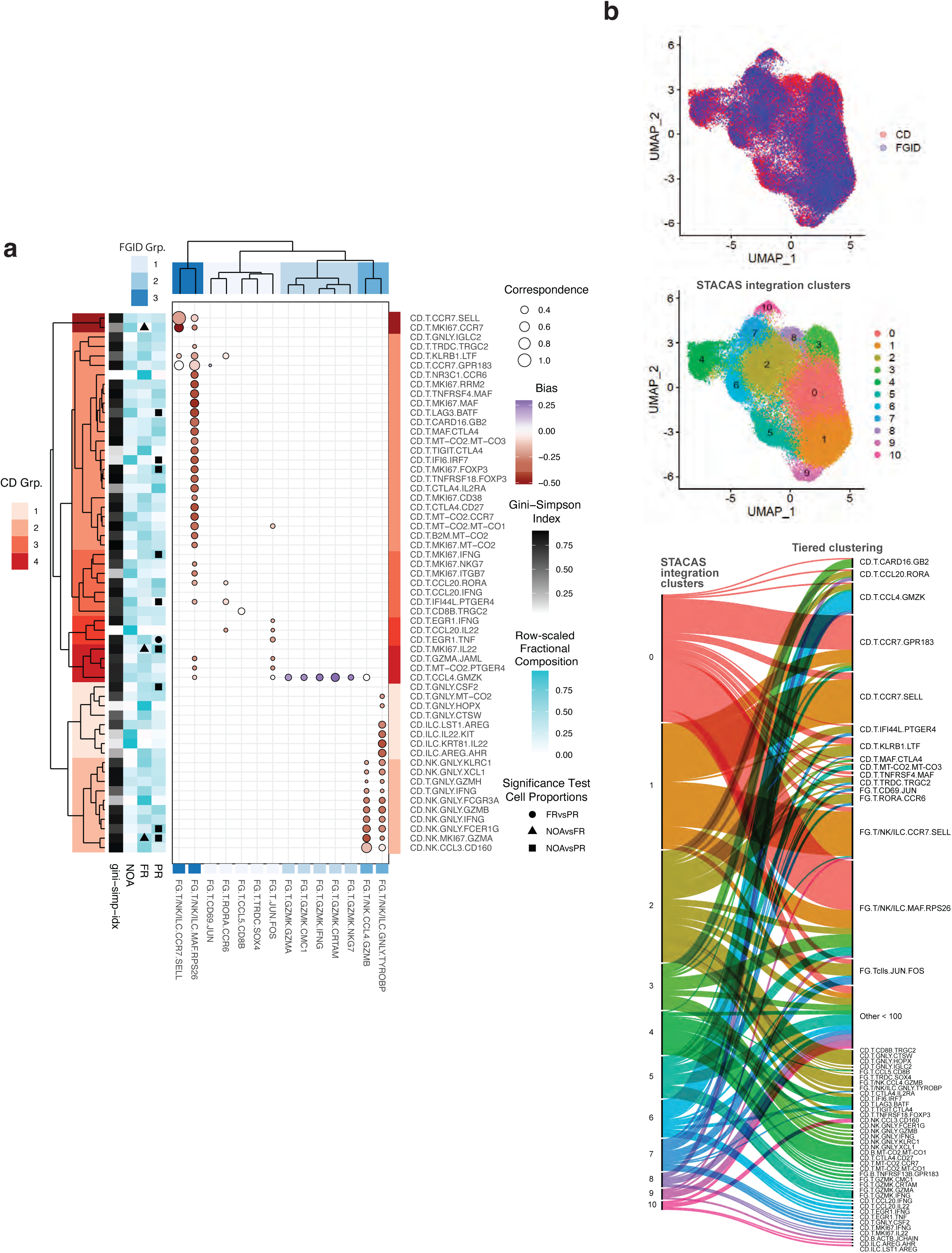
Random Forest classification applied to T cell subsets and integration using STACAS. a. Correspondence between cell subsets from FGID-to-pediCD and pediCD-to-FGID. <u>Top left heatmaps</u>: RF probabilities for each cell averaged over subset to gain probability of each FGID matching onto each pediCD subset (left), and pediCD onto FGID (right). <u>Bubble plot (center)</u>: size = sum(probability matrices) for confidence of predictions, marker color = diff(probability matrices) to show which direction the RF model is more confident on, e.g. more likely for FGID subset to belong to pediCD subset or pediCD subset to belong to FGID subset. Markers are filtered to show the top 10^th^ percentile of correspondence. <u>Dendrograms</u>: separated-tiered clustering on prediction probabilities of FGID (blue) and pediCD (red) using complete linkage with correlation distance metric, clusters are cut at height 0.7 (range 0-1). <u>Heatmap</u>: 1-Gini-Simpson index based on patient diversity, mono-patient clusters (white), full representation (black). Right 3 columns show row-normalized of frequency of NOA, FR, PR representation in each pediCD cell subset. Significant differences (Mann-Whitney, alpha=0.05) are marked, triangle NOA vs. PR and circle NOA vs. FR. b. T cells from the main FGID (n = 29,640 cells) and pediCD (n = 38,031) datasets were integrated using identification of mutual nearest neighbors in a reduced space (reciprocal PCA method) with the STACAS package. UMAP plots show distribution of cells coming from FGID (blue) and pediCD (red) datasets and 11 clusters obtained using Louvain algorithm. Sankey plot shows the contribution of ARBOL clusters to each Louvain cluster in the integrated dataset.

**Supplemental Figure 12.**
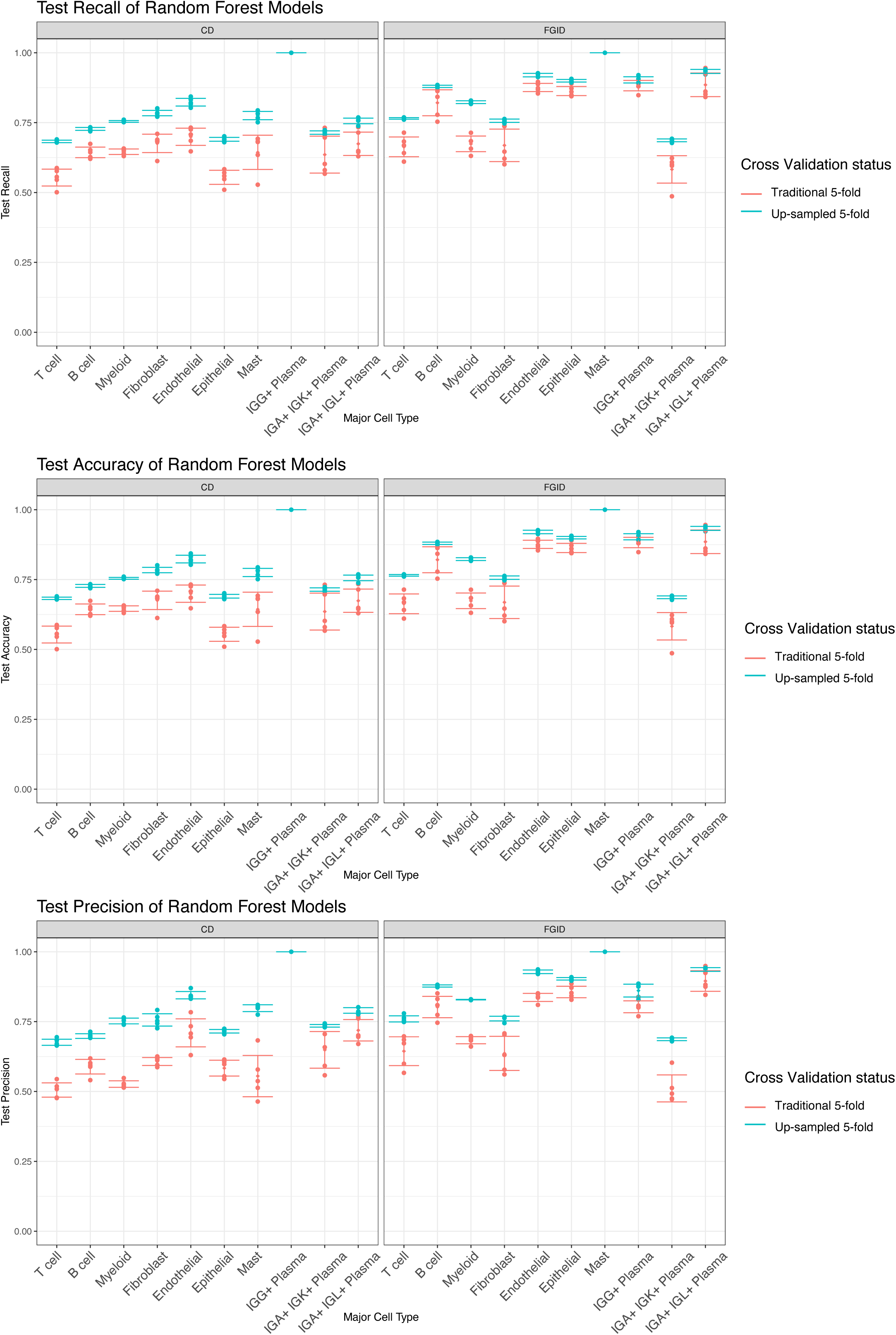
Up-sampling improves Random Forest model test recall, accuracy, and precision.. Points represent the test recall (top left), precision (bottom left), or accuracy (top right) for random forest models. Error bars show the mean and standard deviation of the 5 test scores for each model under two methods of 5-fold cross validation (CV), traditional CV (red) and up-sampled CV (blue). X-axis shows 20 RF models (10 trained on FGID, 10 trained on CD), each trained on a major cell type to predict which ARBOL end-cluster of that major type a provided cell transcriptome belongs to. Recall, accuracy, and precision are calculated on the remaining 20% of the dataset in each of the 5-fold CV. SciKit-Learn’s precision and recall score function https://scikit-learn.org/stable/auto_examples/model_selection/plot_precision_recall.html was used with the “weighted” option to account for multi-class label imbalance. Up-sampled 5-fold cross validation was performed (blue) by up-sampling per label to the largest label size. For up-sampled CV, recall, precision, and accuracy are calculated as normal on the remaining 20% of data. Across major cell types, the accuracy of the RF models improved on average 9.8% when using the up-sampled 5-fold CV in the CD disease condition and improved 6.5% on average across major cell types in the FGID disease condition. Precision increased by 13% in CD and 8.7% in FGID respectively. Recall increased by 9.8% in CD and 6.5% in FGID respectively.

**Supplemental Figure 13:**
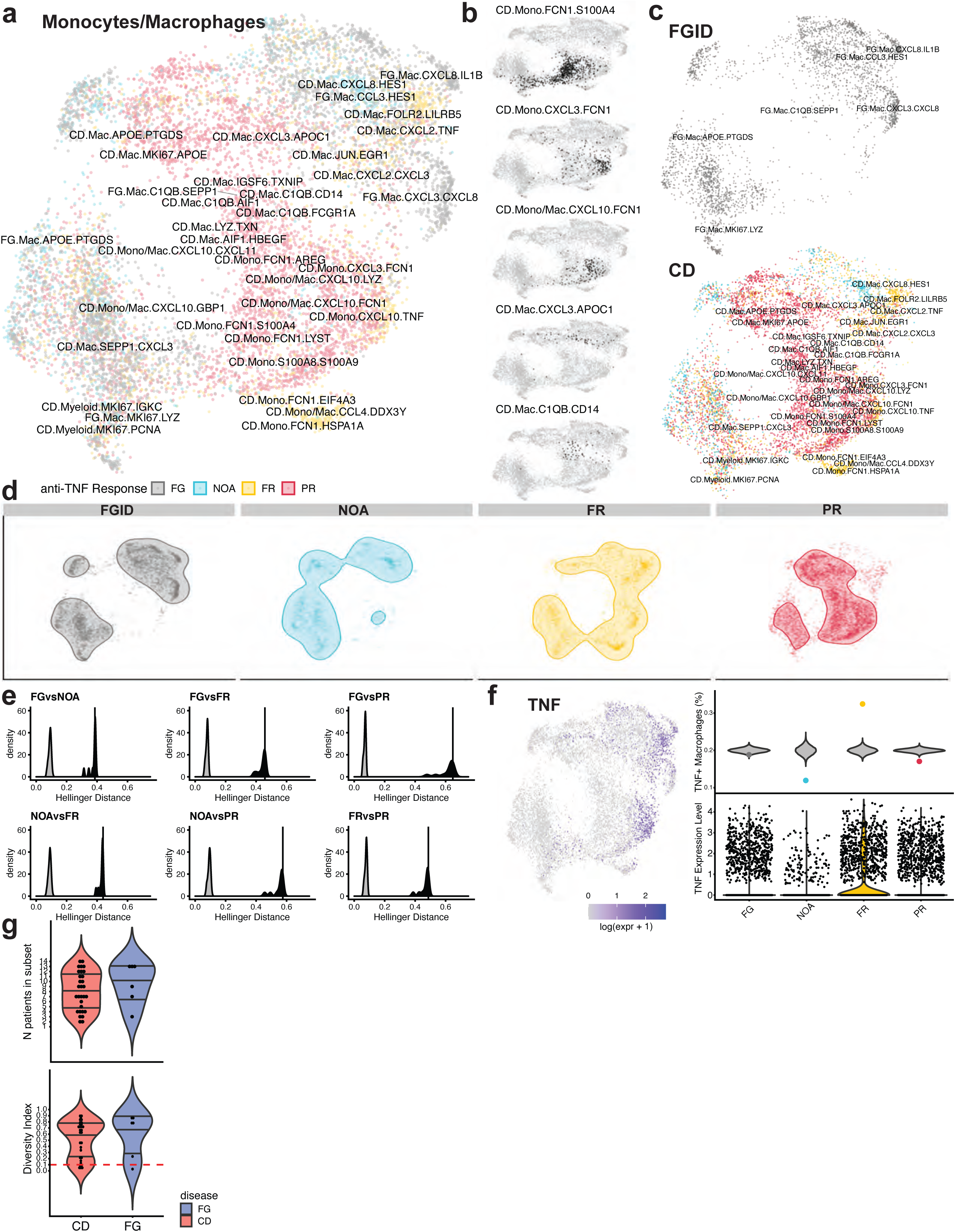
Distinct Distributions of Macrophages Across the pediCD Treatment Response Spectrum Relative to FGID. a. UMAP representation of macrophages (27 patients; 10,134 cells) from FG and pediCD datasets, run across 50 principal components based on 539 genes significantly upregulated (Wilcoxon; p.adj<0.05) in macrophages versus all other cell types and not significantly differentially expressed between FG and pediCD sets. UMAP parameters [min-dist=0.1, N-neighbors=ceiling(sqrt(Ncells)/2)]. Labels are set at median of IQR for each subset. Clinical metadata showing future response to anti-TNF treatment: FGID (grey), NOA (blue), FR (yellow), PR (red). b. Same UMAP as in **a** colored to isolate single subsets. Subsets chosen based on significant Mann-Whitney tests (**Figure 5; Supplemental Figure 7**), (black) cells from subset, (grey) rest of macrophages. c. Same UMAP as in **a** separated into FGID and pediCD. d. Same UMAP as in **a** split into each treatment response group. Shaded area captures 80% most densely populated regions of plot area calculated using 2d KDE estimate from MASS R package. e. Permutation test results using Hellinger distance to measure if 2 conditions are sampled from the same distribution (0 = complete overlap, 1 = no overlap). Hellinger distance is computed with sqrt(1 - sum(sqrt(kde1*kde2))) with a KDE estimation for each condition group calculated across 1000 points uniformly distributed across plot area, with bandwidth selected using ks::Hpi() function. Black distribution shows test statistic varying min-dist parameter with 11 evenly spaced values between 0.01 and 1. Vertical line shows test statistic using UMAP parameters [min-dist=0.1, Neighbors=ceiling(sqrt(Ncells)), Npcs=50, nDim=2]. Grey distribution shows results of 11,000 permutations to treatment response group varied across same min-dist umap parameters between 0.01 and 1. All tests are significant beyond a 0.001 threshold. f. Clockwise from left: UMAP of macrophages with color intensity displaying amount of TNF expression based on ((log(scaledUMI+1)). Plot (top) showing fraction of macrophages expressing TNF with colored dots showing fraction of TNF+ cells within each treatment response group and grey violins showing results of 10,000 permutations of treatment response labels. Violin plot (bottom) of ((log(scaledUMI+1)) TNF expression split on treatment response group. g. Diversity of macrophage clusters in FGID and pediCD: (top) each dot represents a cell subset, y-axis shows how many patients are included within the subset. (bottom) each dot represents a subset, with y position showing (1-Gini-Simpson’s Diversity Index), Subsets below red dashed line set at 0.1 diversity were excluded.

**Supplemental Figure 14.**
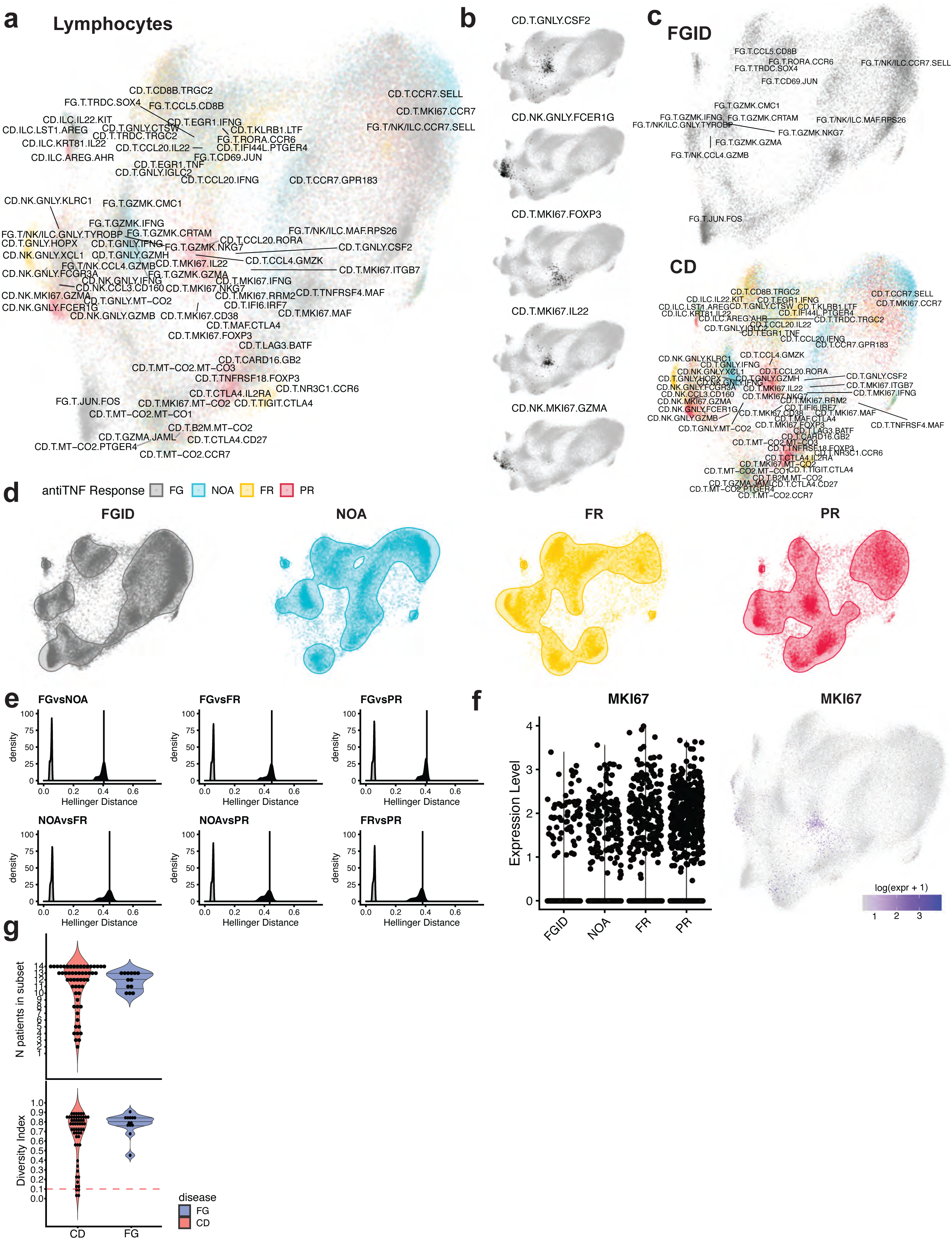
Distinct Distributions of Lymphocytes Across the pediCD Treatment Response Spectrum Relative to FGID. a. UMAP representation of T/NK/ILCs (27 patients; 67,579 cells) from FG and CD datasets, run across 50 principal components based on 345 genes significantly upregulated (Wilcoxon; p.adj<0.05) in lymphocytes versus all other cell types and not significantly differentially expressed between FG and pediCD sets. UMAP parameters [min-dist=0.1, N-neighbors=ceiling(sqrt(Ncells)/2)] Labels are set at median of IQR for each subset. Clinical metadata showing future response to anti-TNF treatment: FGID (grey), NOA (blue), FR (yellow), PR (red). b. Same UMAP as in **a** colored to isolate single subsets. Subsets chosen based on significant Mann-Whitney tests (**Figure 5**), (black) cells from subset, (grey) rest of lymphocytes. c. Same UMAP as in **a** separated into FG and pediCD. d. Same UMAP as in **a** split into each treatment response group. Shaded area captures 80% most densely populated regions of plot area calculated using 2d KDE estimate from MASS R package. e. Permutation test results using Hellinger distance to measure if 2 conditions are sampled from the same distribution (0 = complete overlap, 1 = no overlap). Hellinger distance is computed with sqrt(1 - sum(sqrt(kde1*kde2))) with a KDE estimation for each condition group calculated across 1000 points uniformly distributed across plot area, with bandwidth selected using ks::Hpi() function. Black distribution shows test statistic varying min-dist parameter with 11 evenly spaced values between 0.01 and 1. Vertical line shows test statistic using UMAP parameters [min-dist=0.1, Neighbors=ceiling(sqrt(Ncells)), Npcs=50, nDim=2]. Grey distribution shows results of 11,000 permutations to treatment response group varied across same min-dist umap parameters between 0.01 and 1. All tests are significant beyond a 0.001 threshold. f. Violin plot (left) of ((log(scaledUMI+1)) *MKI67* expression split on treatment response group. UMAP (right) of lymphocytes with color intensity displaying *MKI67* expression based on ((log(scaledUMI+1)) (right). g. Diversity of lymphocyte clusters in FGID and CD: (top) each dot represents a cell subset, y-axis shows how many patients are included within the subset. (bottom) each dot represents a subset, with y position showing (1-Gini-Simpson’s Diversity Index), Subsets below red dashed line set at 0.1 diversity were excluded.

**Supplemental Figure 15.**
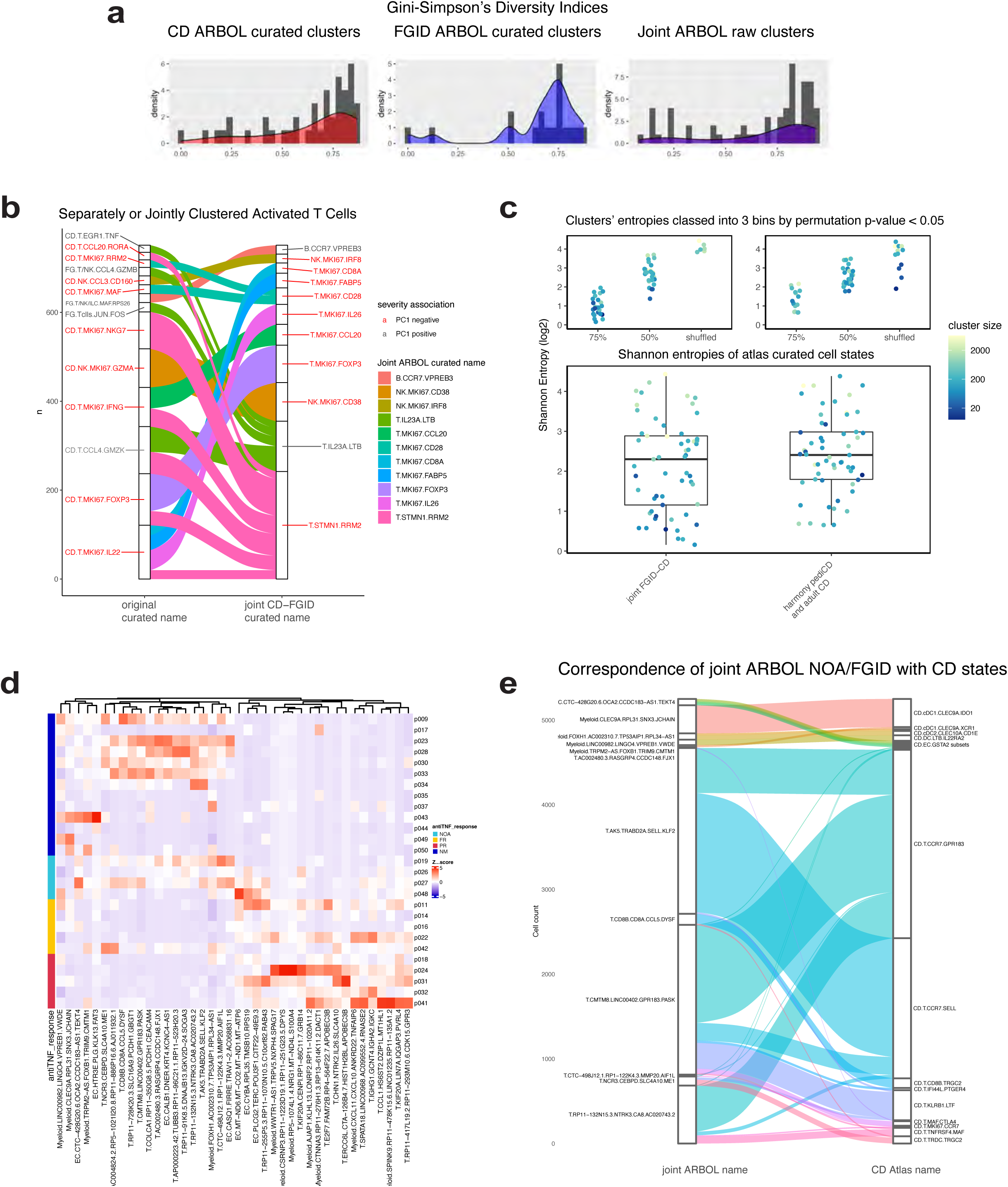
Comparison of Diversity Indices and Entropy of separately- or jointly-clustered T/NK/ILCs from pediCD and FGID patients. a. Distribution of Gini-Simpson’s index of patient diversity in pediCD, FGID, and jointly-clustered T/NK/ILC’s. b. Sankey plot comparing pediCD ARBOL atlas clusters (left) to joint pediCD and FGID ARBOL (right). Each line follows each cell as it moves between in the two cluster sets (back bar split based on cluster identity). Alluvials of 10 cells or less were filtered out. c. Log base 2 Shannon entropies of curated atlas T/NK/ILC cell states in the joint FGID-CD atlas, the harmony-integrated GIMATS + longitudinal PREDICT CD-FGID atlas, and simulated entropies with 75% label preservation, 50% preservation, and random shuffling. d. Cell state compositions per donor associated with joint-PC1 (**Figure 6a**). Note cell state overlap between FGID and NOA CD patients. e. Correspondence of overlapping NOA/FGID cell states with CD-only clusters.

**Supplemental Figure 16.**
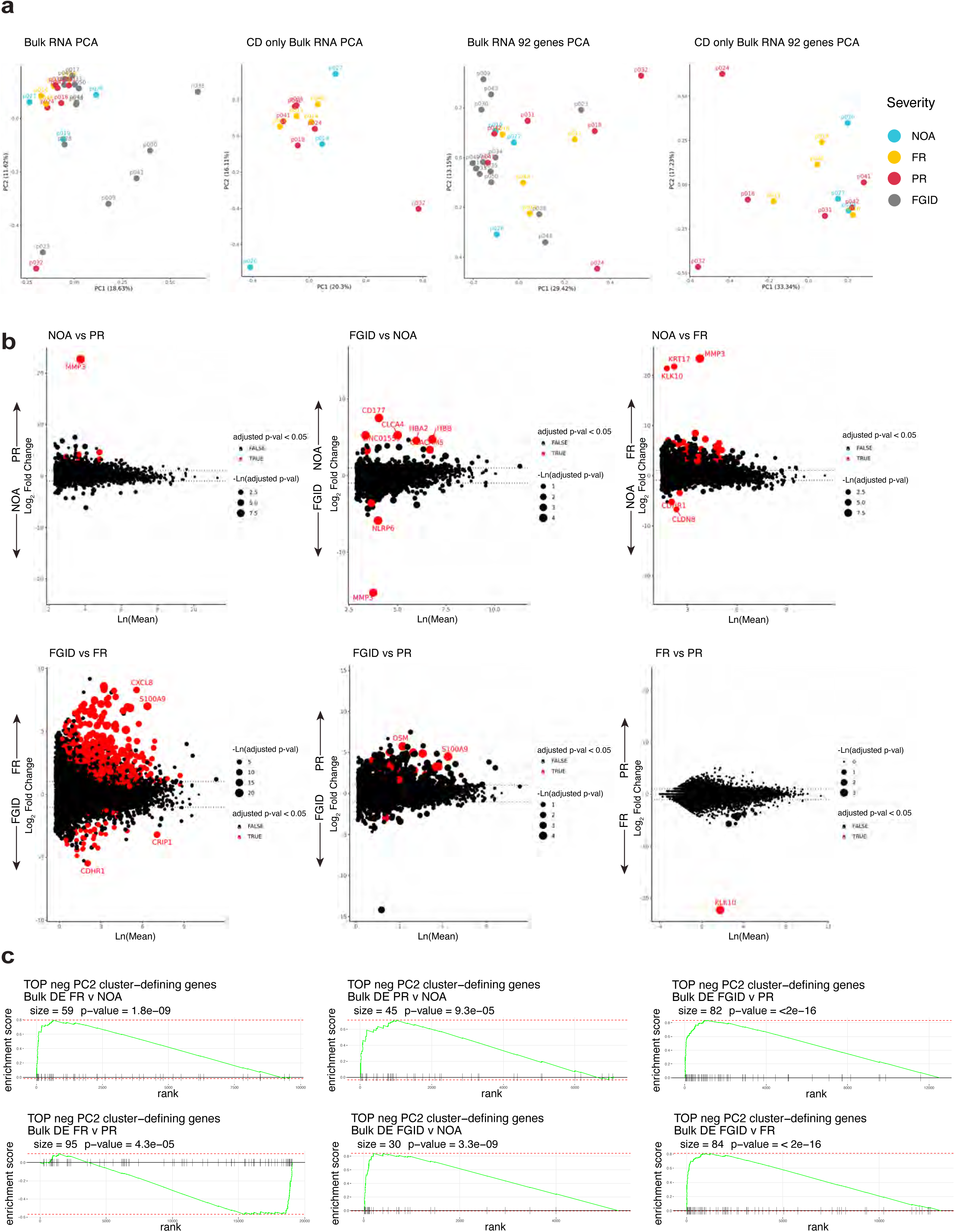
Bulk RNA-seq data from PREDICT cohort is insufficient to recover a disease severity signature, but the 92-gene signature derived from scRNA-seq PREDICT cohort is enriched in severe patients.. a. PCA of Bulk RNA-seq from CD and FGID (gray) biopsies show dominance of sample-level variance including FGID (n = 26) and without FGID samples (n = 13). b. Differential expression between conditions fails to find many significant (FDR < 0.05, red) DE genes, except in FGID (NM) vs. FR. NOA vs FR shows change in KRT17, suggesting bulk RNA seq confounded by variance in epithelial cell dissociation. c. 92 genes are enriched in DE between conditions in bulk RNA-seq data by GSEA.

**Supplemental Figure 17.**
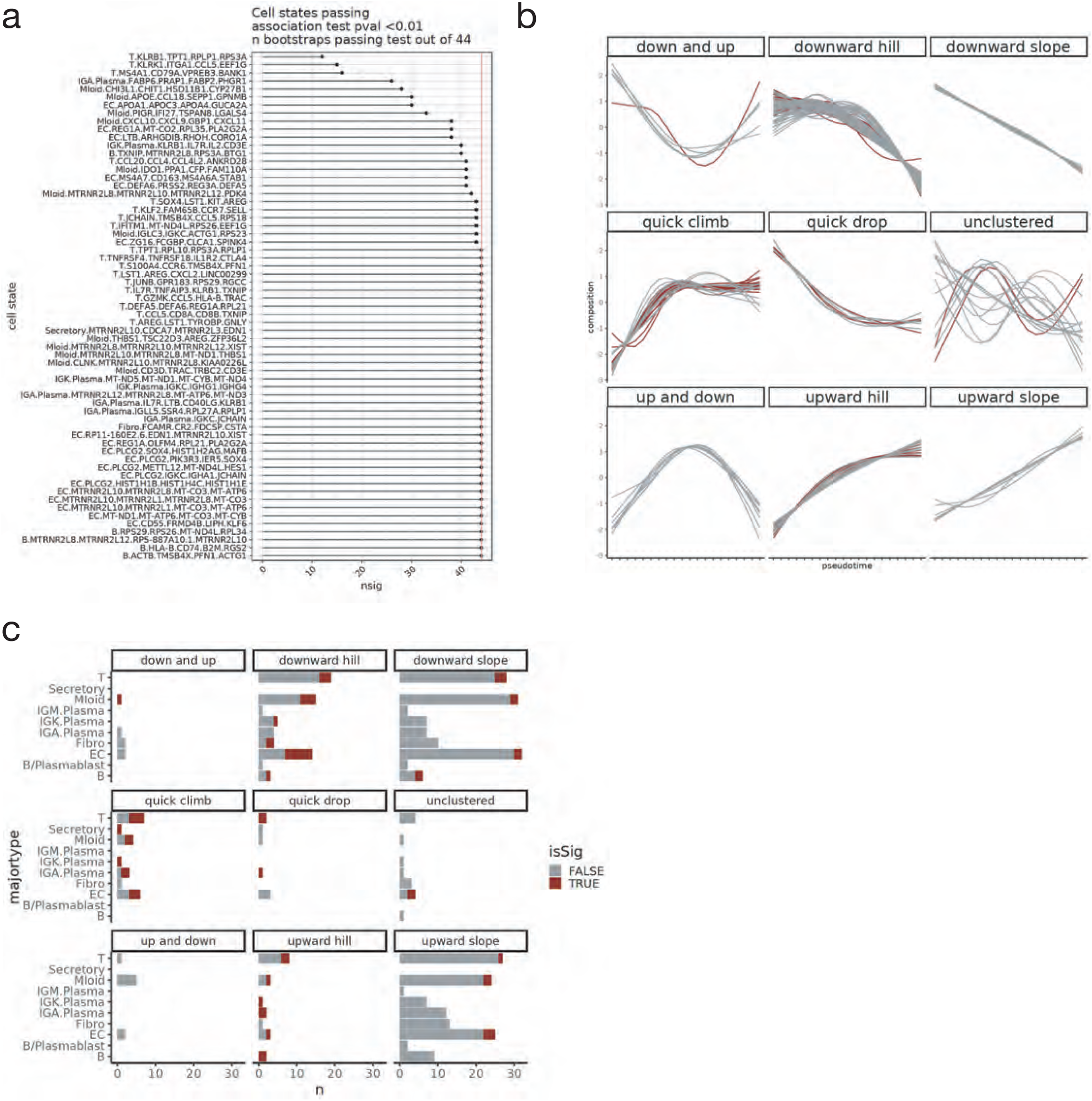
Disease trajectory pseudotime sample-cross validation test and trajectory clustering.. a. Leave-one-out bootstrap test to determine significance of association between cell type composition and disease-pseudotime trajectory. Cell states displayed if association pval < 0.01 and association is robust in at least 10 bootstraps b. Cell state composition GAM fits clustered by trend (Left). Red color highlights cell states for which > 90% of bootstraps are significantly associated with pseudotime across LOOCV. (Right) Count of cell states per major cell type across pseudotime trends, with significant associations highlighted in red. c. Representative significant cell state composition (y axis) trends across pseudotime (x axis) with bootstrapped GAM fits overlaid.

## REFERENCES

Ai, L., Ren, Y., Zhu, M., Lu, S., Qian, Y., Chen, Z., Xu, A., 2021. Synbindin restrains proinflammatory macrophage activation against microbiota and mucosal inflammation during colitis. Gut gutjnl-2020-321094. 10.1136/gutjnl-2020-321094

Andreatta, M., Carmona, S.J., 2021. STACAS: Sub-Type Anchor Correction for Alignment in Seurat to integrate single-cell RNA-seq data. Bioinformatics 37, 882–884. 10.1093/bioinformatics/btaa755

Aschenbrenner, D., Quaranta, M., Banerjee, S., Ilott, N., Jansen, J., Steere, B., Chen, Y.-H., Ho, S., Cox, K., Arancibia-Cárcamo, C.V., Coles, M., Gaffney, E., Travis, S.P., Denson, L., Kugathasan, S., Schmitz, J., Powrie, F., Sansom, S.N., Uhlig, H.H., 2021. Deconvolution of monocyte responses in inflammatory bowel disease reveals an IL-1 cytokine network that regulates IL-23 in genetic and acquired IL-10 resistance. Gut 70, 1023–1036. 10.1136/gutjnl-2020-321731

Atreya, R., Neurath, M.F., Siegmund, B., 2020. Personalizing Treatment in IBD: Hype or Reality in 2020? Can We Predict Response to Anti-TNF? Frontiers in Medicine 7, 517. 10.3389/fmed.2020.00517

Bakken, T.E., Jorstad, N.L., Hu, Q., Lake, B.B., Tian, W., Kalmbach, B.E., Crow, M., Hodge, R.D., Krienen, F.M., Sorensen, S.A., Eggermont, J., Yao, Z., Aevermann, B.D., Aldridge, A.I., Bartlett, A., Bertagnolli, D., Casper, T., Castanon, R.G., Crichton, K., Daigle, T.L., Dalley, R., Dee, N., Dembrow, N., Diep, D., Ding, S.-L., Dong, W., Fang, R., Fischer, S., Goldman, M., Goldy, J., Graybuck, L.T., Herb, B.R., Hou, X., Kancherla, J., Kroll, M., Lathia, K., van Lew, B., Li, Y.E., Liu, C.S., Liu, H., Lucero, J.D., Mahurkar, A., McMillen, D., Miller, J.A., Moussa, M., Nery, J.R., Nicovich, P.R., Niu, S.-Y., Orvis, J., Osteen, J.K., Owen, S., Palmer, C.R., Pham, T., Plongthongkum, N., Poirion, O., Reed, N.M., Rimorin, C., Rivkin, A., Romanow, W.J., Sedeño-Cortés, A.E., Siletti, K., Somasundaram, S., Sulc, J., Tieu, M., Torkelson, A., Tung, H., Wang, X., Xie, F., Yanny, A.M., Zhang, R., Ament, S.A., Behrens, M.M., Bravo, H.C., Chun, J., Dobin, A., Gillis, J., Hertzano, R., Hof, P.R., Höllt, T., Horwitz, G.D., Keene, C.D., Kharchenko, P.V., Ko, A.L., Lelieveldt, B.P., Luo, C., Mukamel, E.A., Pinto-Duarte, A., Preissl, S., Regev, A., Ren, B., Scheuermann, R.H., Smith, K., Spain, W.J., White, O.R., Koch, C., Hawrylycz, M., Tasic, B., Macosko, E.Z., McCarroll, S.A., Ting, J.T., Zeng, H., Zhang, K., Feng, G., Ecker, J.R., Linnarsson, S., Lein, E.S., 2021. Comparative cellular analysis of motor cortex in human, marmoset and mouse. Nature 598, 111–119. 10.1038/s41586-021-03465-8

Banerjee, A., Herring, C.A., Chen, B., Kim, H., Simmons, A.J., Southard-Smith, A.N., Allaman, M.M., White, J.R., Macedonia, M.C., Mckinley, E.T., Ramirez-Solano, M.A., Scoville, E.A., Liu, Q., Wilson, K.T., Coffey, R.J., Washington, M.K., Goettel, J.A., Lau, K.S., 2020. Succinate Produced by Intestinal Microbes Promotes Specification of Tuft Cells to Suppress Ileal Inflammation. Gastroenterology 159, 2101–2115.e5. 10.1053/j.gastro.2020.08.029

Barker, N., van Es, J.H., Kuipers, J., Kujala, P., van den Born, M., Cozijnsen, M., Haegebarth, A., Korving, J., Begthel, H., Peters, P.J., Clevers, H., 2007. Identification of stem cells in small intestine and colon by marker gene Lgr5. Nature 449, 1003–1007. 10.1038/nature06196

Baumgart, D.C., Sandborn, W.J., 2012. Crohn’s disease. The Lancet 380, 1590–1605. 10.1016/S0140-6736(12)60026-9

Betts, B.C., Veerapathran, A., Pidala, J., Yang, H., Horna, P., Walton, K., Cubitt, C.L., Gunawan, S., Lawrence, H.R., Lawrence, N.J., Sebti, S.M., Anasetti, C., 2017. Targeting Aurora kinase A and JAK2 prevents GVHD while maintaining Treg and antitumor CTL function. Sci Transl Med 9, eaai8269. 10.1126/scitranslmed.aai8269

Beumer, J., Puschhof, J., Bauzá-Martinez, J., Martínez-Silgado, A., Elmentaite, R., James, K.R., Ross, A., Hendriks, D., Artegiani, B., Busslinger, G.A., Ponsioen, B., Andersson-Rolf, A., Saftien, A., Boot, C., Kretzschmar, K., Geurts, M.H., Bar-Ephraim, Y.E., Pleguezuelos-Manzano, C., Post, Y., Begthel, H., van der Linden, F., Lopez-Iglesias, C., van de Wetering, W.J., van der Linden, R., Peters, P.J., Heck, A.J.R., Goedhart, J., Snippert, H., Zilbauer, M., Teichmann, S.A., Wu, W., Clevers, H., 2020. High-Resolution mRNA and Secretome Atlas of Human Enteroendocrine Cells. Cell 181, 1291–1306.e19. 10.1016/j.cell.2020.04.036

Bielecki, P., Riesenfeld, S.J., Hütter, J.-C., Torlai Triglia, E., Kowalczyk, M.S., Ricardo-Gonzalez, R.R., Lian, M., Amezcua Vesely, M.C., Kroehling, L., Xu, H., Slyper, M., Muus, C., Ludwig, L.S., Christian, E., Tao, L., Kedaigle, A.J., Steach, H.R., York, A.G., Skadow, M.H., Yaghoubi, P., Dionne, D., Jarret, A., McGee, H.M., Porter, C.B.M., Licona-Limón, P., Bailis, W., Jackson, R., Gagliani, N., Gasteiger, G., Locksley, R.M., Regev, A., Flavell, R.A., 2021. Skin-resident innate lymphoid cells converge on a pathogenic effector state. Nature 592, 128–132. 10.1038/s41586-021-03188-w

Biton, M., Haber, A.L., Rogel, N., Burgin, G., Beyaz, S., Schnell, A., Ashenberg, O., Su, C.-W., Smillie, C., Shekhar, K., Chen, Z., Wu, C., Ordovas-Montanes, J., Alvarez, D., Herbst, R.H., Zhang, M., Tirosh, I., Dionne, D., Nguyen, L.T., Xifaras, M.E., Shalek, A.K., von Andrian, U.H., Graham, D.B., Rozenblatt-Rosen, O., Shi, H.N., Kuchroo, V., Yilmaz, O.H., Regev, A., Xavier, R.J., 2018. T Helper Cell Cytokines Modulate Intestinal Stem Cell Renewal and Differentiation. Cell 175, 1307–1320.e22. 10.1016/j.cell.2018.10.008

Björklund, Å.K., Forkel, M., Picelli, S., Konya, V., Theorell, J., Friberg, D., Sandberg, R., Mjösberg, J., 2016. The heterogeneity of human CD127+ innate lymphoid cells revealed by single-cell RNA sequencing. Nat Immunol 17, 451–460. 10.1038/ni.3368

Black, C.J., Drossman, D.A., Talley, N.J., Ruddy, J., Ford, A.C., 2020. Functional gastrointestinal disorders: advances in understanding and management. The Lancet 396, 1664–1674. 10.1016/S0140-6736(20)32115-2

Blériot, C., Chakarov, S., Ginhoux, F., 2020. Determinants of Resident Tissue Macrophage Identity and Function. Immunity 52, 957–970. 10.1016/j.immuni.2020.05.014

Brulois, K., Rajaraman, A., Szade, A., Nordling, S., Bogoslowski, A., Dermadi, D., Rahman, M., Kiefel, H., O’Hara, E., Koning, J.J., Kawashima, H., Zhou, B., Vestweber, D., Red-Horse, K., Mebius, R.E., Adams, R.H., Kubes, P., Pan, J., Butcher, E.C., 2020. A molecular map of murine lymph node blood vascular endothelium at single cell resolution. Nat Commun 11, 3798. 10.1038/s41467-020-17291-5

Buechler, M.B., Pradhan, R.N., Krishnamurty, A.T., Cox, C., Calviello, A.K., Wang, A.W., Yang, Y.A., Tam, L., Caothien, R., Roose-Girma, M., Modrusan, Z., Arron, J.R., Bourgon, R., Müller, S., Turley, S.J., 2021. Cross-tissue organization of the fibroblast lineage. Nature 1–5. 10.1038/s41586-021-03549-5

Buisine, M.P., Desreumaux, P., Leteurtre, E., Copin, M.C., Colombel, J.F., Porchet, N., Aubert, J.P., 2001. Mucin gene expression in intestinal epithelial cells in Crohn’s disease. Gut 49, 544–551. 10.1136/gut.49.4.544

Cao, J., Spielmann, M., Qiu, X., Huang, X., Ibrahim, D.M., Hill, A.J., Zhang, F., Mundlos, S., Christiansen, L., Steemers, F.J., Trapnell, C., Shendure, J., 2019. The single-cell transcriptional landscape of mammalian organogenesis. Nature 566, 496–502. 10.1038/s41586-019-0969-x

Cappello, M., Morreale, G.C., 2016. The Role of Laboratory Tests in Crohn’s Disease. Clin Med Insights Gastroenterol 9, 51–62. 10.4137/CGast.S38203

Castro-Dopico, T., Fleming, A., Dennison, T.W., Ferdinand, J.R., Harcourt, K., Stewart, B.J., Cader, Z., Tuong, Z.K., Jing, C., Lok, L.S.C., Mathews, R.J., Portet, A., Kaser, A., Clare, S., Clatworthy, M.R., 2020. GM-CSF Calibrates Macrophage Defense and Wound Healing Programs during Intestinal Infection and Inflammation. Cell Reports 32, 107857. 10.1016/j.celrep.2020.107857

Catalan-Serra, I., Sandvik, A.K., Bruland, T., Andreu-Ballester, J.C., 2017. Gammadelta T Cells in Crohn’s Disease: A New Player in the Disease Pathogenesis? Journal of Crohn’s and Colitis 11, 1135–1145. 10.1093/ecco-jcc/jjx039

Chang, J.T., 2020. Pathophysiology of Inflammatory Bowel Diseases. New England Journal of Medicine 383, 2652–2664. 10.1056/NEJMra2002697

Cherrier, D.E., Serafini, N., Di Santo, J.P., 2018. Innate Lymphoid Cell Development: A T Cell Perspective. Immunity 48, 1091–1103. 10.1016/j.immuni.2018.05.010

Cima, I., Corazza, N., Dick, B., Fuhrer, A., Herren, S., Jakob, S., Ayuni, E., Mueller, C., Brunner, T., 2004. Intestinal Epithelial Cells Synthesize Glucocorticoids and Regulate T Cell Activation. J Exp Med 200, 1635–1646. 10.1084/jem.20031958

Cohen, L.J., Cho, J.H., Gevers, D., Chu, H., 2019. Genetic Factors and the Intestinal Microbiome Guide Development of Microbe-Based Therapies for Inflammatory Bowel Diseases. Gastroenterology 156, 2174–2189. 10.1053/j.gastro.2019.03.017

Combes, A.J., Samad, B., Tsui, J., Chew, N.W., Yan, P., Reeder, G.C., Kushnoor, D., Shen, A., Davidson, B., Barczak, A.J., Adkisson, M., Edwards, A., Naser, M., Barry, K.C., Courau, T., Hammoudi, T., Argüello, R.J., Rao, A.A., Olshen, A.B., Cai, C., Zhan, J., Davis, K.C., Kelley, R.K., Chapman, J.S., Atreya, C.E., Patel, A., Daud, A.I., Ha, P., Diaz, A.A., Kratz, J.R., Collisson, E.A., Fragiadakis, G.K., Erle, D.J., Boissonnas, A., Asthana, S., Chan, V., Krummel, M.F., Spitzer, M., Fong, L., Nelson, A., Kumar, R., Lee, J., Burra, A., Hsu, J., Hackett, C., Tolentino, K., Sjarif, J., Johnson, P., Shao, E., Abrau, D., Lupin, L., Shaw, C., Collins, Z., Lea, T., Corvera, C., Nakakura, E., Carnevale, J., Alvarado, M., Loo, K., Chen, L., Chow, M., Grandis, J., Ryan, W., El-Sayed, I., Jablons, D., Woodard, G., Meng, M.W., Porten, S.P., Okada, H., Tempero, M., Ko, A., Kirkwood, K., Vandenberg, S., Guevarra, D., Oropeza, E., Cyr, C., Glenn, P., Bolen, J., Morton, A., Eckalbar, W., 2022. Discovering dominant tumor immune archetypes in a pan-cancer census. Cell 185, 184–203.e19. 10.1016/j.cell.2021.12.004

Corridoni, D., Antanaviciute, A., Gupta, T., Fawkner-Corbett, D., Aulicino, A., Jagielowicz, M., Parikh, K., Repapi, E., Taylor, S., Ishikawa, D., Hatano, R., Yamada, T., Xin, W., Slawinski, H., Bowden, R., Napolitani, G., Brain, O., Morimoto, C., Koohy, H., Simmons, A., 2020a. Single-cell atlas of colonic CD8+ T cells in ulcerative colitis. Nat Med 26, 1480–1490. 10.1038/s41591-020-1003-4

Corridoni, D., Chapman, T., Antanaviciute, A., Satsangi, J., Simmons, A., 2020b. Inflammatory Bowel Disease Through the Lens of Single-cell RNA-seq Technologies. Inflammatory Bowel Diseases 26, 1658–1668. 10.1093/ibd/izaa089

Cyster, J.G., Allen, C.D.C., 2019. B Cell Responses: Cell Interaction Dynamics and Decisions. Cell 177, 524–540. 10.1016/j.cell.2019.03.016

Dann, E., Henderson, N.C., Teichmann, S.A., Morgan, M.D., Marioni, J.C., 2022. Differential abundance testing on single-cell data using k-nearest neighbor graphs. Nat Biotechnol 40, 245–253. 10.1038/s41587-021-01033-z

Das, A., Heesters, B.A., Bialas, A., O’Flynn, J., Rifkin, I.R., Ochando, J., Mittereder, N., Carlesso, G., Herbst, R., Carroll, M.C., 2017. Follicular Dendritic Cell Activation by TLR Ligands Promotes Autoreactive B Cell Responses. Immunity 46, 106–119. 10.1016/j.immuni.2016.12.014

Davidson, S., Coles, M., Thomas, T., Kollias, G., Ludewig, B., Turley, S., Brenner, M., Buckley, C.D., 2021. Fibroblasts as immune regulators in infection, inflammation and cancer. Nature Reviews Immunology 1–14. 10.1038/s41577-021-00540-z

D’Haens, G.R., Deventer, S. van, 2021. 25 years of anti-TNF treatment for inflammatory bowel disease: lessons from the past and a look to the future. Gut 70, 1396–1405. 10.1136/gutjnl-2019-320022

Digby-Bell, J.L., Atreya, R., Monteleone, G., Powell, N., 2020. Interrogating host immunity to predict treatment response in inflammatory bowel disease. Nat Rev Gastroenterol Hepatol 17, 9–20. 10.1038/s41575-019-0228-5

Dominguez-Sola, D., Victora, G.D., Ying, C.Y., Phan, R.T., Saito, M., Nussenzweig, M.C., Dalla-Favera, R., 2012. The proto-oncogene MYC is required for selection in the germinal center and cyclic reentry. Nat Immunol 13, 1083–1091. 10.1038/ni.2428

Dovrolis, N., Michalopoulos, G., Theodoropoulos, G.E., Arvanitidis, K., Kolios, G., Sechi, L.A., Eliopoulos, A.G., Gazouli, M., 2020. The Interplay between Mucosal Microbiota Composition and Host Gene-Expression is Linked with Infliximab Response in Inflammatory Bowel Diseases. Microorganisms 8, 438. 10.3390/microorganisms8030438

Drokhlyansky, E., Smillie, C.S., Wittenberghe, N.V., Ericsson, M., Griffin, G.K., Dionne, D., Cuoco, M.S., Goder-Reiser, M.N., Sharova, T., Aguirre, A.J., Boland, G.M., Graham, D., Rozenblatt-Rosen, O., Xavier, R.J., Regev, A., 2019. The enteric nervous system of the human and mouse colon at a single-cell resolution. bioRxiv 746743. 10.1101/746743

Dutertre, C.-A., Becht, E., Irac, S.E., Khalilnezhad, A., Narang, V., Khalilnezhad, S., Ng, P.Y., van den Hoogen, L.L., Leong, J.Y., Lee, B., Chevrier, M., Zhang, X.M., Yong, P.J.A., Koh, G., Lum, J., Howland, S.W., Mok, E., Chen, J., Larbi, A., Tan, H.K.K., Lim, T.K.H., Karagianni, P., Tzioufas, A.G., Malleret, B., Brody, J., Albani, S., van Roon, J., Radstake, T., Newell, E.W., Ginhoux, F., 2019. Single-Cell Analysis of Human Mononuclear Phagocytes Reveals Subset-Defining Markers and Identifies Circulating Inflammatory Dendritic Cells. Immunity 51, 573–589.e8. 10.1016/j.immuni.2019.08.008

Dwyer, D.F., Ordovas-Montanes, J., Allon, S.J., Buchheit, K.M., Vukovic, M., Derakhshan, T., Feng, C., Lai, J., Hughes, T.K., Nyquist, S.K., Giannetti, M.P., Berger, B., Bhattacharyya, N., Roditi, R.E., Katz, H.R., Nawijn, M.C., Berg, M., van den Berge, M., Laidlaw, T.M., Shalek, A.K., Barrett, N.A., Boyce, J.A., 2021. Human airway mast cells proliferate and acquire distinct inflammation-driven phenotypes during type 2 inflammation. Sci Immunol 6, eabb7221. 10.1126/sciimmunol.abb7221

Elmentaite, R., Ross, A.D.B., Roberts, K., James, K.R., Ortmann, D., Gomes, T., Nayak, K., Tuck, L., Pritchard, S., Bayraktar, O.A., Heuschkel, R., Vallier, L., Teichmann, S.A., Zilbauer, M., 2020. Single-Cell Sequencing of Developing Human Gut Reveals Transcriptional Links to Childhood Crohn’s Disease. Developmental Cell 55, 771–783.e5. 10.1016/j.devcel.2020.11.010

Franzosa, E.A., Sirota-Madi, A., Avila-Pacheco, J., Fornelos, N., Haiser, H.J., Reinker, S., Vatanen, T., Hall, A.B., Mallick, H., McIver, L.J., Sauk, J.S., Wilson, R.G., Stevens, B.W., Scott, J.M., Pierce, K., Deik, A.A., Bullock, K., Imhann, F., Porter, J.A., Zhernakova, A., Fu, J., Weersma, R.K., Wijmenga, C., Clish, C.B., Vlamakis, H., Huttenhower, C., Xavier, R.J., 2019. Gut microbiome structure and metabolic activity in inflammatory bowel disease. Nat Microbiol 4, 293–305. 10.1038/s41564-018-0306-4

French, A.R., Sjölin, H., Kim, S., Koka, R., Yang, L., Young, D.A., Cerboni, C., Tomasello, E., Ma, A., Vivier, E., Kärre, K., Yokoyama, W.M., 2006. DAP12 Signaling Directly Augments Proproliferative Cytokine Stimulation of NK Cells during Viral Infections. The Journal of Immunology 177, 4981–4990. 10.4049/jimmunol.177.8.4981

Friedrich, M., Pohin, M., Powrie, F., 2019. Cytokine Networks in the Pathophysiology of Inflammatory Bowel Disease. Immunity 50, 992–1006. 10.1016/j.immuni.2019.03.017

Furey, T.S., Sethupathy, P., Sheikh, S.Z., 2019. Redefining the IBDs using genome-scale molecular phenotyping. Nat Rev Gastroenterol Hepatol 16, 296–311. 10.1038/s41575-019-0118-x

Furlan, S.N., Watkins, B., Tkachev, V., Flynn, R., Cooley, S., Ramakrishnan, S., Singh, K., Giver, C., Hamby, K., Stempora, L., Garrett, A., Chen, J., Betz, K.M., Ziegler, C.G.K., Tharp, G.K., Bosinger, S.E., Promislow, D.E.L., Miller, J.S., Waller, E.K., Blazar, B.R., Kean, L.S., 2015. Transcriptome analysis of GVHD reveals aurora kinase A as a targetable pathway for disease prevention. Science Translational Medicine 7, 315ra191–315ra191. 10.1126/scitranslmed.aad3231

Gomariz, A., Helbling, P.M., Isringhausen, S., Suessbier, U., Becker, A., Boss, A., Nagasawa, T., Paul, G., Goksel, O., Székely, G., Stoma, S., Nørrelykke, S.F., Manz, M.G., Nombela-Arrieta, C., 2018. Quantitative spatial analysis of haematopoiesis-regulating stromal cells in the bone marrow microenvironment by 3D microscopy. Nat Commun 9, 2532. 10.1038/s41467-018-04770-z

Graham, D.B., Xavier, R.J., 2020. Pathway paradigms revealed from the genetics of inflammatory bowel disease. Nature 578, 527–539. 10.1038/s41586-020-2025-2

Guilliams, M., Mildner, A., Yona, S., 2018. Developmental and Functional Heterogeneity of Monocytes. Immunity 49, 595–613. 10.1016/j.immuni.2018.10.005

Haberman, Y., Tickle, T.L., Dexheimer, P.J., Kim, M.-O., Tang, D., Karns, R., Baldassano, R.N., Noe, J.D., Rosh, J., Markowitz, J., Heyman, M.B., Griffiths, A.M., Crandall, W.V., Mack, D.R., Baker, S.S., Huttenhower, C., Keljo, D.J., Hyams, J.S., Kugathasan, S., Walters, T.D., Aronow, B., Xavier, R.J., Gevers, D., Denson, L.A., 2014. Pediatric Crohn disease patients exhibit specific ileal transcriptome and microbiome signature. J Clin Invest 124, 3617–3633. 10.1172/JCI75436

Hafemeister, C., Satija, R., 2019. Normalization and variance stabilization of single-cell RNA-seq data using regularized negative binomial regression. Genome Biology 20, 296. 10.1186/s13059-019-1874-1

Hao, Y., Hao, S., Andersen-Nissen, E., Mauck, W.M., Zheng, S., Butler, A., Lee, M.J., Wilk, A.J., Darby, C., Zager, M., Hoffman, P., Stoeckius, M., Papalexi, E., Mimitou, E.P., Jain, J., Srivastava, A., Stuart, T., Fleming, L.M., Yeung, B., Rogers, A.J., McElrath, J.M., Blish, C.A., Gottardo, R., Smibert, P., Satija, R., 2021. Integrated analysis of multimodal single-cell data. Cell. 10.1016/j.cell.2021.04.048

Heesters, B.A., Chatterjee, P., Kim, Y.-A., Gonzalez, S.F., Kuligowski, M.P., Kirchhausen, T., Carroll, M.C., 2013. Endocytosis and Recycling of Immune Complexes by Follicular Dendritic Cells Enhances B Cell Antigen Binding and Activation. Immunity 38, 1164–1175. 10.1016/j.immuni.2013.02.023

Hie, B., Bryson, B., Berger, B., 2019. Efficient integration of heterogeneous single-cell transcriptomes using Scanorama. Nat Biotechnol 37, 685–691. 10.1038/s41587-019-0113-3

Hie, B., Peters, J., Nyquist, S.K., Shalek, A.K., Berger, B., Bryson, B.D., 2020. Computational Methods for Single-Cell RNA Sequencing. Annu. Rev. Biomed. Data Sci. 3, 339–364. 10.1146/annurev-biodatasci-012220-100601

Hoytema van Konijnenburg, D.P., Reis, B.S., Pedicord, V.A., Farache, J., Victora, G.D., Mucida, D., 2017. Intestinal Epithelial and Intraepithelial T Cell Crosstalk Mediates a Dynamic Response to Infection. Cell 171, 783–794.e13. 10.1016/j.cell.2017.08.046

Huang, B., Chen, Z., Geng, L., Wang, J., Liang, H., Cao, Y., Chen, H., Huang, W., Su, M., Wang, Hanqing, Xu, Y., Liu, Y., Lu, B., Xian, H., Li, Huiwen, Li, Huilin, Ren, L., Xie, J., Ye, L., Wang, Hongli, Zhao, J., Chen, P., Zhang, L., Zhao, S., Zhang, T., Xu, B., Che, D., Si, W., Gu, X., Zeng, L., Wang, Y., Li, D., Zhan, Y., Delfouneso, D., Lew, A.M., Cui, J., Tang, W.H., Zhang, Yan, Gong, S., Bai, F., Yang, M., Zhang, Yuxia, 2019. Mucosal Profiling of Pediatric-Onset Colitis and IBD Reveals Common Pathogenics and Therapeutic Pathways. Cell 179, 1160–1176.e24. 10.1016/j.cell.2019.10.027

Hyams, J.S., Di Lorenzo, C., Saps, M., Shulman, R.J., Staiano, A., van Tilburg, M., 2016. Childhood Functional Gastrointestinal Disorders: Child/Adolescent. Gastroenterology, Rome IV - Functional GI Disorders: Disorders of Gut-Brain Interaction 150, 1456–1468.e2. 10.1053/j.gastro.2016.02.015

Hyams, J.S., Ferry, G.D., Mandel, F.S., Gryboski, J.D., Kibort, P.M., Kirschner, B.S., Griffiths, A.M., Katz, A.J., Grand, R.J., Boyle, J.T., Michener, W.M., Levy, J.S., Lesser, M.L., 1991. Development and Validation of a Pediatric Crohn’s Disease Activity Index. Journal of Pediatric Gastroenterology and Nutrition 12, 439.

Jaeger, N., Gamini, R., Cella, M., Schettini, J.L., Bugatti, M., Zhao, S., Rosadini, C.V., Esaulova, E., Di Luccia, B., Kinnett, B., Vermi, W., Artyomov, M.N., Wynn, T.A., Xavier, R.J., Jelinsky, S.A., Colonna, M., 2021. Single-cell analyses of Crohn’s disease tissues reveal intestinal intraepithelial T cells heterogeneity and altered subset distributions. Nat Commun 12, 1921. 10.1038/s41467-021-22164-6

Jain, U., Heul, A.M.V., Xiong, S., Gregory, M.H., Demers, E.G., Kern, J.T., Lai, C.-W., Muegge, B.D., Barisas, D.A.G., Leal-Ekman, J.S., Deepak, P., Ciorba, M.A., Liu, T.-C., Hogan, D.A., Debbas, P., Braun, J., McGovern, D.P.B., Underhill, D.M., Stappenbeck, T.S., 2021. Debaryomyces is enriched in Crohn’s disease intestinal tissue and impairs healing in mice. Science 371, 1154–1159. 10.1126/science.abd0919

James, K.R., Gomes, T., Elmentaite, R., Kumar, N., Gulliver, E.L., King, H.W., Stares, M.D., Bareham, B.R., Ferdinand, J.R., Petrova, V.N., Polański, K., Forster, S.C., Jarvis, L.B., Suchanek, O., Howlett, S., James, L.K., Jones, J.L., Meyer, K.B., Clatworthy, M.R., Saeb-Parsy, K., Lawley, T.D., Teichmann, S.A., 2020. Distinct microbial and immune niches of the human colon. Nat Immunol 21, 343–353. 10.1038/s41590-020-0602-z

Kaczorowski, K.J., Shekhar, K., Nkulikiyimfura, D., Dekker, C.L., Maecker, H., Davis, M.M., Chakraborty, A.K., Brodin, P., 2017. Continuous immunotypes describe human immune variation and predict diverse responses. PNAS 114, E6097–E6106. 10.1073/pnas.1705065114

Kinchen, J., Chen, H.H., Parikh, K., Antanaviciute, A., Jagielowicz, M., Fawkner-Corbett, D., Ashley, N., Cubitt, L., Mellado-Gomez, E., Attar, M., Sharma, E., Wills, Q., Bowden, R., Richter, F.C., Ahern, D., Puri, K.D., Henault, J., Gervais, F., Koohy, H., Simmons, A., 2018. Structural Remodeling of the Human Colonic Mesenchyme in Inflammatory Bowel Disease. Cell 175, 372–386.e17. 10.1016/j.cell.2018.08.067

Kobayashi, T., Siegmund, B., Le Berre, C., Wei, S.C., Ferrante, M., Shen, B., Bernstein, C.N., Danese, S., Peyrin-Biroulet, L., Hibi, T., 2020. Ulcerative colitis. Nat Rev Dis Primers 6, 1–20. 10.1038/s41572-020-0205-x

Korotkevich, G., Sukhov, V., Budin, N., Shpak, B., Artyomov, M.N., Sergushichev, A., 2021. Fast gene set enrichment analysis. 10.1101/060012

Korsunsky, I., Millard, N., Fan, J., Slowikowski, K., Zhang, F., Wei, K., Baglaenko, Y., Brenner, M., Loh, P., Raychaudhuri, S., 2019. Fast, sensitive and accurate integration of single-cell data with Harmony. Nat Methods 16, 1289–1296. 10.1038/s41592-019-0619-0

Kugathasan, S., Denson, L.A., Walters, T.D., Kim, M.-O., Marigorta, U.M., Schirmer, M., Mondal, K., Liu, C., Griffiths, A., Noe, J.D., Crandall, W.V., Snapper, S., Rabizadeh, S., Rosh, J.R., Shapiro, J.M., Guthery, S., Mack, D.R., Kellermayer, R., Kappelman, M.D., Steiner, S., Moulton, D.E., Keljo, D., Cohen, S., Oliva-Hemker, M., Heyman, M.B., Otley, A.R., Baker, S.S., Evans, J.S., Kirschner, B.S., Patel, A.S., Ziring, D., Trapnell, B.C., Sylvester, F.A., Stephens, M.C., Baldassano, R.N., Markowitz, J.F., Cho, J., Xavier, R.J., Huttenhower, C., Aronow, B.J., Gibson, G., Hyams, J.S., Dubinsky, M.C., 2017. Prediction of complicated disease course for children newly diagnosed with Crohn’s disease: a multicentre inception cohort study. Lancet 389, 1710–1718. 10.1016/S0140-6736(17)30317-3

La Manno, G., Siletti, K., Furlan, A., Gyllborg, D., Vinsland, E., Mossi Albiach, A., Mattsson Langseth, C., Khven, I., Lederer, A.R., Dratva, L.M., Johnsson, A., Nilsson, M., Lönnerberg, P., Linnarsson, S., 2021. Molecular architecture of the developing mouse brain. Nature 596, 92–96. 10.1038/s41586-021-03775-x

Lampen, A., Meyer, S., Arnhold, T., Nau, H., 2000. Metabolism of vitamin A and its active metabolite all-trans-retinoic acid in small intestinal enterocytes. J Pharmacol Exp Ther 295, 979–985.

Lanier, L.L., 2001. On guard—activating NK cell receptors. Nat Immunol 2, 23–27. 10.1038/83130

Lanier, L.L., Corliss, B., Wu, J., Phillips, J.H., 1998. Association of DAP12 with Activating CD94/NKG2C NK Cell Receptors. Immunity 8, 693–701. 10.1016/S1074-7613(00)80574-9

Leach, S.T., Yang, Z., Messina, I., Song, C., Geczy, C.L., Cunningham, A.M., Day, A.S., 2007. Serum and mucosal S100 proteins, calprotectin (S100A8/S100A9) and S100A12, are elevated at diagnosis in children with inflammatory bowel disease. Scandinavian Journal of Gastroenterology 42, 1321–1331. 10.1080/00365520701416709

Leeb, S.N., Vogl, D., Gunckel, M., Kiessling, S., Falk, W., Göke, M., Schölmerich, J., Gelbmann, C.M., Rogler, G., 2003. Reduced migration of fibroblasts in inflammatory bowel disease: role of inflammatory mediators and focal adhesion kinase. Gastroenterology 125, 1341–1354. 10.1016/j.gastro.2003.07.004

Leonard, N., Hourihane, D.O., Whelan, A., 1995. Neuroproliferation in the mucosa is a feature of coeliac disease and Crohn’s disease. Gut 37, 763–765. 10.1136/gut.37.6.763

Levine, A., Griffiths, A., Markowitz, J., Wilson, D.C., Turner, D., Russell, R.K., Fell, J., Ruemmele, F.M., Walters, T., Sherlock, M., Dubinsky, M., Hyams, J.S., 2011. Pediatric modification of the Montreal classification for inflammatory bowel disease: the Paris classification. Inflamm Bowel Dis 17, 1314–1321. 10.1002/ibd.21493

Lilja, I., Gustafson-Svärd, C., Franzén, L., Sjödahl, R., 2000. Tumor Necrosis Factor-Alpha in Ileal Mast Cells in Patients with Crohn’s Disease. DIG 61, 68–76. 10.1159/000007737

Limon, J.J., Tang, J., Li, D., Wolf, A.J., Michelsen, K.S., Funari, V., Gargus, M., Nguyen, C., Sharma, P., Maymi, V.I., Iliev, I.D., Skalski, J.H., Brown, J., Landers, C., Borneman, J., Braun, J., Targan, S.R., McGovern, D.P.B., Underhill, D.M., 2019. Malassezia Is Associated with Crohn’s Disease and Exacerbates Colitis in Mouse Models. Cell Host & Microbe 25, 377–388.e6. 10.1016/j.chom.2019.01.007

Lindemans, C.A., Calafiore, M., Mertelsmann, A.M., O’Connor, M.H., Dudakov, J.A., Jenq, R.R., Velardi, E., Young, L.F., Smith, O.M., Lawrence, G., Ivanov, J.A., Fu, Y.-Y., Takashima, S., Hua, G., Martin, M.L., O’Rourke, K.P., Lo, Y.-H., Mokry, M., Romera-Hernandez, M., Cupedo, T., Dow, L.E., Nieuwenhuis, E.E., Shroyer, N.F., Liu, C., Kolesnick, R., van den Brink, M.R.M., Hanash, A.M., 2015. Interleukin-22 promotes intestinal-stem-cell-mediated epithelial regeneration. Nature 528, 560–564. 10.1038/nature16460

Love, M.I., Huber, W., Anders, S., 2014. Moderated estimation of fold change and dispersion for RNA-seq data with DESeq2. Genome Biology 15, 550. 10.1186/s13059-014-0550-8

Lucas, C., Wong, P., Klein, J., Castro, T.B.R., Silva, J., Sundaram, M., Ellingson, M.K., Mao, T., Oh, J.E., Israelow, B., Takahashi, T., Tokuyama, M., Lu, P., Venkataraman, A., Park, A., Mohanty, S., Wang, H., Wyllie, A.L., Vogels, C.B.F., Earnest, R., Lapidus, S., Ott, I.M., Moore, A.J., Muenker, M.C., Fournier, J.B., Campbell, M., Odio, C.D., Casanovas-Massana, A., Herbst, R., Shaw, A.C., Medzhitov, R., Schulz, W.L., Grubaugh, N.D., Dela Cruz, C., Farhadian, S., Ko, A.I., Omer, S.B., Iwasaki, A., 2020. Longitudinal analyses reveal immunological misfiring in severe COVID-19. Nature 584, 463–469. 10.1038/s41586-020-2588-y

Luo, O.J., Lei, W., Zhu, G., Ren, Z., Xu, Y., Xiao, C., Zhang, H., Cai, J., Luo, Z., Gao, L., Su, J., Tang, L., Guo, W., Su, H., Zhang, Z.-J., Fang, E.F., Ruan, Y., Leng, S.X., Ju, Z., Lou, H., Gao, J., Peng, N., Chen, J., Bao, Z., Liu, F., Chen, G., 2022. Multidimensional single-cell analysis of human peripheral blood reveals characteristic features of the immune system landscape in aging and frailty. Nat Aging 2, 348–364. 10.1038/s43587-022-00198-9

Mabbott, N.A., Donaldson, D.S., Ohno, H., Williams, I.R., Mahajan, A., 2013. Microfold (M) cells: important immunosurveillance posts in the intestinal epithelium. Mucosal Immunol 6, 666–677. 10.1038/mi.2013.30

Magro, F., Vieira-Coelho, M.A., Fraga, S., Serräo, M.P., Veloso, F.T., Ribeiro, T., Soares-da-Silva, P., 2002. Impaired Synthesis or Cellular Storage of Norepinephrine, Dopamine, and 5-Hydroxytryptamine in Human Inflammatory Bowel Disease. Dig Dis Sci 47, 216–224. 10.1023/A:1013256629600

Marron, T.U., Fiel, M.I., Hamon, P., Fiaschi, N., Kim, E., Ward, S.C., Zhao, Z., Kim, J., Kennedy, P., Gunasekaran, G., Tabrizian, P., Doroshow, D., Legg, M., Hammad, A., Magen, A., Kamphorst, A.O., Shareef, M., Gupta, N.T., Deering, R., Wang, W., Wang, F., Thanigaimani, P., Mani, J., Troncoso, L., Tabachnikova, A., Chang, C., Akturk, G., Buckup, M., Hamel, S., Ioannou, G., Hennequin, C., Jamal, H., Brown, H., Bonaccorso, A., Labow, D., Sarpel, U., Rosenbloom, T., Sung, M.W., Kou, B., Li, S., Jankovic, V., James, N., Hamon, S.C., Cheung, H.K., Sims, J.S., Miller, E., Bhardwaj, N., Thurston, G., Lowy, I., Gnjatic, S., Taouli, B., Schwartz, M.E., Merad, M., 2022. Neoadjuvant cemiplimab for resectable hepatocellular carcinoma: a single-arm, open-label, phase 2 trial. The Lancet Gastroenterology & Hepatology 7, 219–229. 10.1016/S2468-1253(21)00385-X

Mårtensson, J., Jain, A., Meister, A., 1990. Glutathione is required for intestinal function. Proc Natl Acad Sci U S A 87, 1715–1719. 10.1073/pnas.87.5.1715

Martin, J.C., Chang, C., Boschetti, G., Ungaro, R., Giri, M., Grout, J.A., Gettler, K., Chuang, L., Nayar, S., Greenstein, A.J., Dubinsky, M., Walker, L., Leader, A., Fine, J.S., Whitehurst, C.E., Mbow, M.L., Kugathasan, S., Denson, L.A., Hyams, J.S., Friedman, J.R., Desai, P.T., Ko, H.M., Laface, I., Akturk, G., Schadt, E.E., Salmon, H., Gnjatic, S., Rahman, A.H., Merad, M., Cho, J.H., Kenigsberg, E., 2019. Single-Cell Analysis of Crohn’s Disease Lesions Identifies a Pathogenic Cellular Module Associated with Resistance to Anti-TNF Therapy. Cell 178, 1493–1508.e20. 10.1016/j.cell.2019.08.008

Martínez-Augustin, O., de Medina, F.S., 2008. Intestinal bile acid physiology and pathophysiology. World J Gastroenterol 14, 5630–5640. 10.3748/wjg.14.5630

Mathew, D., Giles, J.R., Baxter, A.E., Oldridge, D.A., Greenplate, A.R., Wu, J.E., Alanio, C., Kuri-Cervantes, L., Pampena, M.B., D’Andrea, K., Manne, S., Chen, Z., Huang, Y.J., Reilly, J.P., Weisman, A.R., Ittner, C.A.G., Kuthuru, O., Dougherty, J., Nzingha, K., Han, N., Kim, J., Pattekar, A., Goodwin, E.C., Anderson, E.M., Weirick, M.E., Gouma, S., Arevalo, C.P., Bolton, M.J., Chen, F., Lacey, S.F., Ramage, H., Cherry, S., Hensley, S.E., Apostolidis, S.A., Huang, A.C., Vella, L.A., Unit†, T.Up.C.P., Betts, M.R., Meyer, N.J., Wherry, E.J., 2020. Deep immune profiling of COVID-19 patients reveals distinct immunotypes with therapeutic implications. Science 369. 10.1126/science.abc8511

Mauri, M., Elli, T., Caviglia, G., Uboldi, G., Azzi, M., 2017. RAWGraphs: A Visualisation Platform to Create Open Outputs, in: Proceedings of the 12th Biannual Conference on Italian SIGCHI Chapter, CHItaly ‘17. Association for Computing Machinery, New York, NY, USA, pp. 1–5. 10.1145/3125571.3125585

McOmber, M.A., Shulman, R.J., 2008. Pediatric Functional Gastrointestinal Disorders. Nutrition in Clinical Practice 23, 268–274. 10.1177/0884533608318671

Mehta, P., Porter, J.C., Manson, J.J., Isaacs, J.D., Openshaw, P.J.M., McInnes, I.B., Summers, C., Chambers, R.C., 2020. Therapeutic blockade of granulocyte macrophage colony-stimulating factor in COVID-19-associated hyperinflammation: challenges and opportunities. The Lancet Respiratory Medicine 8, 822–830. 10.1016/S2213-2600(20)30267-8

Meijer, C.J.L.M., Bosman, F.T., Lindeman, J., 1979. Evidence for Predominant Involvement of the B-Cell System in the Inflammatory Process in Crohn’s Disease. Scandinavian Journal of Gastroenterology 14, 21–32. 10.3109/00365527909179842

Mitsialis, V., Wall, S., Liu, P., Ordovas-Montanes, J., Parmet, T., Vukovic, M., Spencer, D., Field, M., McCourt, C., Toothaker, J., Bousvaros, A., Ballal, S., Bonilla, S., Fawaz, R., Fishman, L.N., Flores, A., Fox, V., Grover, A.S., Higuchi, L., Huh, S., Kahn, S., Lee, C., Mobassaleh, M., Ouahed, J., Pleskow, R.G., Regan, B., Rufo, P.A., Sabharwal, S., Silverstein, J., Verhave, M., Wolf, A., Zimmerman, L., Zitomersky, N., Allegretti, J.R., De Silva, P., Friedman, S., Hamilton, M., Korzenik, J., Makrauer, F., Norton, B.-A., Winter, R.W., Shalek, A.K., Kean, L., Horwitz, B., Goldsmith, J., Tseng, G., Snapper, S.B., Konnikova, L., 2020. Single-Cell Analyses of Colon and Blood Reveal Distinct Immune Cell Signatures of Ulcerative Colitis and Crohn’s Disease. Gastroenterology 159, 591–608.e10. 10.1053/j.gastro.2020.04.074

Miura, A., Sootome, H., Fujita, N., Suzuki, T., Fukushima, H., Mizuarai, S., Masuko, N., Ito, K., Hashimoto, A., Uto, Y., Sugimoto, T., Takahashi, H., Mitsuya, M., Hirai, H., 2021. TAS-119, a novel selective Aurora A and TRK inhibitor, exhibits antitumor efficacy in preclinical models with deregulated activation of the Myc, β-Catenin, and TRK pathways. Invest New Drugs 39, 724–735. 10.1007/s10637-020-01019-9

Moor, A.E., Harnik, Y., Ben-Moshe, S., Massasa, E.E., Rozenberg, M., Eilam, R., Bahar Halpern, K., Itzkovitz, S., 2018. Spatial Reconstruction of Single Enterocytes Uncovers Broad Zonation along the Intestinal Villus Axis. Cell 175, 1156–1167.e15. 10.1016/j.cell.2018.08.063

Müller, S., Lory, J., Corazza, N., Griffiths, G.M., Z’graggen, K., Mazzucchelli, L., Kappeler, A., Mueller, C., 1998. Activated CD4+ and CD8+ cytotoxic cells are present in increased numbers in the intestinal mucosa from patients with active inflammatory bowel disease. Am J Pathol 152, 261–268.

Muro, M., Mrowiec, A., 2015. Interleukin (IL)-1 Gene Cluster in Inflammatory Bowel Disease: Is IL-1RA Implicated in the Disease Onset and Outcome? Dig Dis Sci 60, 1126–1128. 10.1007/s10620-015-3571-6

Neurath, M.F., 2019. Targeting immune cell circuits and trafficking in inflammatory bowel disease. Nat Immunol 20, 970–979. 10.1038/s41590-019-0415-0

Nieto, P., Elosua-Bayes, M., Trincado, J.L., Marchese, D., Massoni-Badosa, R., Salvany, M., Henriques, A., Nieto, J., Aguilar-Fernández, S., Mereu, E., Moutinho, C., Ruiz, S., Lorden, P., Chin, V.T., Kaczorowski, D., Chan, C.-L., Gallagher, R., Chou, A., Planas-Rigol, E., Rubio-Perez, C., Gut, I., Piulats, J.M., Seoane, J., Powell, J.E., Batlle, E., Heyn, H., 2021. A single-cell tumor immune atlas for precision oncology. Genome Res. 31, 1913–1926. 10.1101/gr.273300.120

Ordás, I., Mould, D.R., Feagan, B.G., Sandborn, W.J., 2012. Anti-TNF monoclonal antibodies in inflammatory bowel disease: pharmacokinetics-based dosing paradigms. Clin Pharmacol Ther 91, 635–646. 10.1038/clpt.2011.328

Ordovas-Montanes, J., Dwyer, D.F., Nyquist, S.K., Buchheit, K.M., Vukovic, M., Deb, C., Wadsworth, M.H., Hughes, T.K., Kazer, S.W., Yoshimoto, E., Cahill, K.N., Bhattacharyya, N., Katz, H.R., Berger, B., Laidlaw, T.M., Boyce, J.A., Barrett, N.A., Shalek, A.K., 2018. Allergic inflammatory memory in human respiratory epithelial progenitor cells. Nature 560, 649–654. 10.1038/s41586-018-0449-8

Parikh, K., Antanaviciute, A., Fawkner-Corbett, D., Jagielowicz, M., Aulicino, A., Lagerholm, C., Davis, S., Kinchen, J., Chen, H.H., Alham, N.K., Ashley, N., Johnson, E., Hublitz, P., Bao, L., Lukomska, J., Andev, R.S., Björklund, E., Kessler, B.M., Fischer, R., Goldin, R., Koohy, H., Simmons, A., 2019. Colonic epithelial cell diversity in health and inflammatory bowel disease. Nature 567, 49–55. 10.1038/s41586-019-0992-y

Parsa, R., London, M., Rezende de Castro, T.B., Reis, B., Buissant des Amorie, J., Smith, J.G., Mucida, D., 2022. Newly recruited intraepithelial Ly6A+CCR9+CD4+ T cells protect against enteric viral infection. Immunity. 10.1016/j.immuni.2022.05.001

Pedregosa, F., Varoquaux, G., Gramfort, A., Michel, V., Thirion, B., Grisel, O., Blondel, M., Prettenhofer, P., Weiss, R., Dubourg, V., Vanderplas, J., Passos, A., Cournapeau, D., Brucher, M., Perrot, M., Duchesnay, É., 2011. Scikit-learn: Machine Learning in Python. Journal of Machine Learning Research 12, 2825–2830.

Peng, Y.-R., Shekhar, K., Yan, W., Herrmann, D., Sappington, A., Bryman, G.S., van Zyl, T., Do, M. Tri. H., Regev, A., Sanes, J.R., 2019. Molecular Classification and Comparative Taxonomics of Foveal and Peripheral Cells in Primate Retina. Cell 176, 1222–1237.e22. 10.1016/j.cell.2019.01.004

Pereira, J.P., Kelly, L.M., Xu, Y., Cyster, J.G., 2009. EBI2 mediates B cell segregation between the outer and centre follicle. Nature 460, 1122–1126. 10.1038/nature08226

Persson, E.K., Uronen-Hansson, H., Semmrich, M., Rivollier, A., Hägerbrand, K., Marsal, J., Gudjonsson, S., Håkansson, U., Reizis, B., Kotarsky, K., Agace, W.W., 2013. IRF4 Transcription-Factor-Dependent CD103+CD11b+ Dendritic Cells Drive Mucosal T Helper 17 Cell Differentiation. Immunity 38, 958–969. 10.1016/j.immuni.2013.03.009

Pliner, H.A., Shendure, J., Trapnell, C., 2019. Supervised classification enables rapid annotation of cell atlases. Nat Methods 16, 983–986. 10.1038/s41592-019-0535-3

Rajca, S., Grondin, V., Louis, E., Vernier-Massouille, G., Grimaud, J.-C., Bouhnik, Y., Laharie, D., Dupas, J.-L., Pillant, H., Picon, L., Veyrac, M., Flamant, M., Savoye, G., Jian, R., Devos, M., Paintaud, G., Piver, E., Allez, M., Mary, J.Y., Sokol, H., Colombel, J.-F., Seksik, P., 2014. Alterations in the Intestinal Microbiome (Dysbiosis) as a Predictor of Relapse After Infliximab Withdrawal in Crohn’s Disease. Inflammatory Bowel Diseases 20, 978–986. 10.1097/MIB.0000000000000036

Ramanujam, M., Steffgen, J., Visvanathan, S., Mohan, C., Fine, J.S., Putterman, C., 2020. Phoenix from the flames: Rediscovering the role of the CD40–CD40L pathway in systemic lupus erythematosus and lupus nephritis. Autoimmunity Reviews 19, 102668. 10.1016/j.autrev.2020.102668

Renoux, V.M., Zriwil, A., Peitzsch, C., Michaëlsson, J., Friberg, D., Soneji, S., Sitnicka, E., 2015. Identification of a Human Natural Killer Cell Lineage-Restricted Progenitor in Fetal and Adult Tissues. Immunity 43, 394–407. 10.1016/j.immuni.2015.07.011

Robinette, M.L., Colonna, M., 2016. Immune modules shared by innate lymphoid cells and T cells. J Allergy Clin Immunol 138, 1243–1251. 10.1016/j.jaci.2016.09.006

Roda, G., Chien Ng, S., Kotze, P.G., Argollo, M., Panaccione, R., Spinelli, A., Kaser, A., Peyrin-Biroulet, L., Danese, S., 2020. Crohn’s disease. Nat Rev Dis Primers 6, 1–19. 10.1038/s41572-020-0156-2

Roncarolo, M.G., Gregori, S., Bacchetta, R., Battaglia, M., Gagliani, N., 2018. The Biology of T Regulatory Type 1 Cells and Their Therapeutic Application in Immune-Mediated Diseases. Immunity 49, 1004–1019. 10.1016/j.immuni.2018.12.001

Rousseeuw, P.J., 1987. Silhouettes: A graphical aid to the interpretation and validation of cluster analysis. Journal of Computational and Applied Mathematics 20, 53–65. 10.1016/0377-0427(87)90125-7

Ruemmele, F.M., Veres, G., Kolho, K.L., Griffiths, A., Levine, A., Escher, J.C., Amil Dias, J., Barabino, A., Braegger, C.P., Bronsky, J., Buderus, S., Martín-de-Carpi, J., De Ridder, L., Fagerberg, U.L., Hugot, J.P., Kierkus, J., Kolacek, S., Koletzko, S., Lionetti, P., Miele, E., Navas López, V.M., Paerregaard, A., Russell, R.K., Serban, D.E., Shaoul, R., Van Rheenen, P., Veereman, G., Weiss, B., Wilson, D., Dignass, A., Eliakim, A., Winter, H., Turner, D., European Crohn’s and Colitis Organisation, European Society of Pediatric Gastroenterology, Hepatology and Nutrition, 2014. Consensus guidelines of ECCO/ESPGHAN on the medical management of pediatric Crohn’s disease. J Crohns Colitis 8, 1179–1207. 10.1016/j.crohns.2014.04.005

Sallusto, F., Lenig, D., Förster, R., Lipp, M., Lanzavecchia, A., 1999. Two subsets of memory T lymphocytes with distinct homing potentials and effector functions. Nature 401, 708–712. 10.1038/44385

Sandborn, W.J., 2014. Crohn’s Disease Evaluation and Treatment: Clinical Decision Tool. Gastroenterology 147, 702–705. 10.1053/j.gastro.2014.07.022

Santucci, N.R., Saps, M., van Tilburg, M.A., 2020. New advances in the treatment of paediatric functional abdominal pain disorders. The Lancet Gastroenterology & Hepatology 5, 316–328. 10.1016/S2468-1253(19)30256-0

Schulte-Schrepping, J., Reusch, N., Paclik, D., Baßler, K., Schlickeiser, S., Zhang, B., Krämer, B., Krammer, T., Brumhard, S., Bonaguro, L., De Domenico, E., Wendisch, D., Grasshoff, M., Kapellos, T.S., Beckstette, M., Pecht, T., Saglam, A., Dietrich, O., Mei, H.E., Schulz, A.R., Conrad, C., Kunkel, D., Vafadarnejad, E., Xu, C.-J., Horne, A., Herbert, M., Drews, A., Thibeault, C., Pfeiffer, M., Hippenstiel, S., Hocke, A., Müller-Redetzky, H., Heim, K.-M., Machleidt, F., Uhrig, A., Bosquillon de Jarcy, L., Jürgens, L., Stegemann, M., Glösenkamp, C.R., Volk, H.-D., Goffinet, C., Landthaler, M., Wyler, E., Georg, P., Schneider, M., Dang-Heine, C., Neuwinger, N., Kappert, K., Tauber, R., Corman, V., Raabe, J., Kaiser, K.M., Vinh, M.T., Rieke, G., Meisel, C., Ulas, T., Becker, M., Geffers, R., Witzenrath, M., Drosten, C., Suttorp, N., von Kalle, C., Kurth, F., Händler, K., Schultze, J.L., Aschenbrenner, A.C., Li, Y., Nattermann, J., Sawitzki, B., Saliba, A.-E., Sander, L.E., 2020. Severe COVID-19 Is Marked by a Dysregulated Myeloid Cell Compartment. Cell 182, 1419–1440.e23. 10.1016/j.cell.2020.08.001

Selin, K.A., Hedin, C.R.H., Villablanca, E.J., 2021. Immunological networks defining the heterogeneity of inflammatory bowel diseases. Journal of Crohn’s and Colitis. 10.1093/ecco-jcc/jjab085

Shalek, A.K., Satija, R., Adiconis, X., Gertner, R.S., Gaublomme, J.T., Raychowdhury, R., Schwartz, S., Yosef, N., Malboeuf, C., Lu, D., Trombetta, J.J., Gennert, D., Gnirke, A., Goren, A., Hacohen, N., Levin, J.Z., Park, H., Regev, A., 2013. Single-cell transcriptomics reveals bimodality in expression and splicing in immune cells. Nature 498, 236–240. 10.1038/nature12172

Shalek, A.K., Satija, R., Shuga, J., Trombetta, J.J., Gennert, D., Lu, D., Chen, P., Gertner, R.S., Gaublomme, J.T., Yosef, N., Schwartz, S., Fowler, B., Weaver, S., Wang, J., Wang, X., Ding, R., Raychowdhury, R., Friedman, N., Hacohen, N., Park, H., May, A.P., Regev, A., 2014. Single-cell RNA-seq reveals dynamic paracrine control of cellular variation. Nature 510, 363–369. 10.1038/nature13437

Shekhar, K., Lapan, S.W., Whitney, I.E., Tran, N.M., Macosko, E.Z., Kowalczyk, M., Adiconis, X., Levin, J.Z., Nemesh, J., Goldman, M., McCarroll, S.A., Cepko, C.L., Regev, A., Sanes, J.R., 2016. COMPREHENSIVE CLASSIFICATION OF RETINAL BIPOLAR NEURONS BY SINGLE-CELL TRANSCRIPTOMICS. Cell 166, 1308–1323.e30. 10.1016/j.cell.2016.07.054

Sido, B., Hack, V., Hochlehnert, A., Lipps, H., Herfarth, C., Dröge, W., 1998. Impairment of intestinal glutathione synthesis in patients with inflammatory bowel disease. Gut 42, 485–492. 10.1136/gut.42.4.485

Sieber, G., Herrmann, F., Zeitz, M., Teichmann, H., Rühl, H., 1984. Abnormalities of B-cell activation and immunoregulation in patients with Crohn’s disease. Gut 25, 1255–1261. 10.1136/gut.25.11.1255

Sikkema, L., Strobl, D., Zappia, L., Madissoon, E., Markov, N.S., Zaragosi, L., Ansari, M., Arguel, M., Apperloo, L., Bécavin, C., Berg, M., Chichelnitskiy, E., Chung, M., Collin, A., Gay, A.C.A., Kashani, B.H., Jain, M., Kapellos, T., Kole, T.M., Mayr, C., Papen, M. von, Peter, L., Ramírez-Suástegui, C., Schniering, J., Taylor, C., Walzthoeni, T., Xu, C., Bui, L.T., Donno, C. de, Dony, L., Guo, M., Gutierrez, A.J., Heumos, L., Huang, N., Ibarra, I., Jackson, N., Murthy, P.K.L., Lotfollahi, M., Tabib, T., Talavera-Lopez, C., Travaglini, K., Wilbrey-Clark, A., Worlock, K.B., Yoshida, M., Consortium, L.B.N., Desai, T., Eickelberg, O., Falk, C., Kaminski, N., Krasnow, M., Lafyatis, R., Nikolíc, M., Powell, J., Rajagopal, J., Rozenblatt-Rosen, O., Seibold, M.A., Sheppard, D., Shepherd, D., Teichmann, S.A., Tsankov, A., Whitsett, J., Xu, Y., Banovich, N.E., Barbry, P., Duong, T.E., Meyer, K.B., Kropski, J.A., Pe’er, D., Schiller, H.B., Tata, P.R., Schultze, J.L., Misharin, A.V., Nawijn, M.C., Luecken, M.D., Theis, F., 2022. An integrated cell atlas of the human lung in health and disease. 10.1101/2022.03.10.483747

Silverberg, M.S., Satsangi, J., Ahmad, T., Arnott, I.D.R., Bernstein, C.N., Brant, S.R., Caprilli, R., Colombel, J.-F., Gasche, C., Geboes, K., Jewell, D.P., Karban, A., Loftus, E.V., Peña, A.S., Riddell, R.H., Sachar, D.B., Schreiber, S., Steinhart, A.H., Targan, S.R., Vermeire, S., Warren, B.F., 2005. Toward an integrated clinical, molecular and serological classification of inflammatory bowel disease: report of a Working Party of the 2005 Montreal World Congress of Gastroenterology. Can J Gastroenterol 19 Suppl A, 5A-36A. 10.1155/2005/269076

Simpson, E.H., 1949. Measurement of Diversity. Nature 163, 688–688. 10.1038/163688a0

Smillie, C.S., Biton, M., Ordovas-Montanes, J., Sullivan, K.M., Burgin, G., Graham, D.B., Herbst, R.H., Rogel, N., Slyper, M., Waldman, J., Sud, M., Andrews, E., Velonias, G., Haber, A.L., Jagadeesh, K., Vickovic, S., Yao, J., Stevens, C., Dionne, D., Nguyen, L.T., Villani, A.-C., Hofree, M., Creasey, E.A., Huang, H., Rozenblatt-Rosen, O., Garber, J.J., Khalili, H., Desch, A.N., Daly, M.J., Ananthakrishnan, A.N., Shalek, A.K., Xavier, R.J., Regev, A., 2019. Intra- and Inter-cellular Rewiring of the Human Colon during Ulcerative Colitis. Cell 178, 714–730.e22. 10.1016/j.cell.2019.06.029

Sootome, H., Miura, A., Masuko, N., Suzuki, T., Uto, Y., Hirai, H., 2020. Aurora A Inhibitor TAS-119 Enhances Antitumor Efficacy of Taxanes In Vitro and In Vivo: Preclinical Studies as Guidance for Clinical Development and Trial Design. Mol Cancer Ther 19, 1981–1991. 10.1158/1535-7163.MCT-20-0036

Souza, H.S., Elia, C.C.S., Spencer, J., MacDonald, T.T., 1999. Expression of lymphocyte-endothelial receptor-ligand pairs, α4β7/MAdCAM-1 and OX40/OX40 ligand in the colon and jejunum of patients with inflammatory bowel disease. Gut 45, 856–863. 10.1136/gut.45.6.856

Stappenbeck, T.S., McGovern, D.P.B., 2017. Paneth Cell Alterations in the Development and Phenotype of Crohn’s Disease. Gastroenterology, Inflammatory Bowel Disease 2017: Innovations and Changing Paradigms 152, 322–326. 10.1053/j.gastro.2016.10.003

Stevens, T.W., Matheeuwsen, M., Lönnkvist, M.H., Parker, C.E., Wildenberg, M.E., Gecse, K.B., D’Haens, G.R., 2018. Systematic review: predictive biomarkers of therapeutic response in inflammatory bowel disease-personalised medicine in its infancy. Aliment Pharmacol Ther 48, 1213–1231. 10.1111/apt.15033

Stuart, T., Butler, A., Hoffman, P., Hafemeister, C., Papalexi, E., Mauck, W.M., Hao, Y., Stoeckius, M., Smibert, P., Satija, R., 2019. Comprehensive Integration of Single-Cell Data. Cell 177, 1888–1902.e21. 10.1016/j.cell.2019.05.031

Su, Y., Chen, D., Yuan, D., Lausted, C., Choi, J., Dai, C.L., Voillet, V., Duvvuri, V.R., Scherler, K., Troisch, P., Baloni, P., Qin, G., Smith, B., Kornilov, S.A., Rostomily, C., Xu, A., Li, J., Dong, S., Rothchild, A., Zhou, J., Murray, K., Edmark, R., Hong, S., Heath, J.E., Earls, J., Zhang, R., Xie, J., Li, S., Roper, R., Jones, L., Zhou, Y., Rowen, L., Liu, R., Mackay, S., O’Mahony, D.S., Dale, C.R., Wallick, J.A., Algren, H.A., Zager, M.A., Wei, W., Price, N.D., Huang, S., Subramanian, N., Wang, K., Magis, A.T., Hadlock, J.J., Hood, L., Aderem, A., Bluestone, J.A., Lanier, L.L., Greenberg, P.D., Gottardo, R., Davis, M.M., Goldman, J.D., Heath, J.R., 2020. Multi-Omics Resolves a Sharp Disease-State Shift between Mild and Moderate COVID-19. Cell 183, 1479–1495.e20. 10.1016/j.cell.2020.10.037

Sullivan, Z.A., Khoury-Hanold, W., Lim, J., Smillie, C., Biton, M., Reis, B.S., Zwick, R.K., Pope, S.D., Israni-Winger, K., Parsa, R., Philip, N.H., Rashed, S., Palm, N., Wang, A., Mucida, D., Regev, A., Medzhitov, R., 2021. γδ T cells regulate the intestinal response to nutrient sensing. Science 371. 10.1126/science.aba8310

Sýkora, J., Pomahačová, R., Kreslová, M., Cvalínová, D., Štych, P., Schwarz, J., 2018. Current global trends in the incidence of pediatric-onset inflammatory bowel disease. World J Gastroenterol 24, 2741–2763. 10.3748/wjg.v24.i25.2741

Takayama, T., Kamada, N., Chinen, H., Okamoto, S., Kitazume, M.T., Chang, J., Matuzaki, Y., Suzuki, S., Sugita, A., Koganei, K., Hisamatsu, T., Kanai, T., Hibi, T., 2010. Imbalance of NKp44+NKp46− and NKp44−NKp46+ Natural Killer Cells in the Intestinal Mucosa of Patients With Crohn’s Disease. Gastroenterology 139, 882–892.e3. 10.1053/j.gastro.2010.05.040

Tasic, B., Yao, Z., Graybuck, L.T., Smith, K.A., Nguyen, T.N., Bertagnolli, D., Goldy, J., Garren, E., Economo, M.N., Viswanathan, S., Penn, O., Bakken, T., Menon, V., Miller, J., Fong, O., Hirokawa, K.E., Lathia, K., Rimorin, C., Tieu, M., Larsen, R., Casper, T., Barkan, E., Kroll, M., Parry, S., Shapovalova, N.V., Hirschstein, D., Pendergraft, J., Sullivan, H.A., Kim, T.K., Szafer, A., Dee, N., Groblewski, P., Wickersham, I., Cetin, A., Harris, J.A., Levi, B.P., Sunkin, S.M., Madisen, L., Daigle, T.L., Looger, L., Bernard, A., Phillips, J., Lein, E., Hawrylycz, M., Svoboda, K., Jones, A.R., Koch, C., Zeng, H., 2018. Shared and distinct transcriptomic cell types across neocortical areas. Nature 563, 72–78. 10.1038/s41586-018-0654-5

Thiriot, A., Perdomo, C., Cheng, G., Novitzky-Basso, I., McArdle, S., Kishimoto, J.K., Barreiro, O., Mazo, I., Triboulet, R., Ley, K., Rot, A., von Andrian, U.H., 2017. Differential DARC/ACKR1 expression distinguishes venular from non-venular endothelial cells in murine tissues. BMC Biology 15, 45. 10.1186/s12915-017-0381-7

Travaglini, K.J., Nabhan, A.N., Penland, L., Sinha, R., Gillich, A., Sit, R.V., Chang, S., Conley, S.D., Mori, Y., Seita, J., Berry, G.J., Shrager, J.B., Metzger, R.J., Kuo, C.S., Neff, N., Weissman, I.L., Quake, S.R., Krasnow, M.A., 2020. A molecular cell atlas of the human lung from single-cell RNA sequencing. Nature 587, 619–625. 10.1038/s41586-020-2922-4

Turner, D., Griffiths, A.M., Walters, T.D., Seah, T., Markowitz, J., Pfefferkorn, M., Keljo, D., Waxman, J., Otley, A., LeLeiko, N.S., Mack, D., Hyams, J., Levine, A., 2012. Mathematical weighting of the pediatric Crohn’s disease activity index (PCDAI) and comparison with its other short versions. Inflamm Bowel Dis 18, 55–62. 10.1002/ibd.21649

Turner, D., Levine, A., Walters, T.D., Focht, G., Otley, A., López, V.N., Koletzko, S., Baldassano, R., Mack, D., Hyams, J., Griffiths, A.M., 2017. Which PCDAI Version Best Reflects Intestinal Inflammation in Pediatric Crohn Disease? Journal of Pediatric Gastroenterology and Nutrition 64, 254–260. 10.1097/MPG.0000000000001227

Uzzan, M., Martin, J.C., Mesin, L., Livanos, A.E., Castro-Dopico, T., Huang, R., Petralia, F., Magri, G., Kumar, S., Zhao, Q., Rosenstein, A.K., Tokuyama, M., Sharma, K., Ungaro, R., Kosoy, R., Jha, D., Fischer, J., Singh, H., Keir, M.E., Ramamoorthi, N., Gorman, W.E.O., Cohen, B.L., Rahman, A., Cossarini, F., Seki, A., Leyre, L., Vaquero, S.T., Gurunathan, S., Grasset, E.K., Losic, B., Dubinsky, M., Greenstein, A.J., Gottlieb, Z., Legnani, P., George, J., Irizar, H., Stojmirovic, A., Brodmerkel, C., Kasarkis, A., Sands, B.E., Furtado, G., Lira, S.A., Tuong, Z.K., Ko, H.M., Cerutti, A., Elson, C.O., Clatworthy, M.R., Merad, M., Suárez-Fariñas, M., Argmann, C., Hackney, J.A., Victora, G.D., Randolph, G.J., Kenigsberg, E., Colombel, J.F., Mehandru, S., 2022. Ulcerative colitis is characterized by a plasmablast-skewed humoral response associated with disease activity. Nat Med 28, 766–779. 10.1038/s41591-022-01680-y

van der Flier, L.G., Clevers, H., 2009. Stem Cells, Self-Renewal, and Differentiation in the Intestinal Epithelium. Annu. Rev. Physiol. 71, 241–260. 10.1146/annurev.physiol.010908.163145

Verstockt, B., Verstockt, S., Dehairs, J., Ballet, V., Blevi, H., Wollants, W.-J., Breynaert, C., Van Assche, G., Vermeire, S., Ferrante, M., 2019. Low TREM1 expression in whole blood predicts anti-TNF response in inflammatory bowel disease. EBioMedicine 40, 733–742. 10.1016/j.ebiom.2019.01.027

Victora, G.D., Schwickert, T.A., Fooksman, D.R., Kamphorst, A.O., Meyer-Hermann, M., Dustin, M.L., Nussenzweig, M.C., 2010. Germinal Center Dynamics Revealed by Multiphoton Microscopy with a Photoactivatable Fluorescent Reporter. Cell 143, 592–605. 10.1016/j.cell.2010.10.032

von Moltke, J., Ji, M., Liang, H.-E., Locksley, R.M., 2016. Tuft-cell-derived IL-25 regulates an intestinal ILC2–epithelial response circuit. Nature 529, 221–225. 10.1038/nature16161

Wen, J., Rawls, J.F., 2020. Feeling the Burn: Intestinal Epithelial Cells Modify Their Lipid Metabolism in Response to Bacterial Fermentation Products. Cell Host & Microbe 27, 314–316. 10.1016/j.chom.2020.02.009

Whitsett, J.A., Kalin, T.V., Xu, Y., Kalinichenko, V.V., 2019. Building and Regenerating the Lung Cell by Cell. Physiol Rev 99, 513–554. 10.1152/physrev.00001.2018

Yarur, A.J., Jain, A., Sussman, D.A., Barkin, J.S., Quintero, M.A., Princen, F., Kirkland, R., Deshpande, A.R., Singh, S., Abreu, M.T., 2016. The association of tissue anti-TNF drug levels with serological and endoscopic disease activity in inflammatory bowel disease: the ATLAS study. Gut 65, 249–255. 10.1136/gutjnl-2014-308099

Ye, Y., Manne, S., Treem, W.R., Bennett, D., 2020. Prevalence of Inflammatory Bowel Disease in Pediatric and Adult Populations: Recent Estimates From Large National Databases in the United States, 2007-2016. Inflamm Bowel Dis 26, 619–625. 10.1093/ibd/izz182

Yilmaz, B., Juillerat, P., Øyås, O., Ramon, C., Bravo, F.D., Franc, Y., Fournier, N., Michetti, P., Mueller, C., Geuking, M., Pittet, V.E.H., Maillard, M.H., Rogler, G., Wiest, R., Stelling, J., Macpherson, A.J., 2019. Microbial network disturbances in relapsing refractory Crohn’s disease. Nat Med 25, 323–336. 10.1038/s41591-018-0308-z

Zeisel, A., Hochgerner, H., Lönnerberg, P., Johnsson, A., Memic, F., van der Zwan, J., Häring, M., Braun, E., Borm, L.E., La Manno, G., Codeluppi, S., Furlan, A., Lee, K., Skene, N., Harris, K.D., Hjerling-Leffler, J., Arenas, E., Ernfors, P., Marklund, U., Linnarsson, S., 2018. Molecular Architecture of the Mouse Nervous System. Cell 174, 999–1014.e22. 10.1016/j.cell.2018.06.021

Ziegler, C.G.K., Allon, S.J., Nyquist, S.K., Mbano, I.M., Miao, V.N., Tzouanas, C.N., Cao, Y., Yousif, A.S., Bals, J., Hauser, B.M., Feldman, J., Muus, C., Wadsworth, M.H., Kazer, S.W., Hughes, T.K., Doran, B., Gatter, G.J., Vukovic, M., Taliaferro, F., Mead, B.E., Guo, Z., Wang, J.P., Gras, D., Plaisant, M., Ansari, M., Angelidis, I., Adler, H., Sucre, J.M.S., Taylor, C.J., Lin, B., Waghray, A., Mitsialis, V., Dwyer, D.F., Buchheit, K.M., Boyce, J.A., Barrett, N.A., Laidlaw, T.M., Carroll, S.L., Colonna, L., Tkachev, V., Peterson, C.W., Yu, A., Zheng, H.B., Gideon, H.P., Winchell, C.G., Lin, P.L., Bingle, C.D., Snapper, S.B., Kropski, J.A., Theis, F.J., Schiller, H.B., Zaragosi, L.-E., Barbry, P., Leslie, A., Kiem, H.-P., Flynn, J.L., Fortune, S.M., Berger, B., Finberg, R.W., Kean, L.S., Garber, M., Schmidt, A.G., Lingwood, D., Shalek, A.K., Ordovas-Montanes, J., Banovich, N., Barbry, P., Brazma, A., Desai, T., Duong, T.E., Eickelberg, O., Falk, C., Farzan, M., Glass, I., Haniffa, M., Horvath, P., Hung, D., Kaminski, N., Krasnow, M., Kropski, J.A., Kuhnemund, M., Lafyatis, R., Lee, H., Leroy, S., Linnarson, S., Lundeberg, J., Meyer, K., Misharin, A., Nawijn, M., Nikolic, M.Z., Ordovas-Montanes, J., Pe’er, D., Powell, J., Quake, S., Rajagopal, J., Tata, P.R., Rawlins, E.L., Regev, A., Reyfman, P.A., Rojas, M., Rosen, O., Saeb-Parsy, K., Samakovlis, C., Schiller, H., Schultze, J.L., Seibold, M.A., Shalek, A.K., Shepherd, D., Spence, J., Spira, A., Sun, X., Teichmann, S., Theis, F., Tsankov, A., van den Berge, M., von Papen, M., Whitsett, J., Xavier, R., Xu, Y., Zaragosi, L.-E., Zhang, K., 2020. SARS-CoV-2 Receptor ACE2 Is an Interferon-Stimulated Gene in Human Airway Epithelial Cells and Is Detected in Specific Cell Subsets across Tissues. Cell 181, 1016–1035.e19. 10.1016/j.cell.2020.04.035

Ziegler, C.G.K., Miao, V.N., Owings, A.H., Navia, A.W., Tang, Y., Bromley, J.D., Lotfy, P., Sloan, M., Laird, H., Williams, H.B., George, M., Drake, R.S., Christian, T., Parker, A., Sindel, C.B., Burger, M.W., Pride, Y., Hasan, M., Abraham, G.E., Senitko, M., Robinson, T.O., Shalek, A.K., Glover, S.C., Horwitz, B.H., Ordovas-Montanes, J., 2021. Impaired local intrinsic immunity to SARS-CoV-2 infection in severe COVID-19. Cell. 10.1016/j.cell.2021.07.023

